# Conceptualising the context and mechanisms for tackling loneliness in older adults through interventions: A Critical Interpretive Synthesis

**DOI:** 10.1101/2025.01.06.25320003

**Authors:** John Ratcliffe, Faith Matcham, Erika Molteni, Michela Antonelli, Jessica Rees, Yu Shi, Jingqi Liu, Sebastian Ourselin, Anthea Tinker, Yi Zhou, Wei Liu

## Abstract

**Background:** Loneliness in later life has been widely associated with poor mental and physical health. However, despite many reviews, current evidence does not provide a clear picture of how to tackle loneliness in older adults through interventions. Research examining the contexts and mechanisms for reducing loneliness through interventions is required to identify how, why, and for who interventions work or do not work.

**Methods:** A critical interpretive synthesis, a review method designed to rigorously and reflexively re-examine existing literature, was conducted to re-consider and identify what matters in interventions. This was enacted through a broad search strategy incorporating database searches, contacting experts, reference-chaining, and team discussions, enabling a flexible and iterative review able to generate new theory.

**Results:** 274 papers were included in the analysis. We identified four dimensions to whether and how interventions worked. *Intended outcome* mirrored different theoretical perspectives on how to reduce loneliness. Four intended outcomes were identified: cognitive change; improved social connections; physiological change; and a more meaningful life. The second dimension was the *level* of an intervention - whether it intended to intervene at a micro, meso, or macro level. Thirdly, 13 *positive features* of interventions were noted to have been evidenced to lead to or constitute more effective interventions. These were interventions that are organised, adaptable, have good staff/volunteers, provide support to attend, routinised, built on shared interests/identities, personalised, culturally aware, co-produced, involve active participation, strengths-based, lasting, and targeted at the appropriate people. Lastly, nine *emotions that represent less loneliness* were identified: a feeling of reciprocated interactions; emotional and social support; belonging; perspective; self-efficacy; mattering; pride; purpose; and empowerment.

**Conclusions:** Interventions and evaluations should look beyond comparing activities to see which are best and recognise that multidimensionality and variation is vital for individuals and communities with different needs. Identifying what the intended outcome of an intervention is, what level of society it intends to impact, and whether it actions the positive features identified in this study, can be employed to better target the variety of emotions that represent less loneliness.

## Background

Loneliness is a significant public health concern associated with depression,^1^ cardiovascular disorder,^2^ early onset dementia,^3,4^ and end-of-life symptoms such as pain and breathlessness.^5^ Age-related situations such as retirement and bereavement increase the odds of loneliness,^6,7^ and stereotypical associations between loneliness and ageing may contribute to its stigma and worsen its impact.^8,9^ Despite a wealth of research and reviews, a clear understanding of which interventions are most effective at tackling loneliness in older adults is yet to be identified. This has led to calls for research that can investigate the context and mechanisms of loneliness interventions.^10,11^

Meta-analytical reviews focused on the activities in an intervention have found inconsistent evidence. Among older adults in long-term care, Hoang et al^12^ found that animal therapy and videoconferencing showed the greatest positive effect. Veronese et al,^13^ on the other hand, found that among a general cross-section of older adults, meditation/mindfulness, social cognitive training, and social support interventions displayed a statistically significant positive effect, but that technological and animal therapy interventions did *not*. In both these reviews, effect sizes were consistently small and positive regardless of statistical significance, leading Veronese et al^13^ to suggest that simply providing any intervention may do enough good to register a small impact.

Pre and post-test methods, which constitute the focus of reviews such as these, can only investigate whether the interventions worked, with little way to investigate why. Some studies employing a qualitative follow up, though, have found that participants gave a different rationale for its efficacy to that which the intervention provider had intended.^14,15^ Furthermore, reviews employing a broader methodology have highlighted that the success of an intervention may originate from factors other than the activity it provides. Gardiner et al^16^ concluded that adaptability, a community development approach, and productive engagement (i.e., meaningful activities) resulted in effective interventions. Boulton et al^17^ found that successful interventions included some form of pastoral guidance, and enabled participants to speak freely, form close relationships, and share experiences/characteristics. Morrish et al^18^ conclude that effective interventions promote between session interaction, consist of clear learning mechanisms, facilitate active participation, contain opportunities for group or facilitator interaction, and, across different interventions, provide opportunities to take part in activities that include a variety of teaching/learning styles.

Such reviews usefully focus on the quality of an intervention participant’s experience. However, they provide disparate conclusions that are not easily actioned. Furthermore, some of their recommendations, such as a need for a variety of teaching/learning styles,^18^ and for interventions where people share characteristics,^17^ suggest a need for plurality in the wider policy and practice response. This is logical as different causes of loneliness, and even different forms of it, are well established in loneliness studies.^19–21^ It is vital, then, for interventions to provide a contextualised response to the specific problems of different individuals and groups. Work has highlighted that interventions with a theoretical framework for success are more effective,^22^ but current literature does not provide guidance on how to construct a contextualised and mechanistic interventions.^23^ The current paper presents a Critical Interpretative Synthesis (CIS) re-examining the existing literature on loneliness interventions for older adults, and uses it to build a theoretical framework that can inform the formation of contextualised and mechanistic research, policy, and practice.

## Method

CIS is a review method that aims to iteratively identify and critique relevant literature.^24^ Built from meta ethnography, it does not consist of a wholly a priori search strategy and analysis plan. Rather, it utilises an inductive approach in which the goal is to build thematic interpretations of the data called ‘synthetic constructs’, then create ‘synthesising arguments’ based on a critical reinterpretation of the synthetic constructs.^24,25^ This inductive and reflexive approach enables CIS to generate new theoretical perspectives.^26^ The flexibility inherent to CIS enables it to examine a large and diverse set of papers, allowing it to generate synthetic constructs from across methodologies and disciplines.^25^ It is also useful for reviewing ‘challenging’ areas such as mental health (and loneliness) as it can analyse and interpret beyond the data presented in the articles it collates.^27^ CIS is useful for the current review as we are not aiming summarise or integrate existing data, but to identify and critique the theoretical mechanisms implicit to interventions for loneliness in older adults.

The inductive and flexible nature of CIS has led to criticisms that it lacks transparency, trustworthiness, and systematicity. ^25^ Most notably, the lack of an a priori search strategy can render it indistinct from a narrative review.^28^ Constructing an a priori strategy, though, is antithetical to a CIS as this removes its ability to flexibly and iteratively identify relevant literature from which new theoretical ideas can be constructed.^24,25^ It is therefore vital to present what Noyes et al^29^ call an ‘evidence to decision framework’ without losing the critical and iterative features that make CIS appropriate for the current review. Templier and Pare^28^ argue that theory driven reviews can improve their transparency by adopting structures in which the flexibility and iteration are stated in the structure. To do so, we will structure the current paper using terminology recommended by Preferred Reporting Items for Systematic reviews and Meta-Analyses (PRISMA)^30^, and conduct the review using Depraetere et al’s^25^ six sequential processes that define CIS:

1. An open research question.
2. A broad and flexible search strategy that is built atop a more structured approach.
3. A flexible method of selecting literature based on relevance rather than systematic criteria.
4. A consideration of quality appraisal based on a paper’s theoretical contribution.
5. Data extraction that is iterative, flexible, and reflexive.
6. The formulation of synthetic constructs that are then refined into synthesising arguments.

### Search strategy

CIS aims to generate new concepts from a wide array of literature, therefore the current review did not employ the kind of rigid strategy common to systematic reviews.^24,25^ Nevertheless, to promote transparency, a search strategy with defined space for flexibility and iteration was constructed.^28^ This consisted of four components.

1. Researchers who work in studies related to loneliness, and were known to the lead author, were contacted by email to give their opinions on theoretical mechanisms and direct the review team to literature they believed was relevant.^31^ Those who engaged are credited with an acknowledgement. This aimed to promote a broad strategy in which varied perspectives could be considered.^24^
2. Database searches were conducted in Medline, Scopus, and Psycinfo. Table 1 displays the full search strings. These databases were chosen because of their size and focus on medical and social science research. This provided the structured foundations of the search strategy.^25^
3. Reference chaining, based on theoretically relevant literature cited in the initial texts retrieved for full text screening, was employed to broaden the search strategy. ^25^
4. Relevant articles identified from other sources were added at any stage during the review. These were expected to arise via reflexive thought and team discussion. This allowed the review it to go beyond the confines of an a priori search string. ^24,25^

**Table 1:** Search strings in Medline, Psychinfo, and Scopus.

| Database | Search strings (entered on 30/07/2024) |
| --- | --- |
| Medline and psycinfo (Ovid) | (lonel* and intervention* and (old* or "later life")).ab,ti.<br>Limit to English<br>Limit to humans<br>(lonel* and intervention* and (old* or "later life")).ab,ti.<br>Limit to English<br>Limit to humans<br>Remove duplicates |
| Scopus | TITLE-ABS-KEY ( ( lonel* AND intervention* AND ( old* OR "later life" ) ) . ) AND ( LIMIT-TO ( LANGUAGE , "English" ) ) |

### Selection criteria

The selection criteria were intended to facilitate a transparent approach without losing the inductive and flexible approach core to CIS. The four criteria relayed below were kept broad enough to allow relevant data to be included from unexpected sources.^24^ Any research method was acceptable, including systematic reviews, and reviews of reviews, as they might have new information or ideas that were not reported in the original studies. Theoretical pieces with relevant perspectives or ideas were also permitted. The inclusion criteria require a stated focus on older adults, rather than a defined age range, as this allowed the review to avoid arbitrary inclusions/exclusions. The relevance of age was reflexively considered throughout the analysis and write-up. English papers were sought to aid the manageability of the project, and peer reviewed papers were specified to avoid excessive amounts of poorly reasoned/evidenced perspectives.^32^ To be eligible for inclusion, the papers must:

1. State or provide evidence for a theoretical perspective on why/how interventions for loneliness work or do not work.
2. Include a substantive focus on older adults.
3. Be written in English.
4. Be peer reviewed.

### Screening

The following screening process was followed:

1. Records from all four search strategy components were uploaded to Endnote and duplications removed. Records were continuously added as they arose.
2. Title and abstract screening. Articles that did not meet the four criteria above were removed.
3. The full text of all remaining articles were examined to identify whether they relayed or implied a meaningful theoretical mechanism explaining why interventions do or not work.
4. The second author read a random selection of 10% of the full texts retrieved. Differences were discussed, and the remaining 90% of full texts were re-examined as appropriate.
5. As the analysis progressed, the final list of included/excluded articles were reviewed for relevance to the synthetic constructs and synthesising arguments. Articles added/removed after step 4 were noted and the reason for addition/removal was recorded.

### Quality appraisal

CIS prioritises papers that are relevant rather than papers that meet particular methodological standards,^24^ and the inclusion of papers from a wide variety of methods means included papers often draw on a wide variety ontological and epistemological perspectives.^33^ It is therefore common for CIS not to employ quality appraisal tools - a study of limited quality according to a standardised tool could still contain useful perspectives, whereas a high-quality study may contain no meaningful theoretical contribution to the review. ^24,25,34^ The use of standardised quality appraisal tools was therefore considered inappropriate for the current review.

Templier and Pare^28^ suggest theory development reviews have been insufficiently explicit about quality appraisal, leading to distrust in their findings. It is for this reason that point 4 of Depraetere et al’s^25^ six processes states that even if quality appraisal is not conducted, it is necessary to discuss and justify the action taken (or not taken). In the current review, two actions were taken. First, inclusion was limited to ‘trustworthy’ materials in peer-reviewed journals.^32^ Second, Dixon-woods et al’s^24^ five ‘appraisal prompts’ were used to identify ‘fatally flawed’ studies – studies that are easily identified as extremely poor and unsuitable even for CIS.

1. Are the aims and objectives of the research clearly stated?
2. Is the research design clearly specified and appropriate for the aims and objectives of the research?
3. Do the researchers provide a clear account of the process by which their findings we reproduced?
4. Do the researchers display enough data to support their interpretations and conclusions?
5. Is the method of analysis appropriate and adequately explicated?

### Data extraction and analysis

CIS consists of two key stages of analysis: ^24,25^ i) the creation of synthetic constructs, akin to theme coding in a qualitative thematic analysis; and ii), using these to construct ‘synthesising arguments’ that describe the newly generated theory. CIS typically aims to allow the important theory to emerge organically rather than through systematic criteria.^24,35^ Nevertheless, to improve the evidence to decision framework, and to facilitate reflexivity during analysis, two sets of tabular findings were constructed.^29,31^ These were constructed within a 6-stage procedure designed to detail the process without sacrificing flexibility and reflexivity.

1. When reviewing the full texts, included articles were entered into a table detailing the title, study method and population (if applicable), and an overarching description of the relevant information it gives. Analysing different methods will be handled by stating the relevant data the study provides regardless of its methodology.
2. The second author read the table constructed in step 1, and 10% of the papers identified for full text review. Differences of inclusion and interpretation were discussed, and the selection process revised as appropriate.
3. Synthetic constructs were identified thematically via the full text readings and examination of the table constructed in steps 1 and 2. ^26^ A set of tables were constructed to detail how the included articles provide relevant data or theory for the synthetic constructs.
4. Synthesising arguments were constructed by reflecting on the meaning and ramifications of the synthetic constructs.
5. The quality appraisal was conducted. It was delayed until this stage as methodologically weak papers may still provide relevant insights. ^26^
6. Meetings and feedback were used to improve how logical, meaningful, and well evidenced the synthesising arguments are.
7. The two sets of tabular data, and the synthesising arguments, were reviewed and revised until the current article was complete.

## Results

### Searches

274 articles were included in the analysis. Figure 1 displays the stages conducted in the literature review and how many articles were retained at each stage. Table 2 displays the full list of included articles, and the reason for inclusion. The second author read 61 papers (10% of the papers retrieved for full text reviewing), resulting in 11 disagreements. The disagreements were all papers that included an implied mechanism by which the intervention intended to tackle loneliness that was not significantly discussed or investigated. We decided these were unlikely to add to the analysis, therefore all texts that did not significantly discuss or investigate the theoretical mechanism for tackling loneliness were removed unless the implicit mechanism was particularly innovative or rare. We did not seek access to articles that were unavailable as we believed theoretical saturation was sufficiently attained via the large number of accessible papers. ^29^ No papers were removed due to being ‘fatally flawed’. ^26^

**Figure 1.**
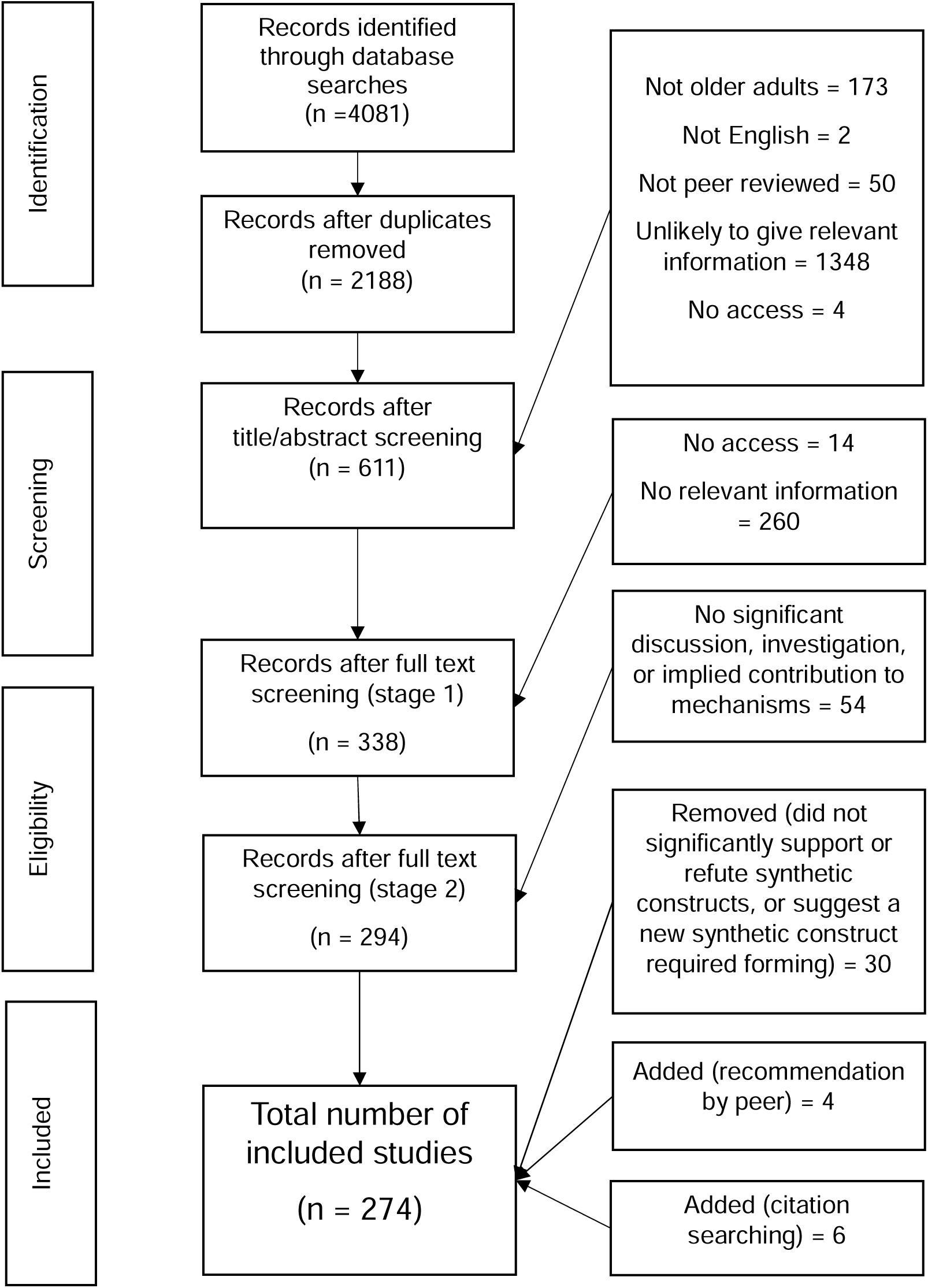
PRISMA style chart displaying the selection process.

**Table 2:**
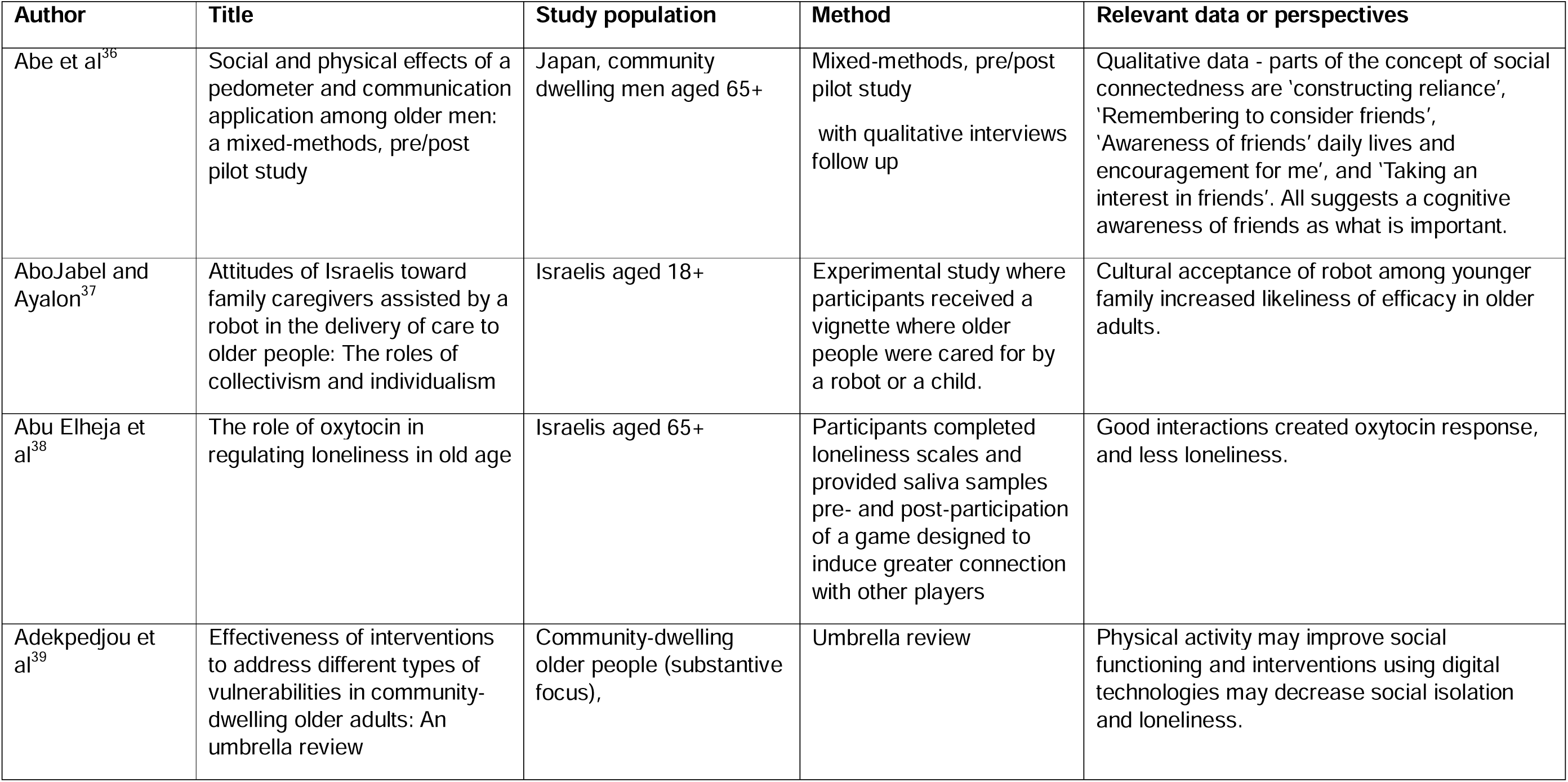
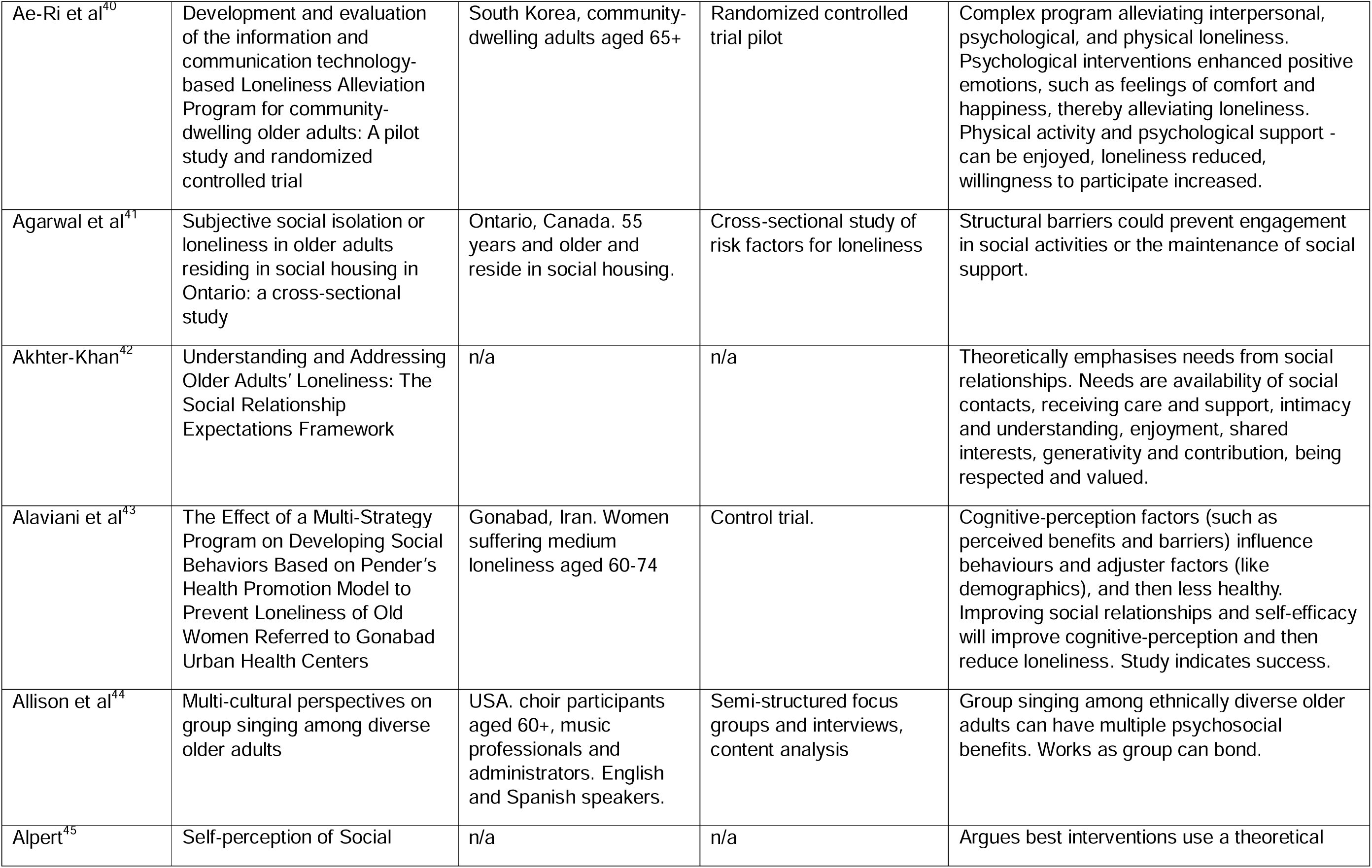
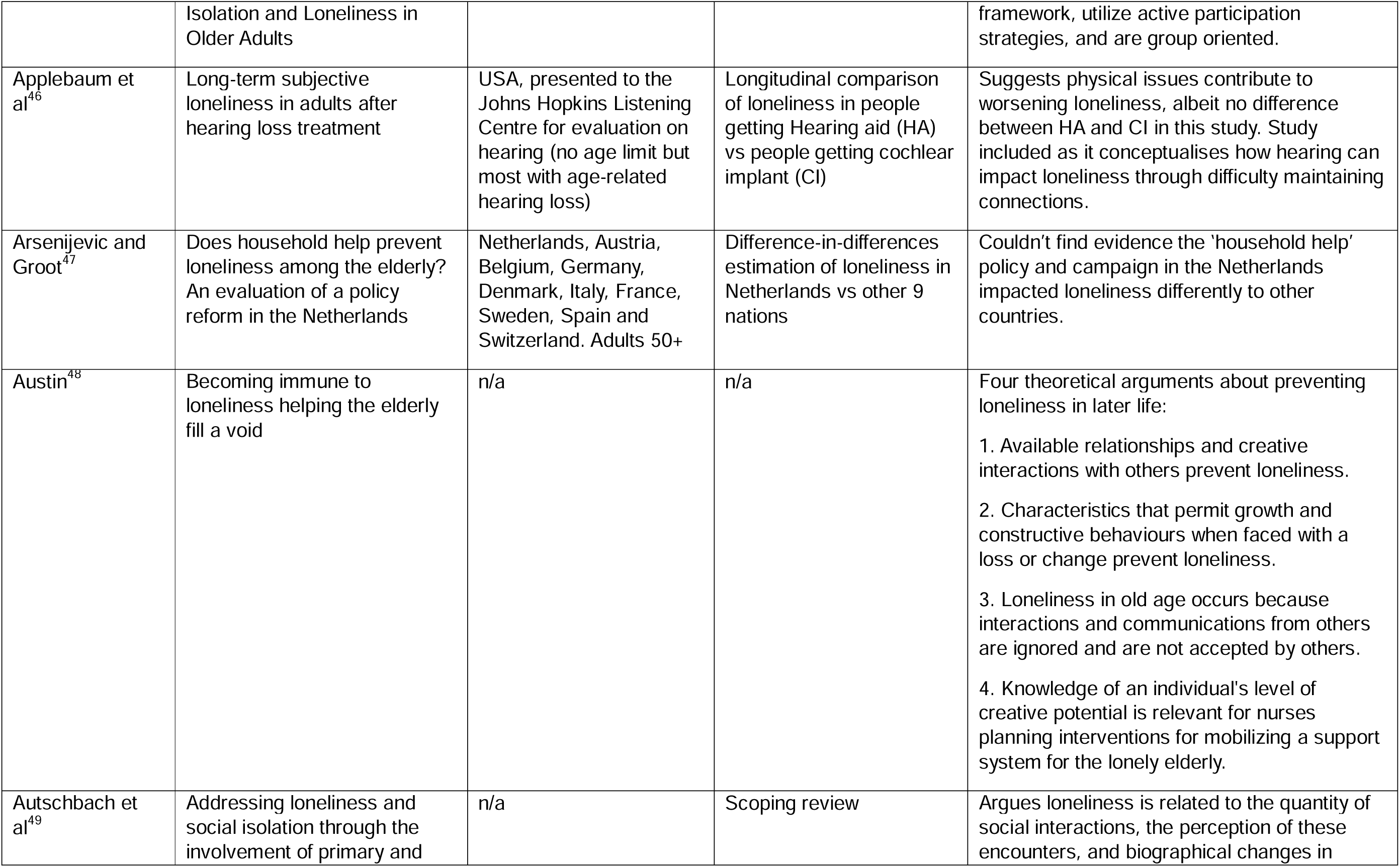
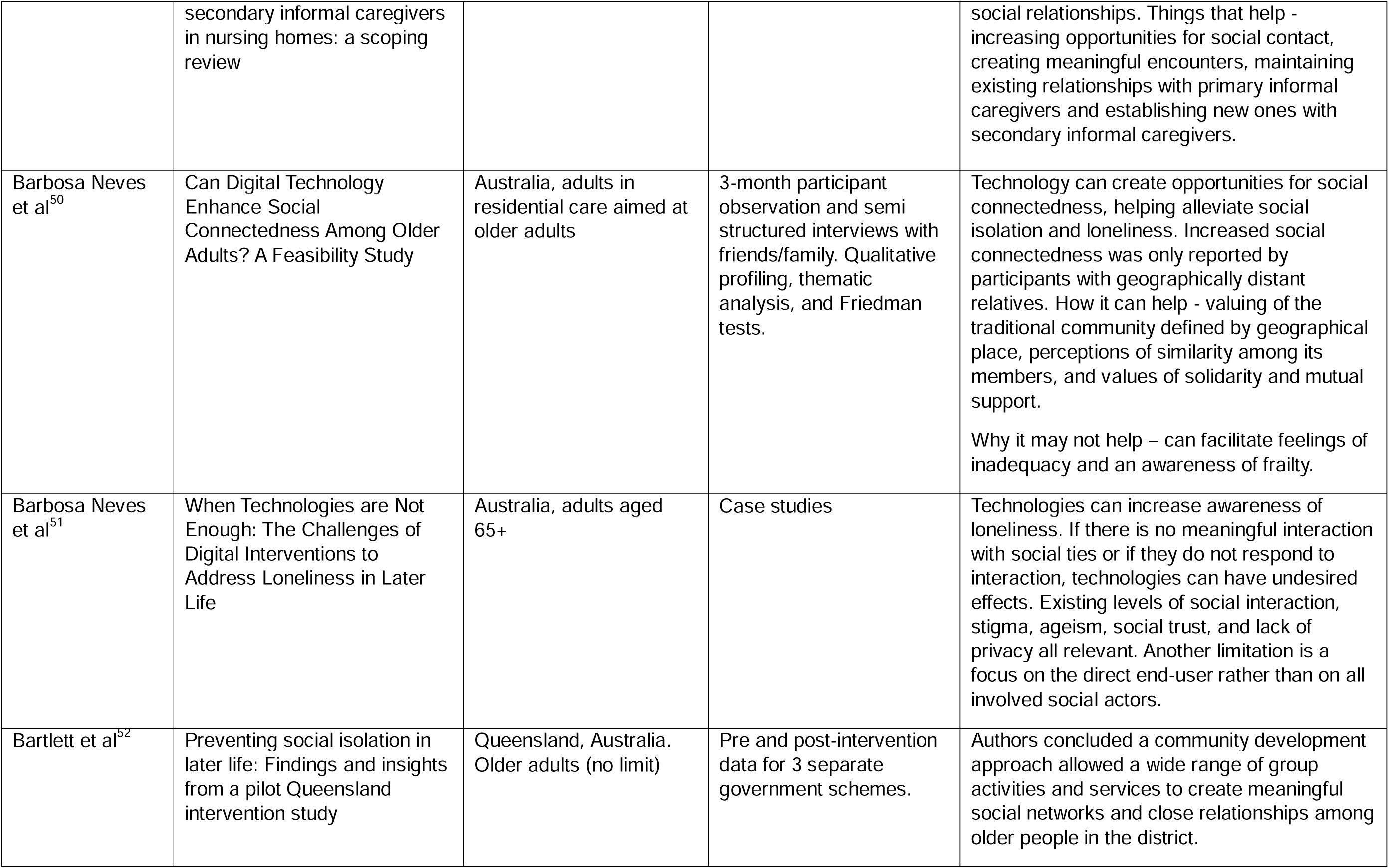
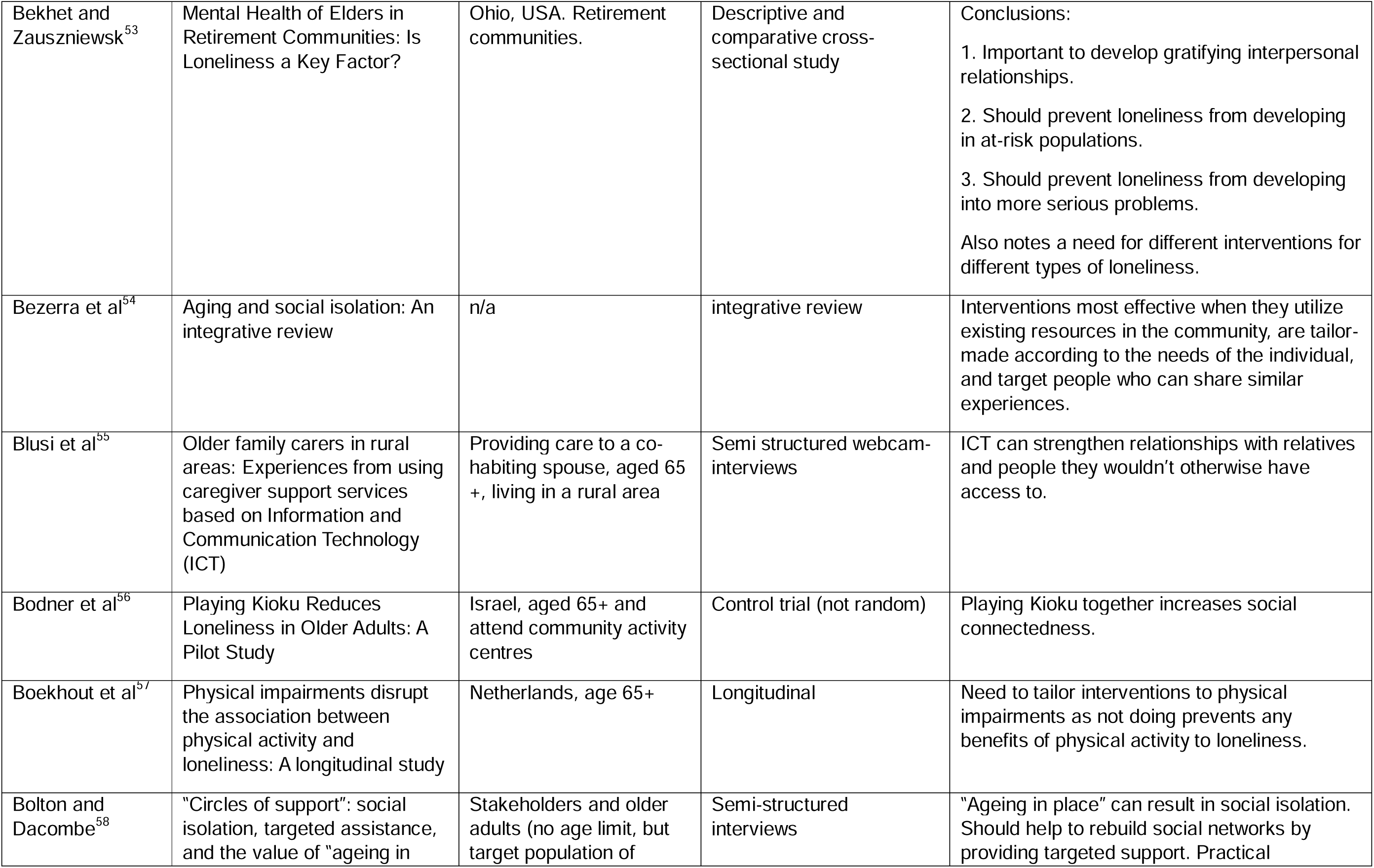
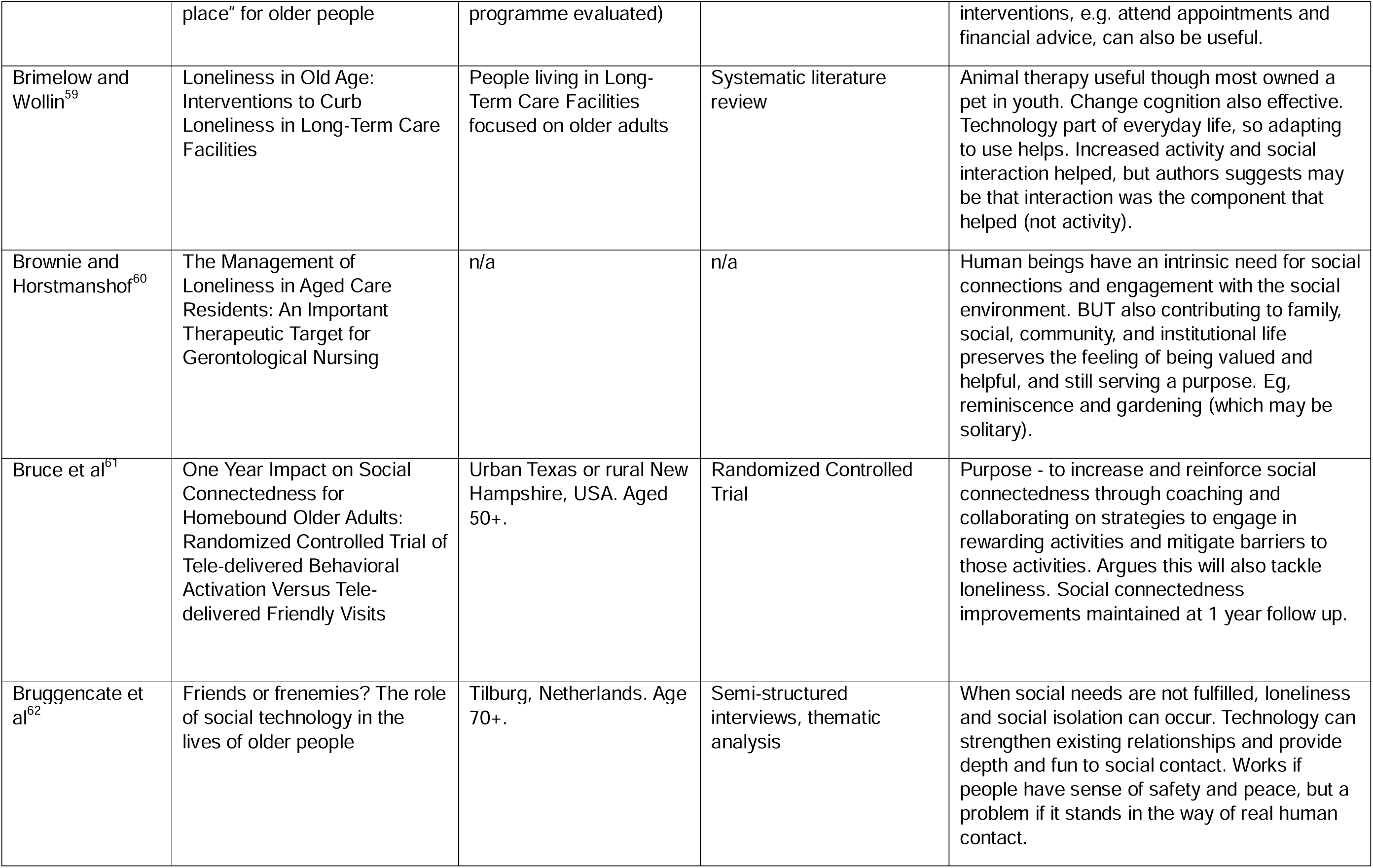
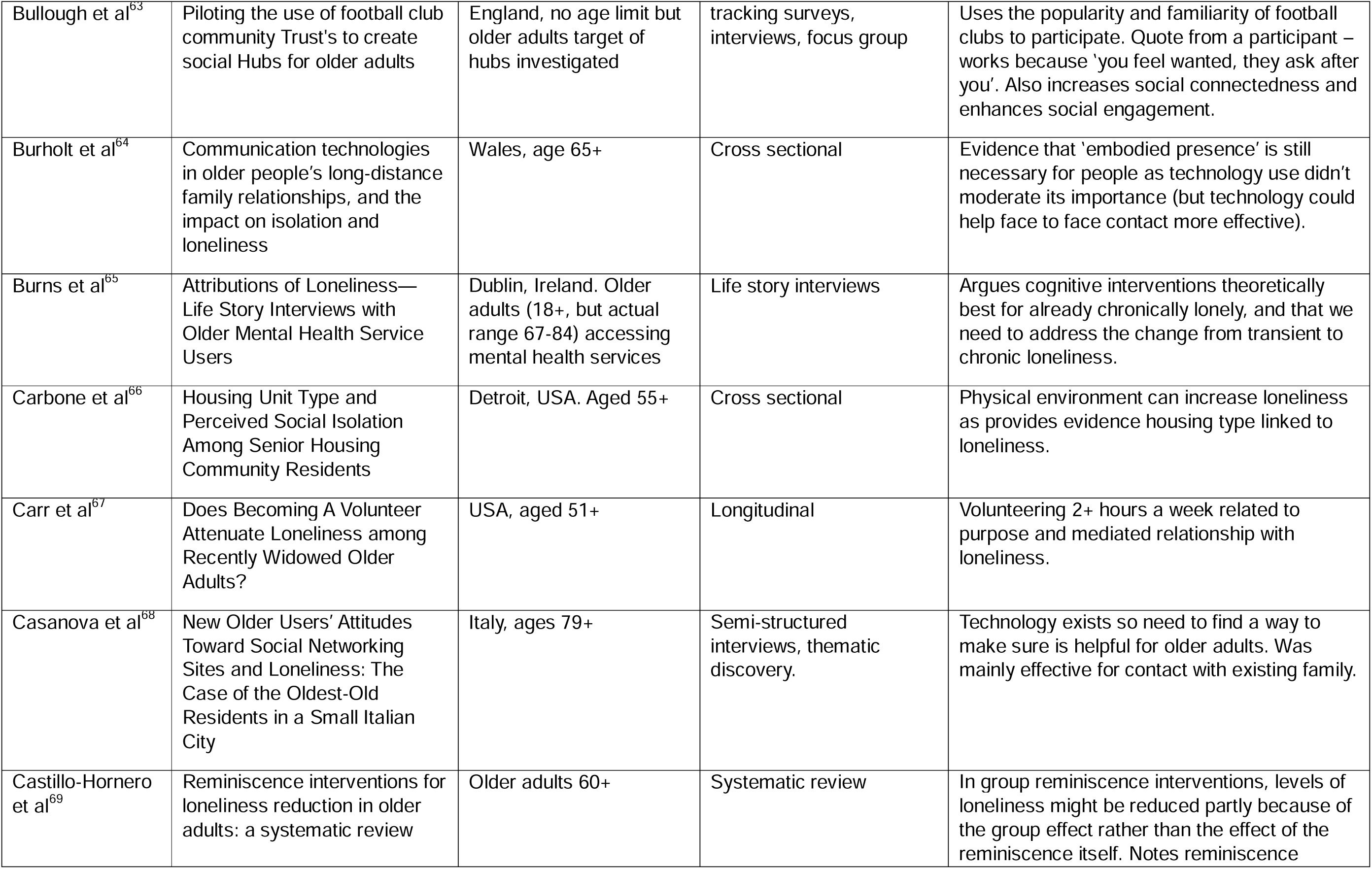
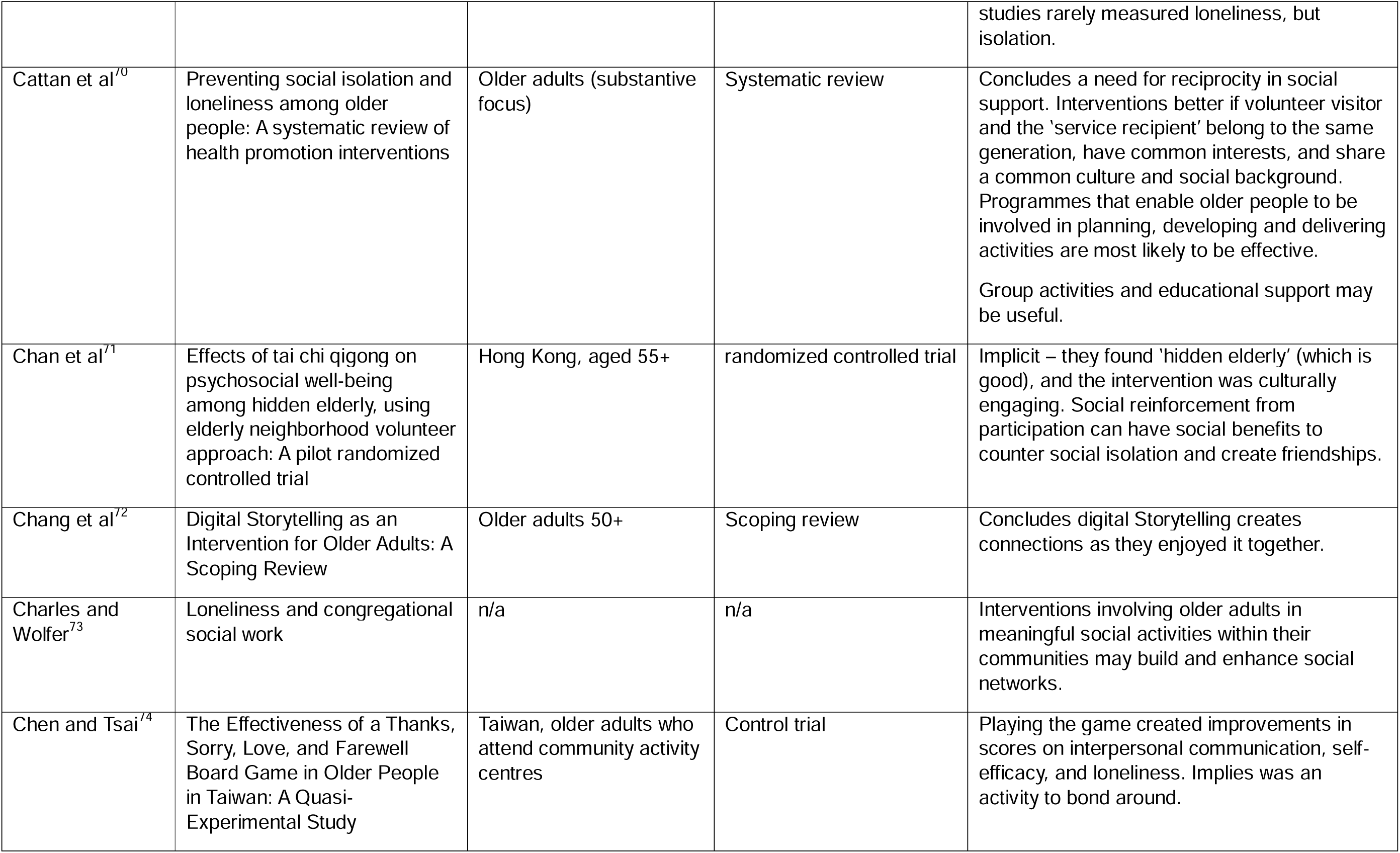
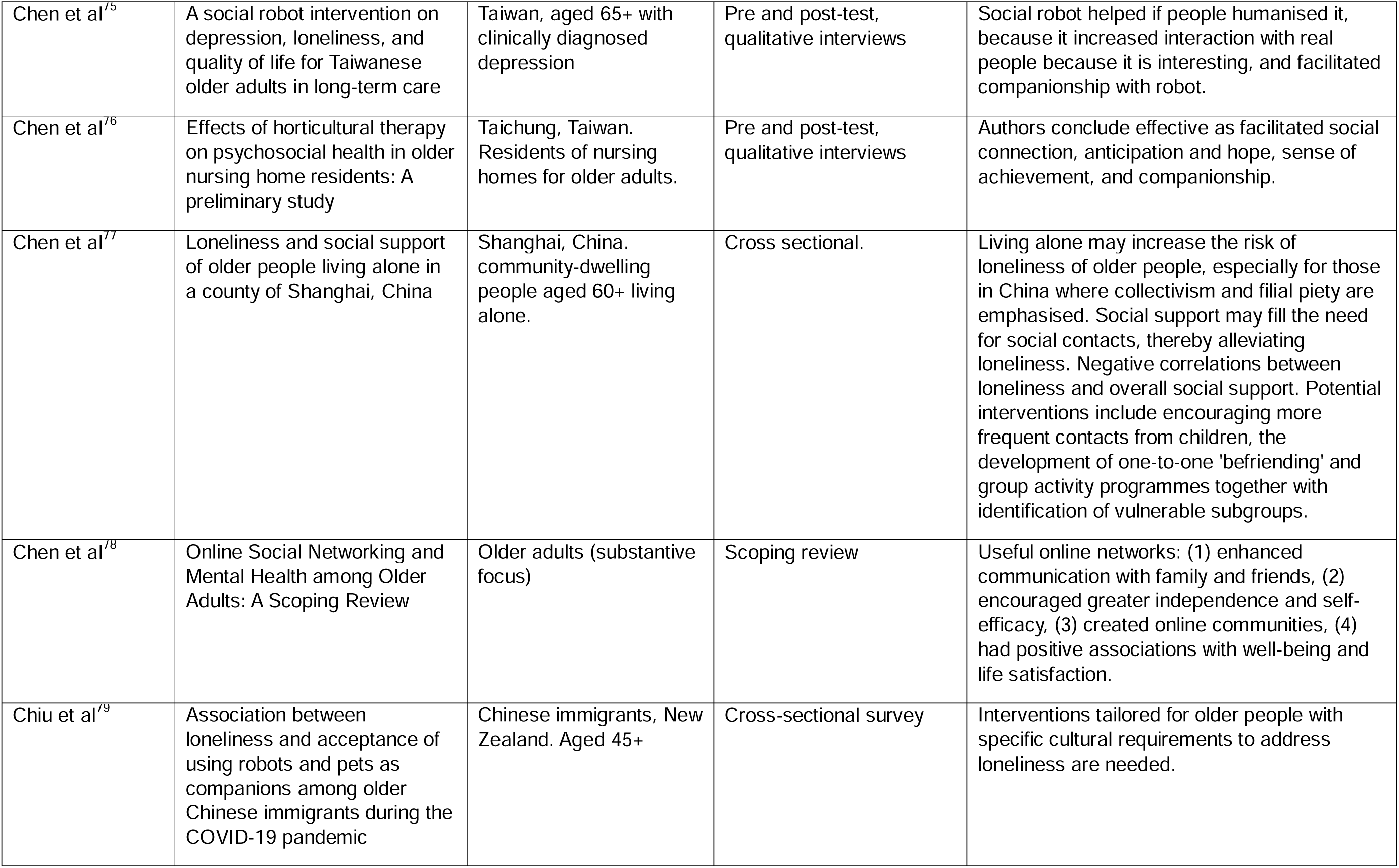
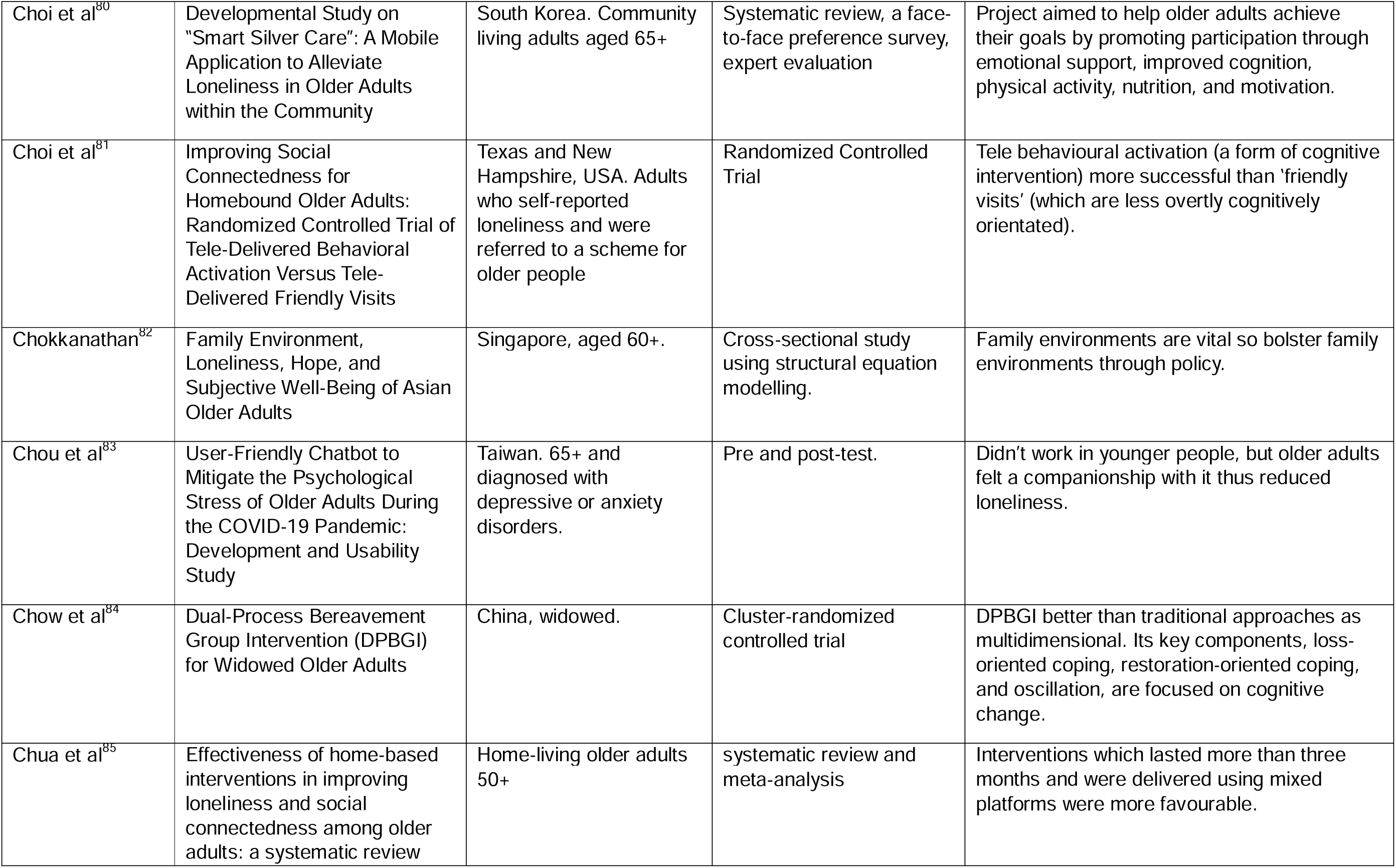
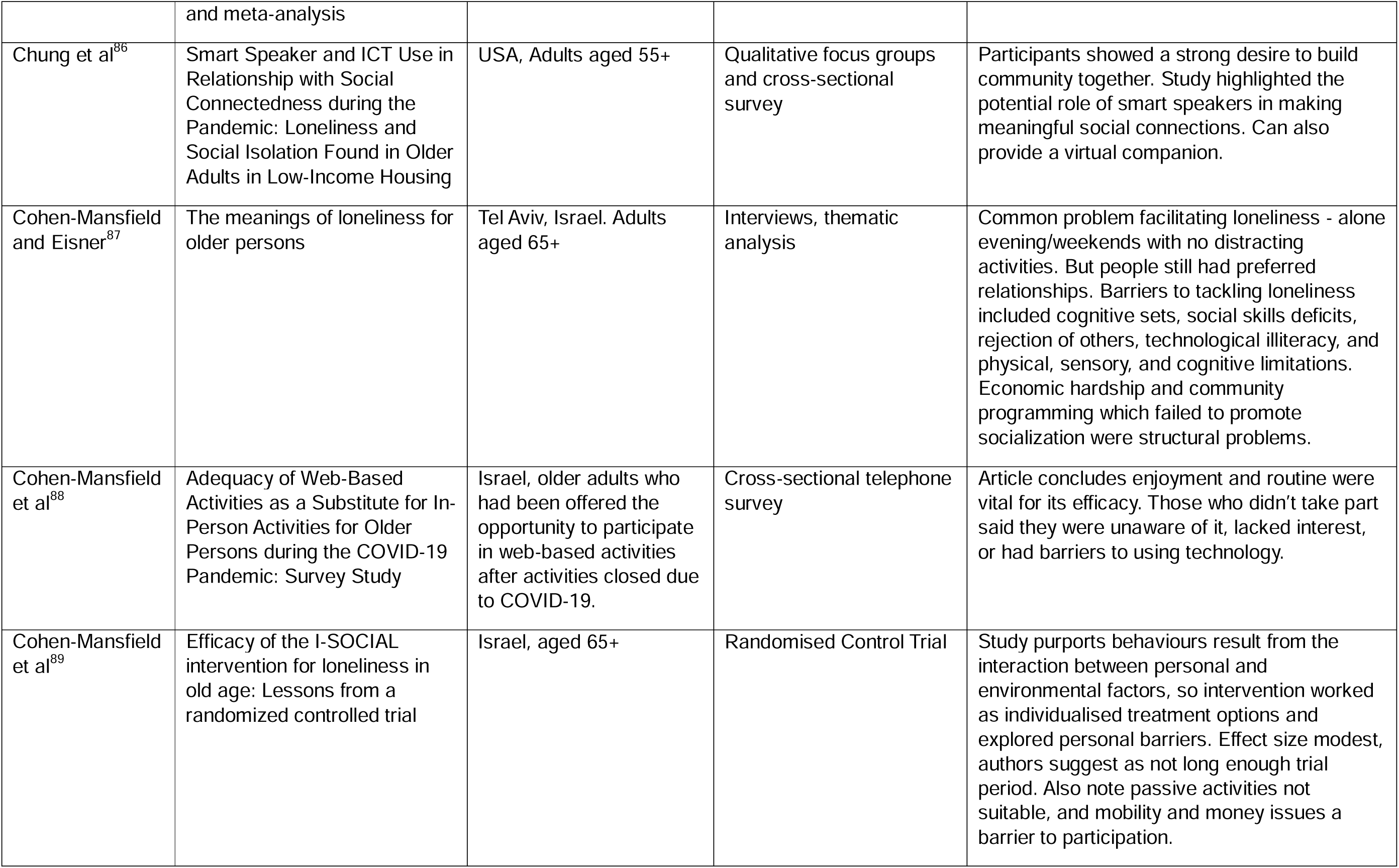
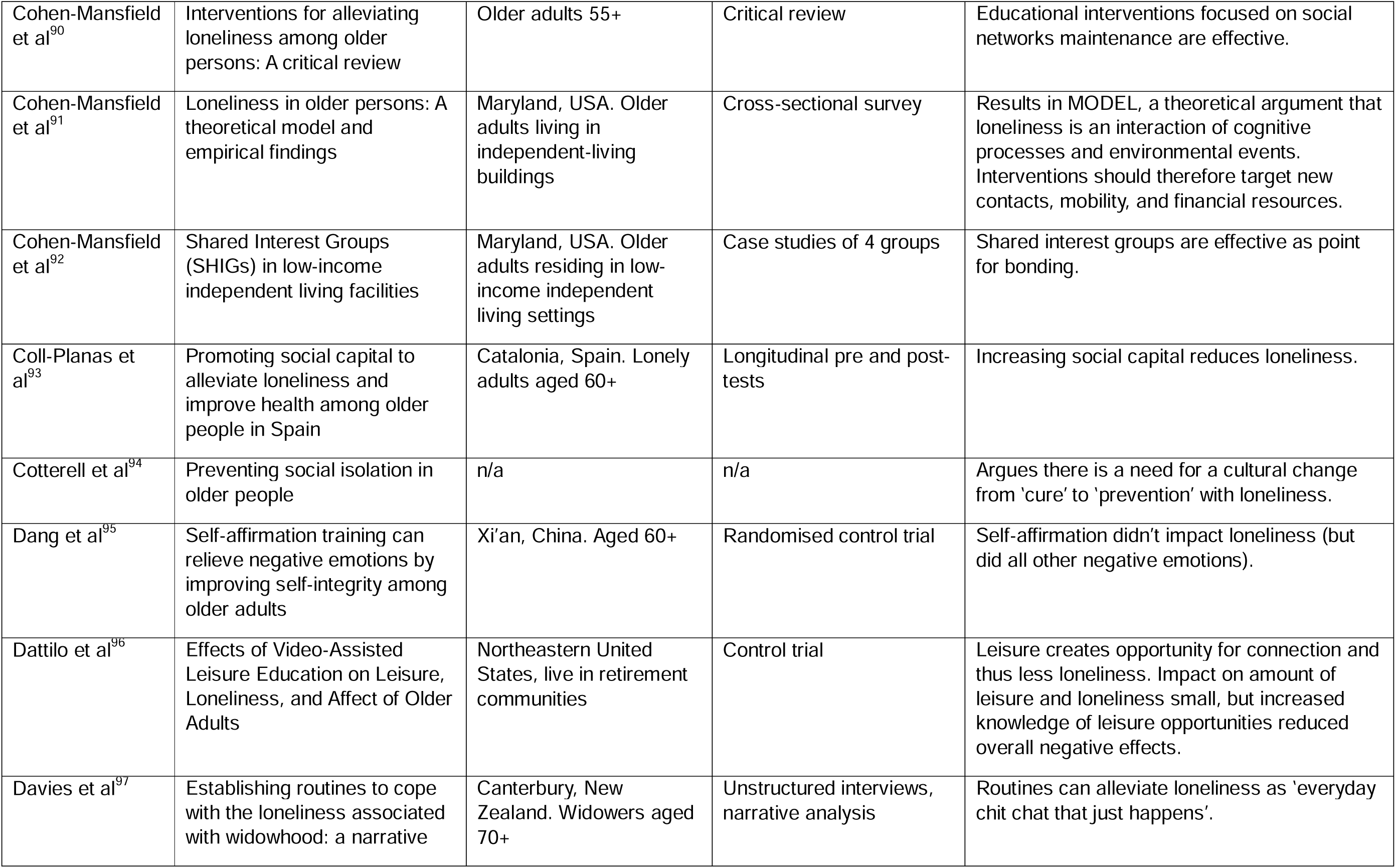
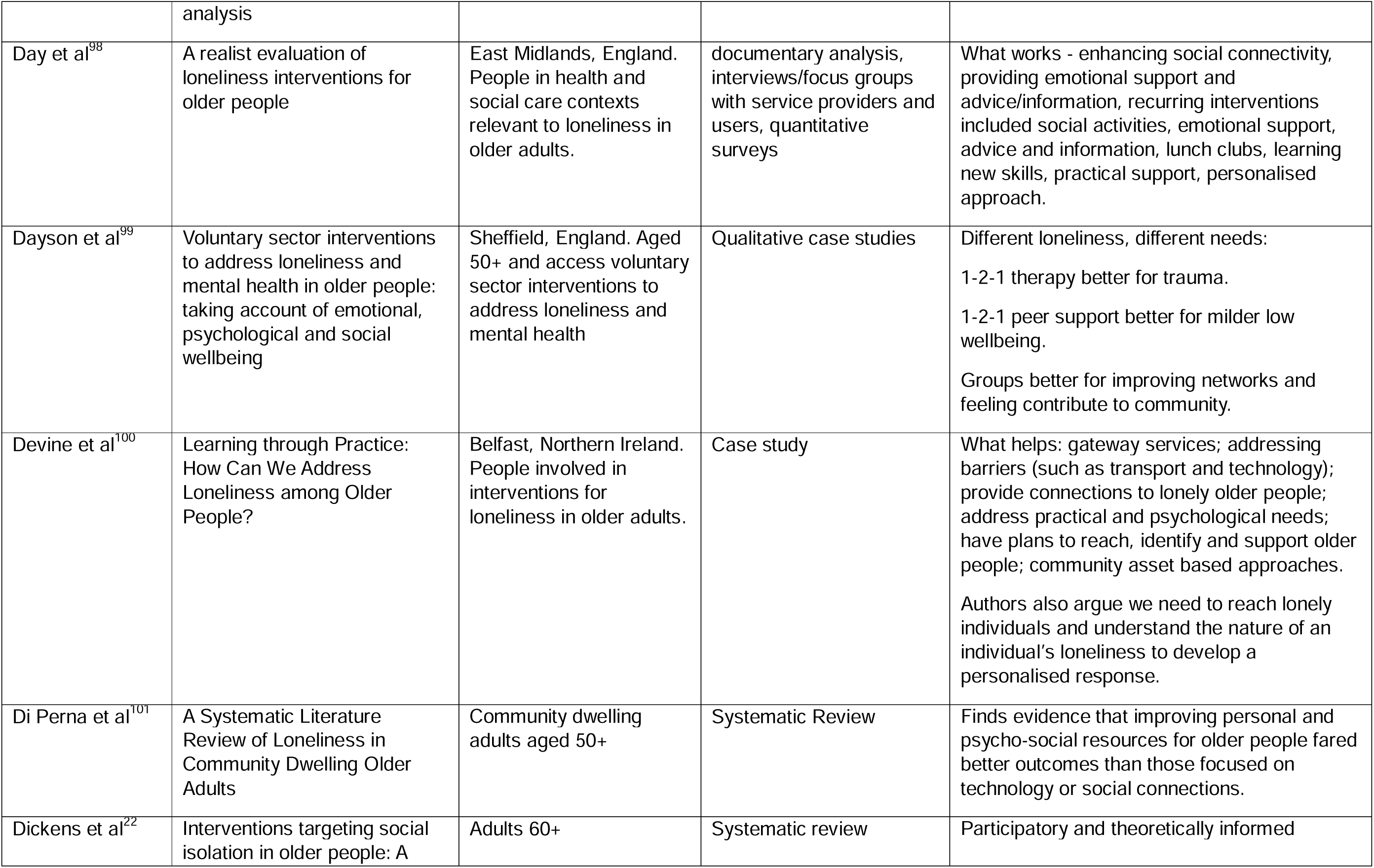
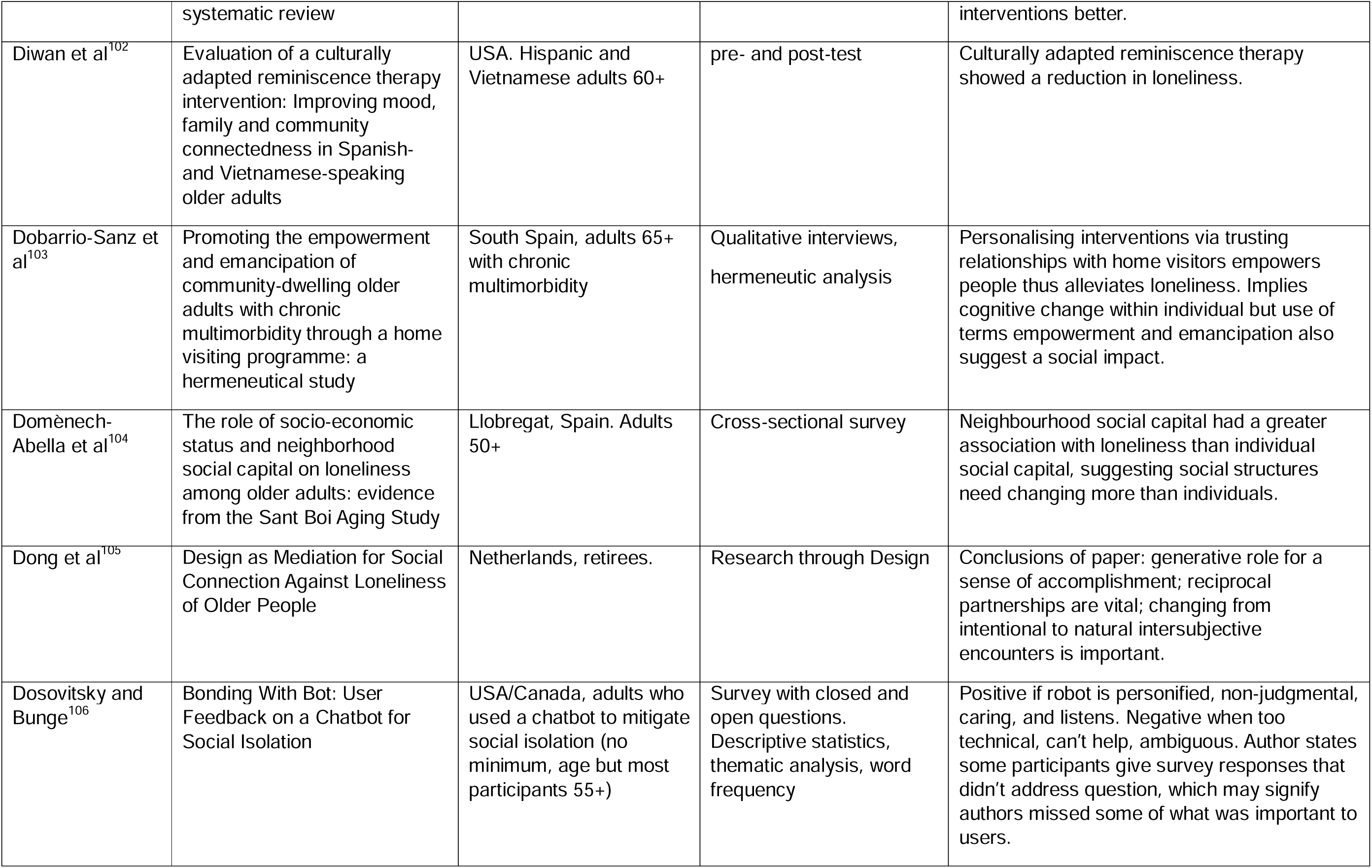
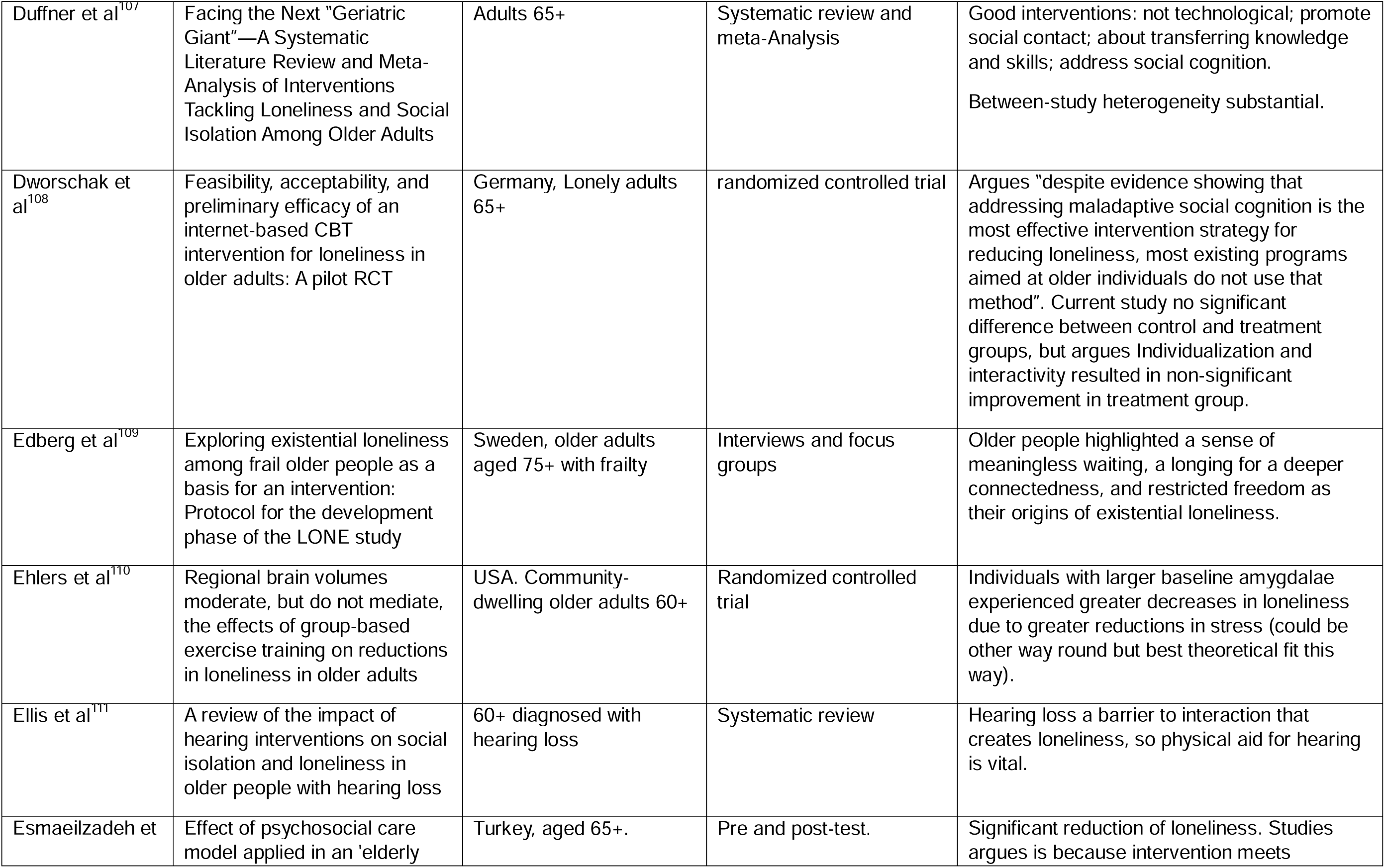
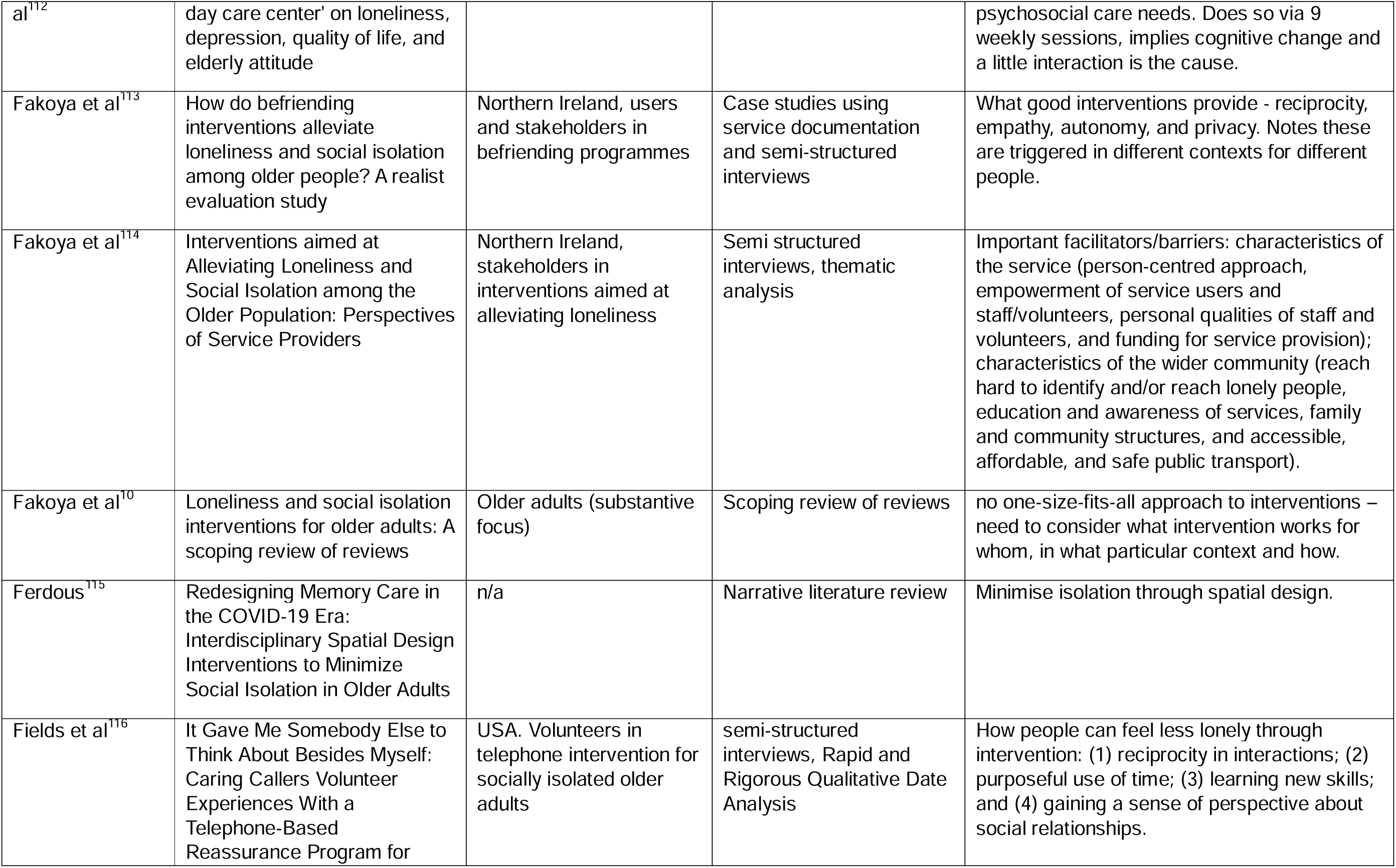
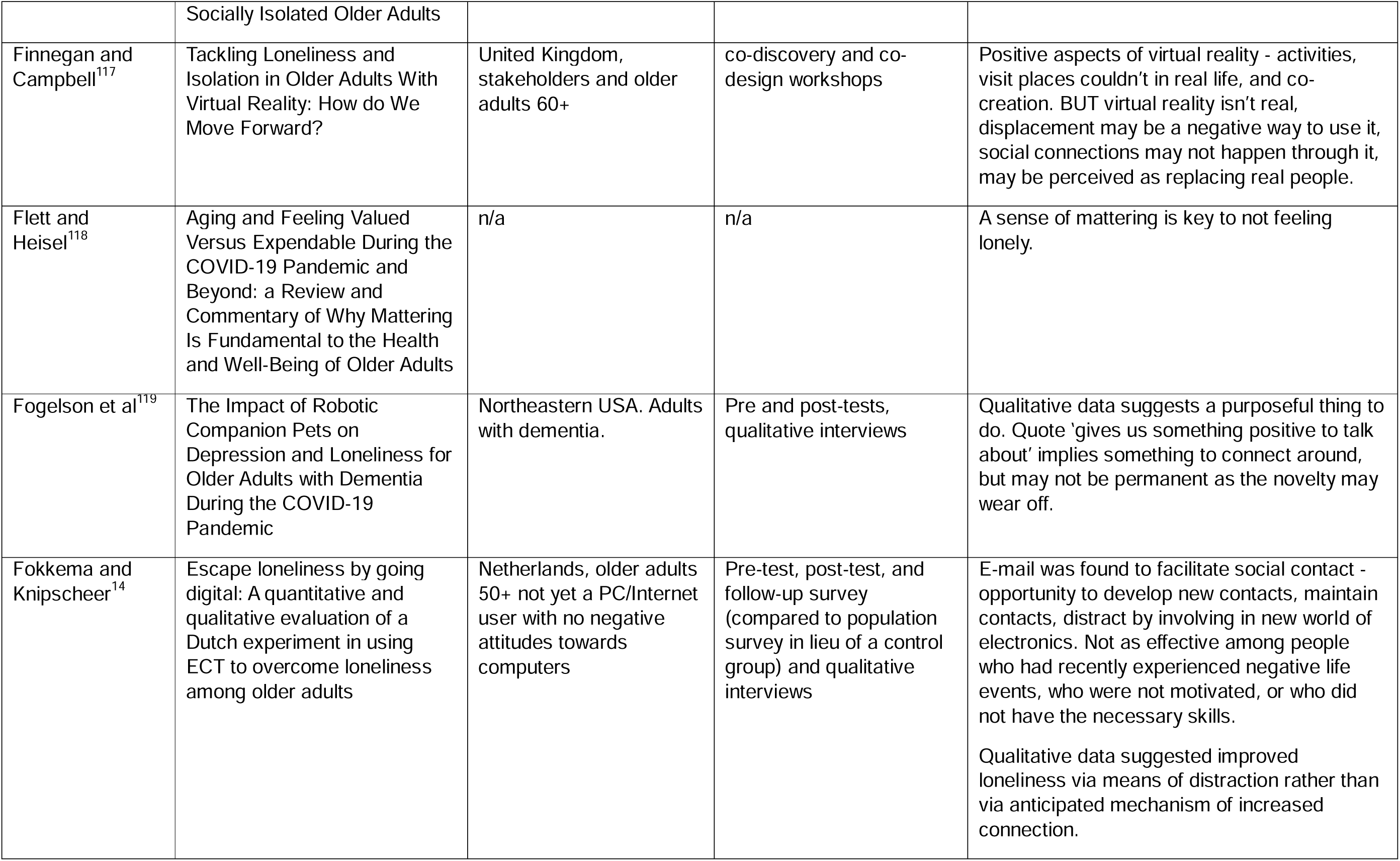
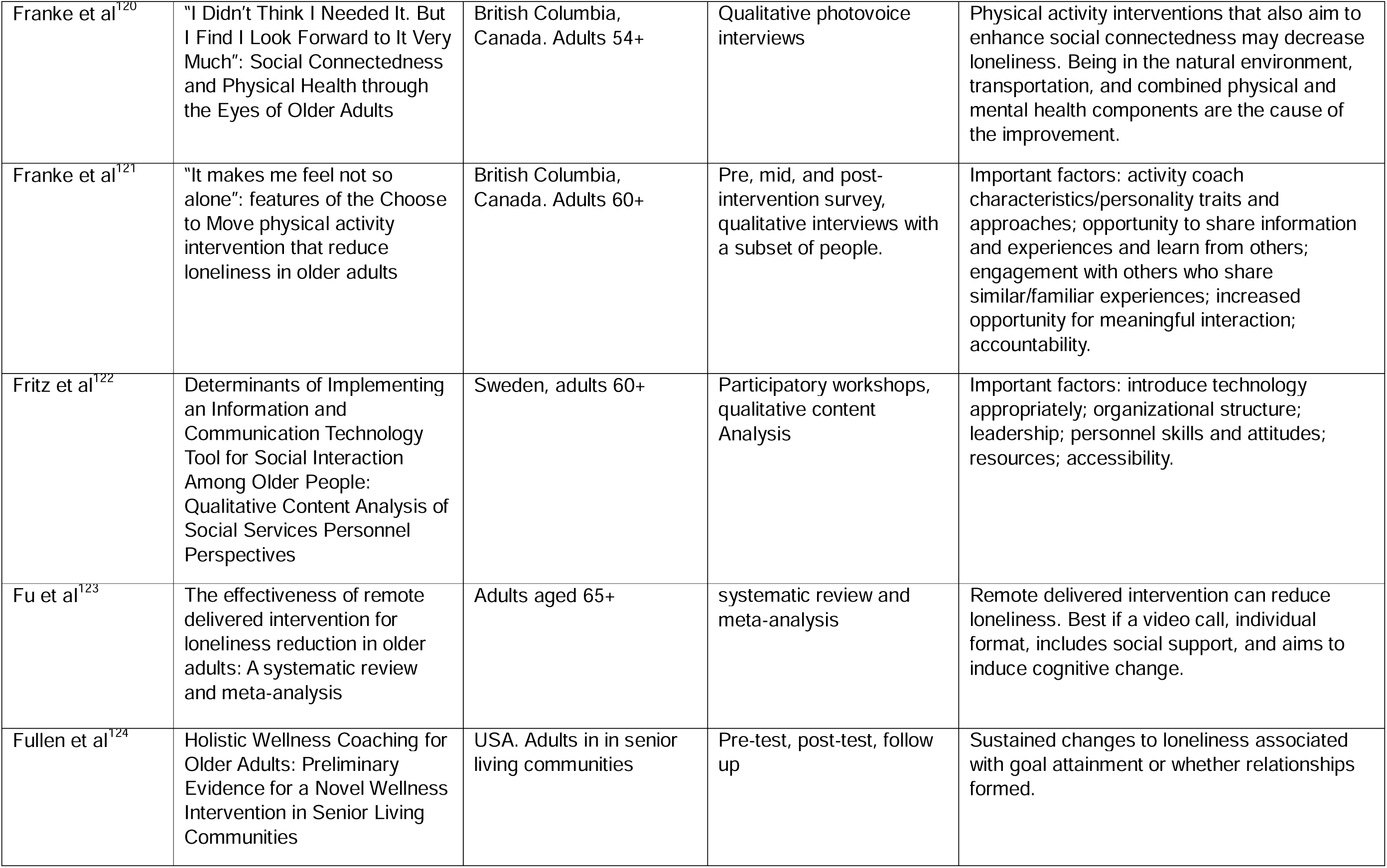
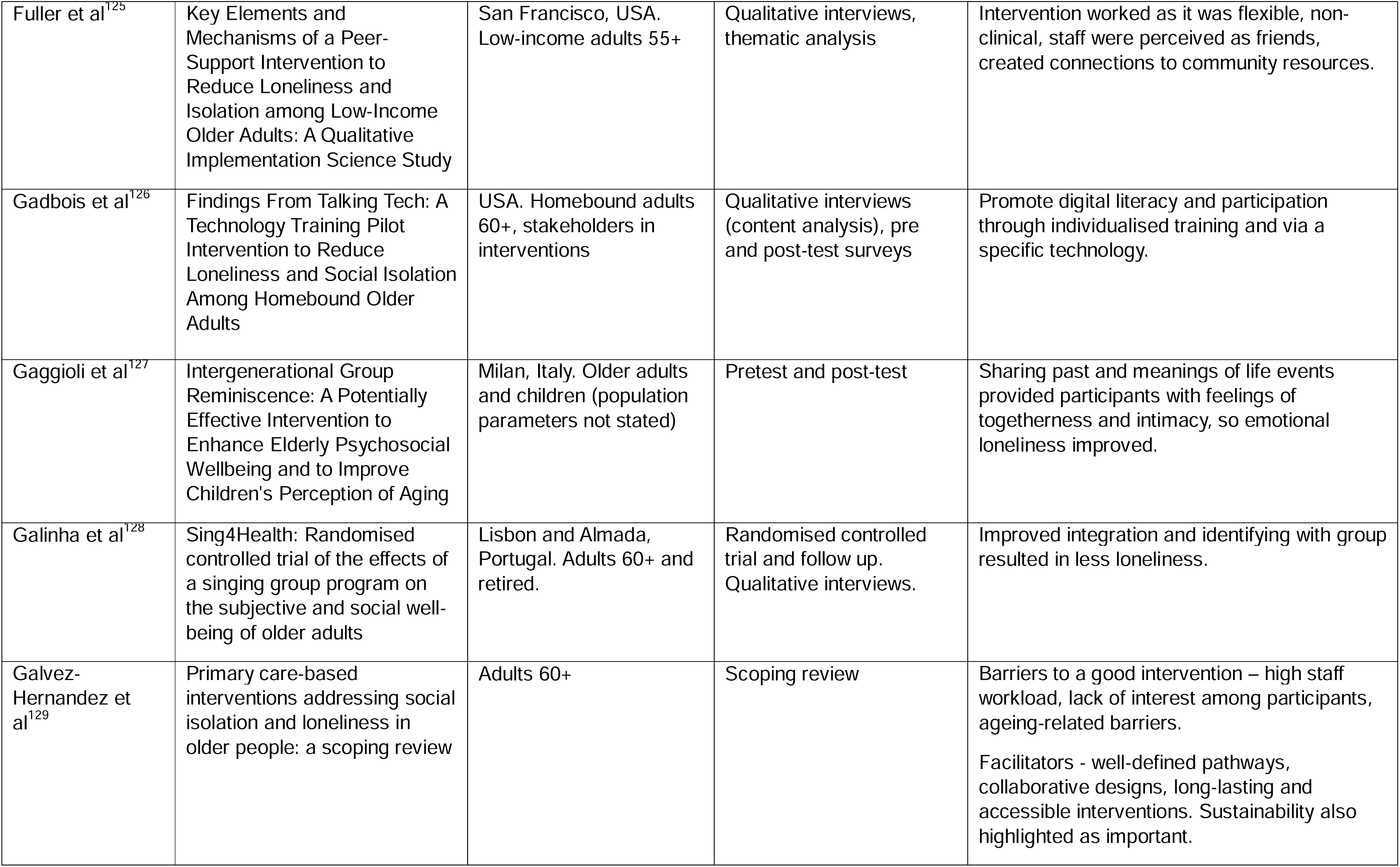
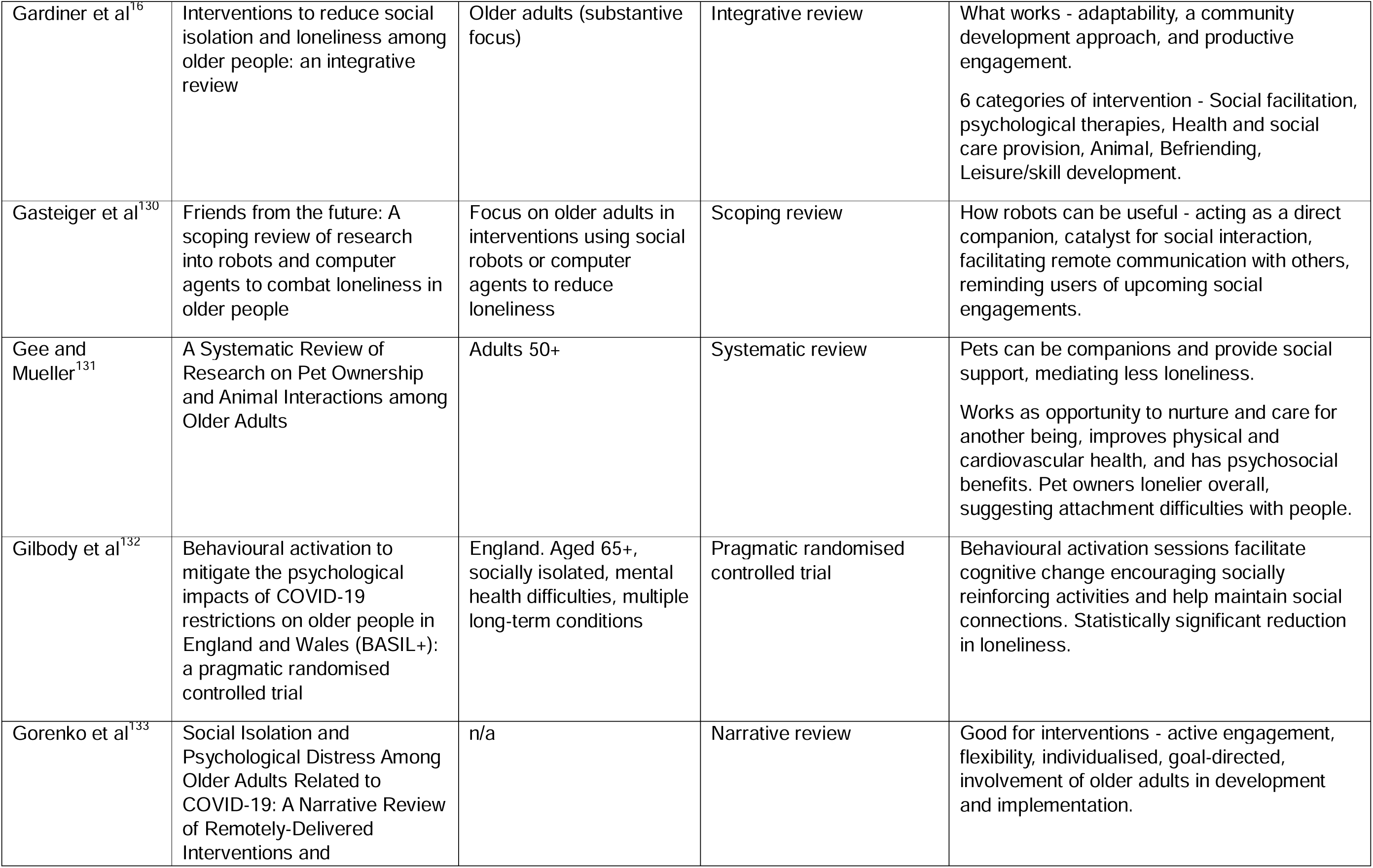
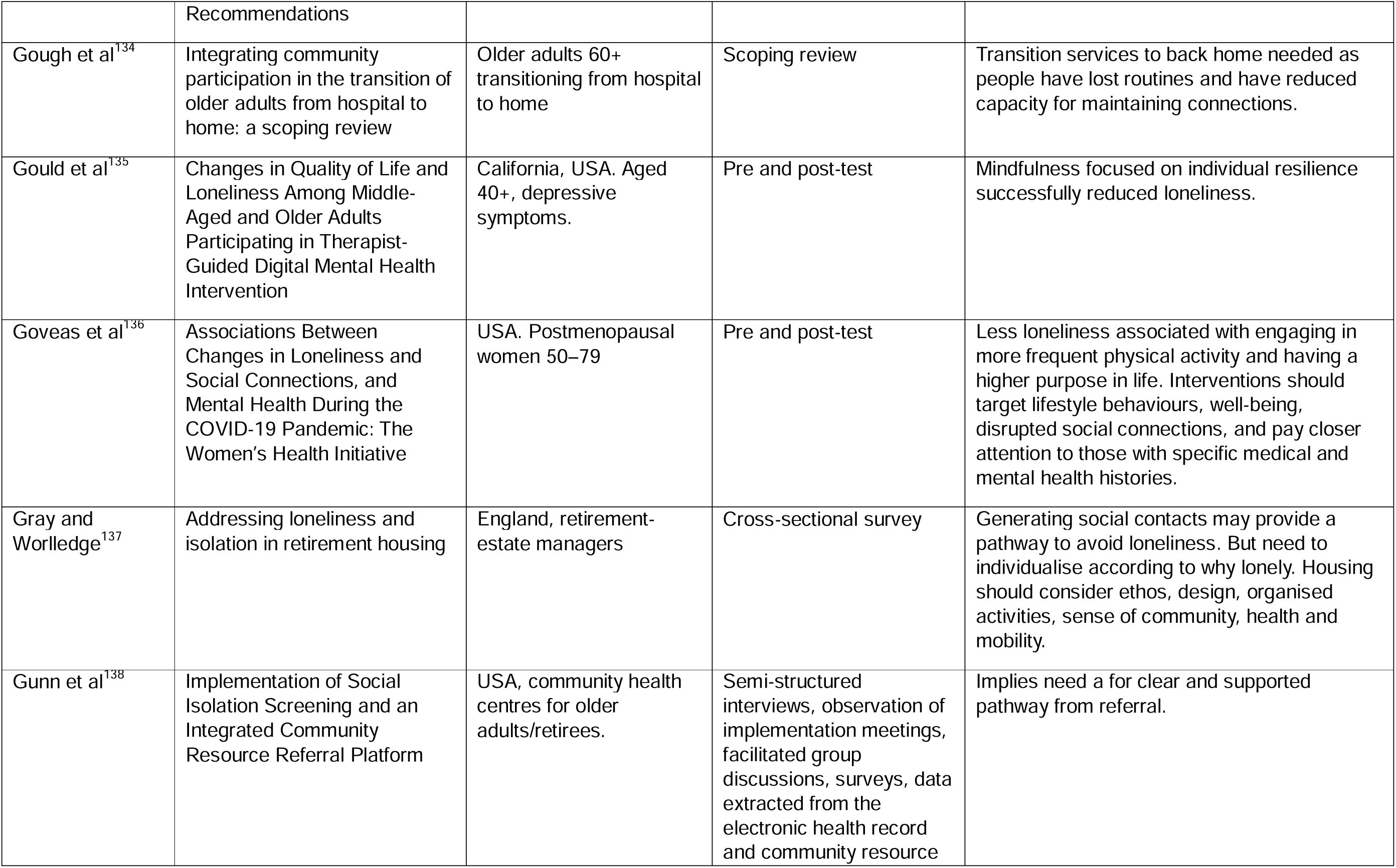
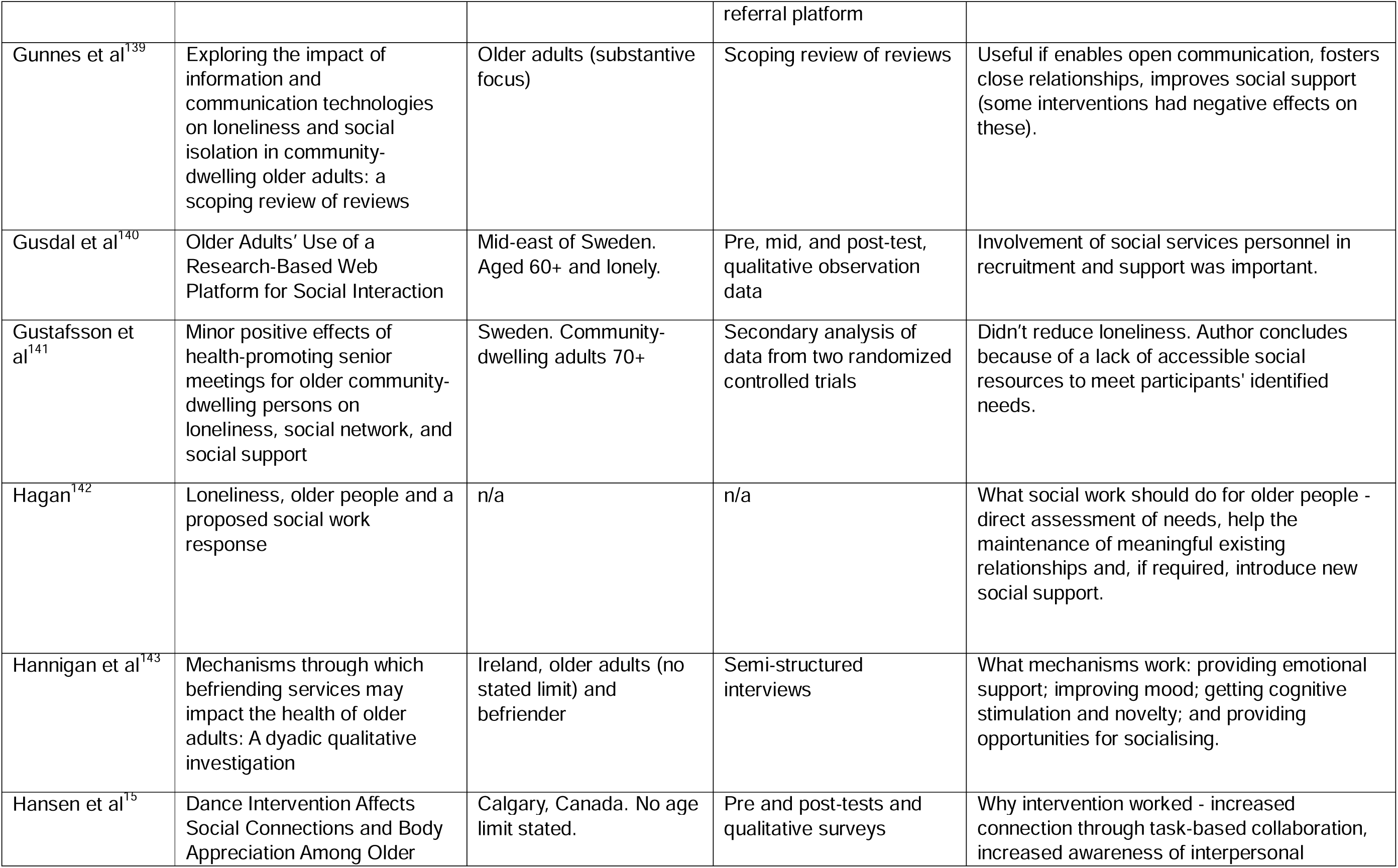
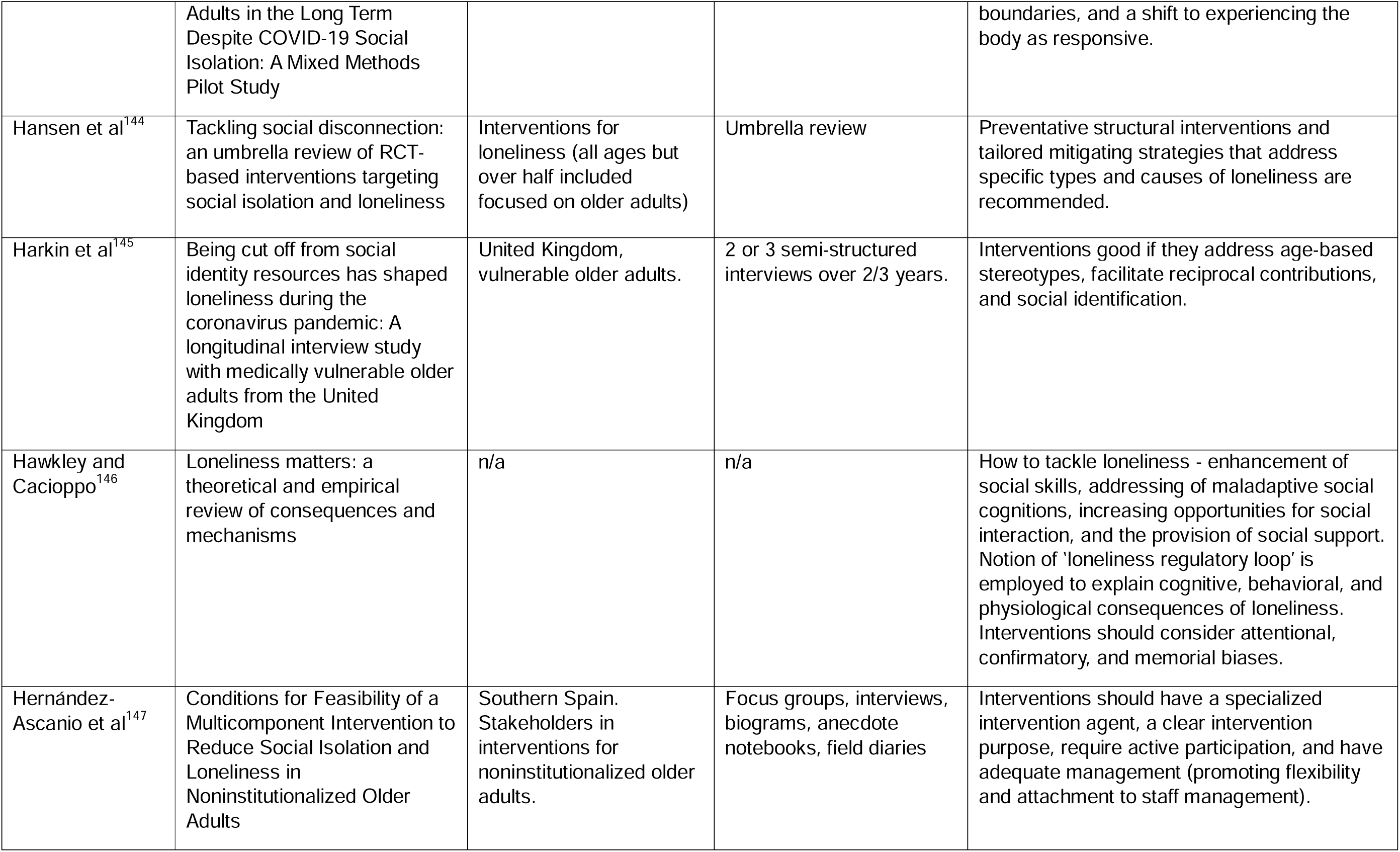
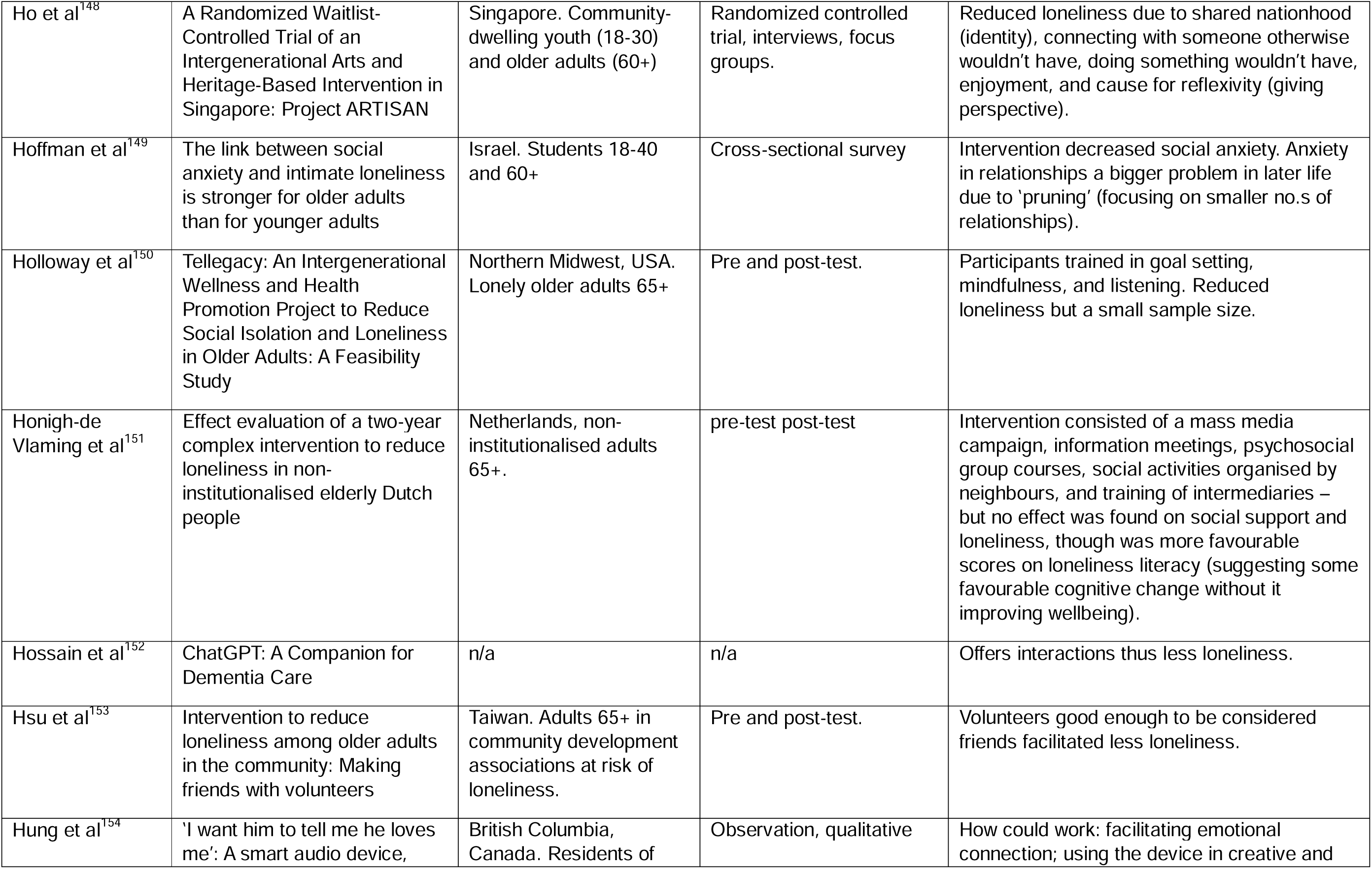
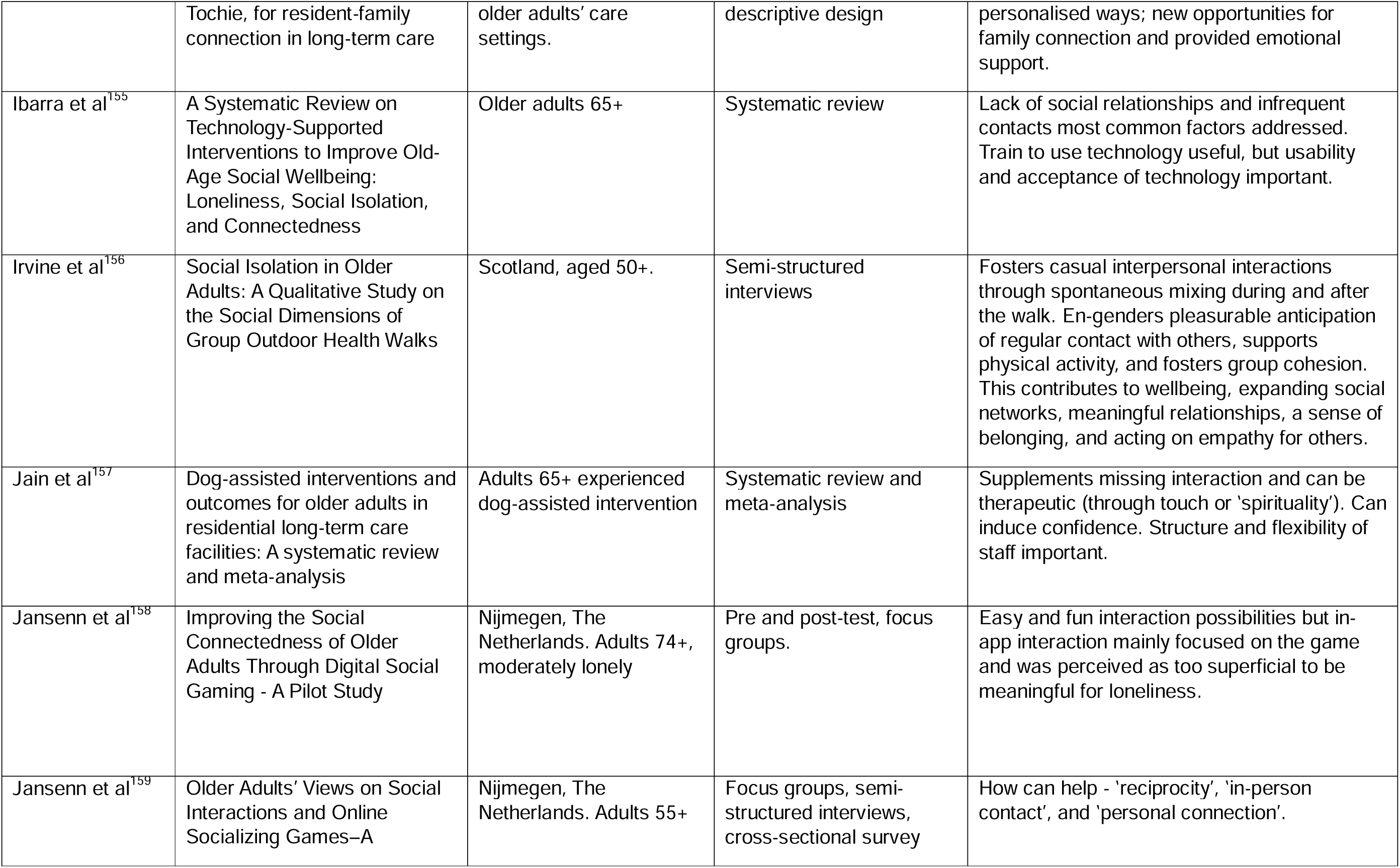
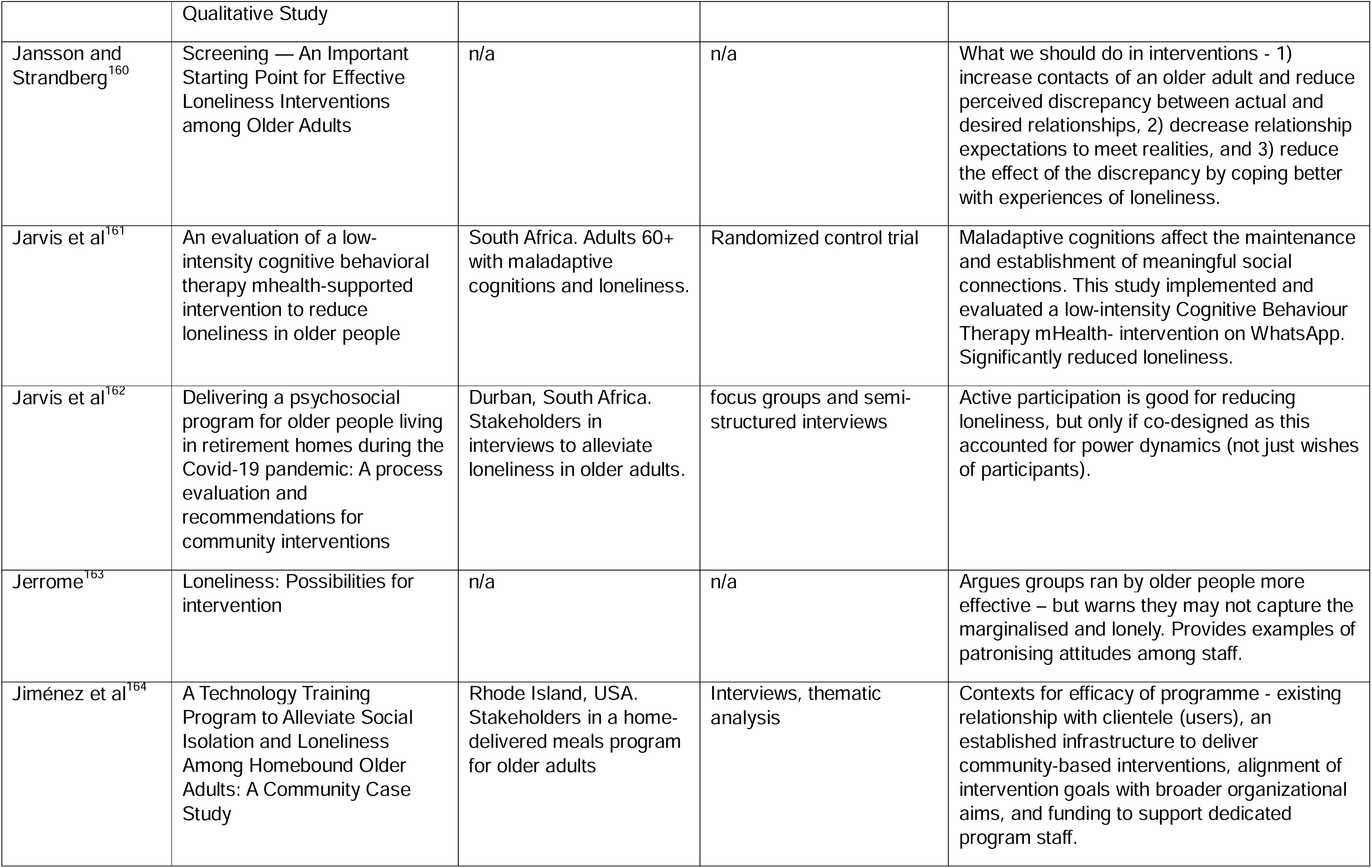
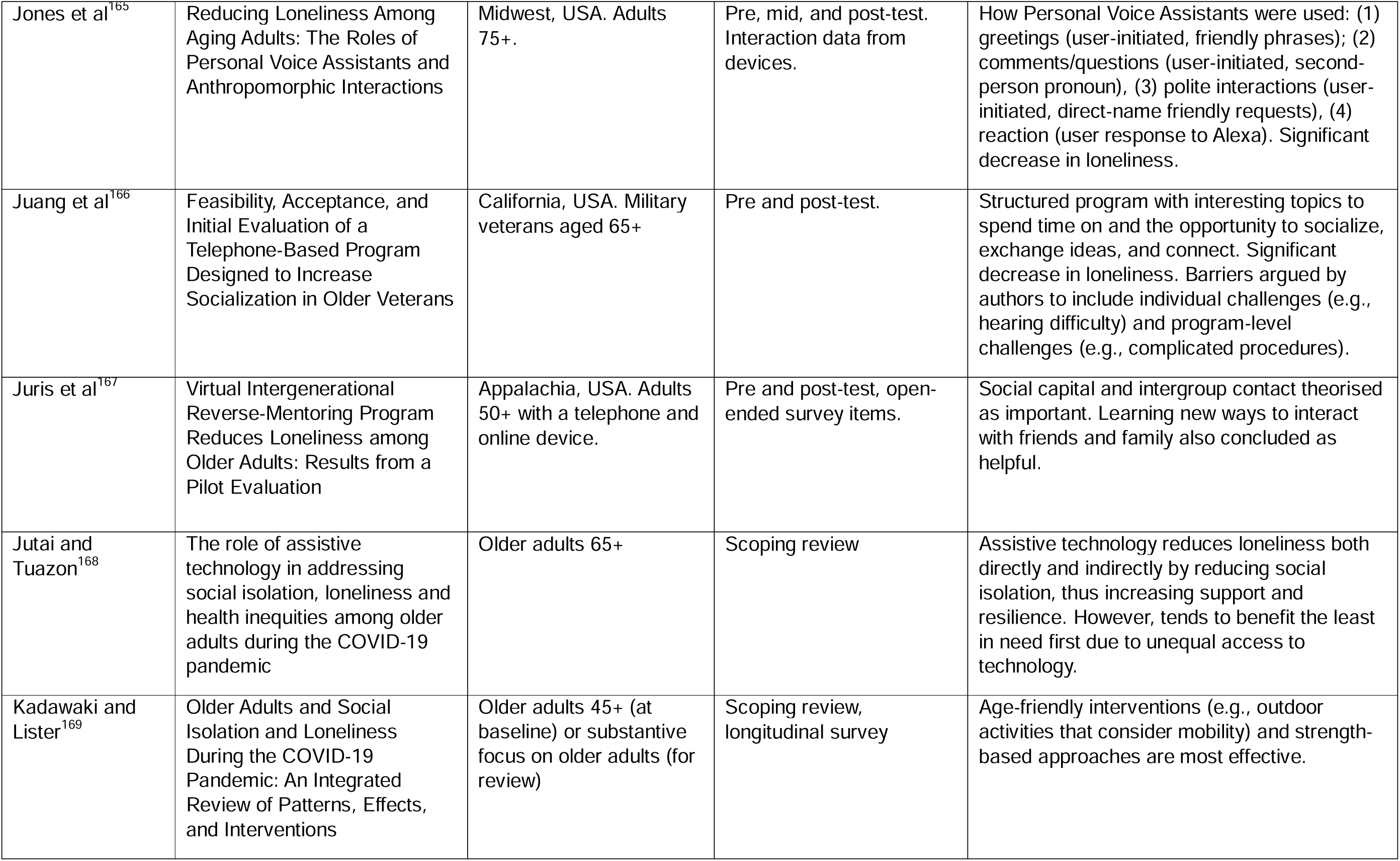
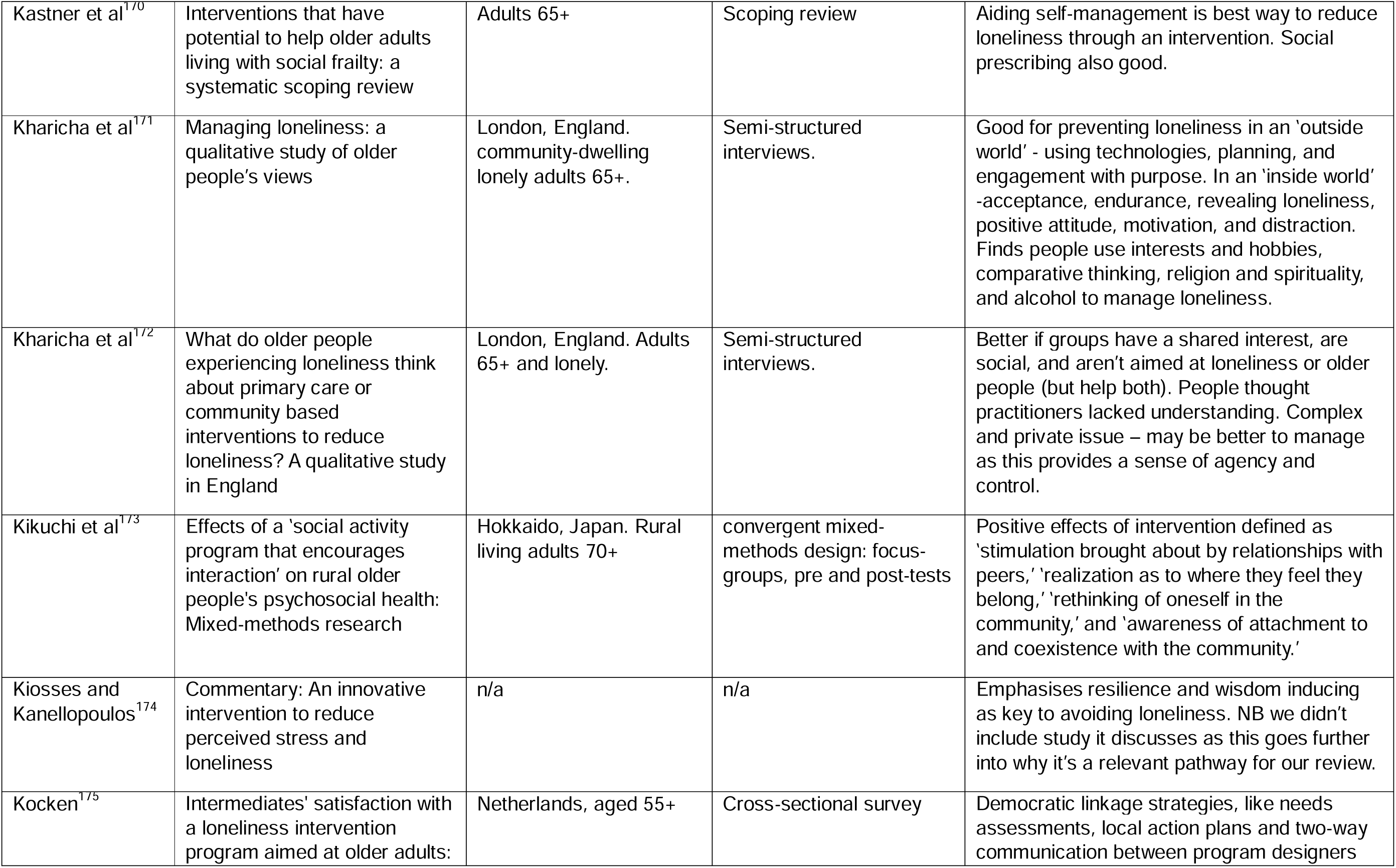
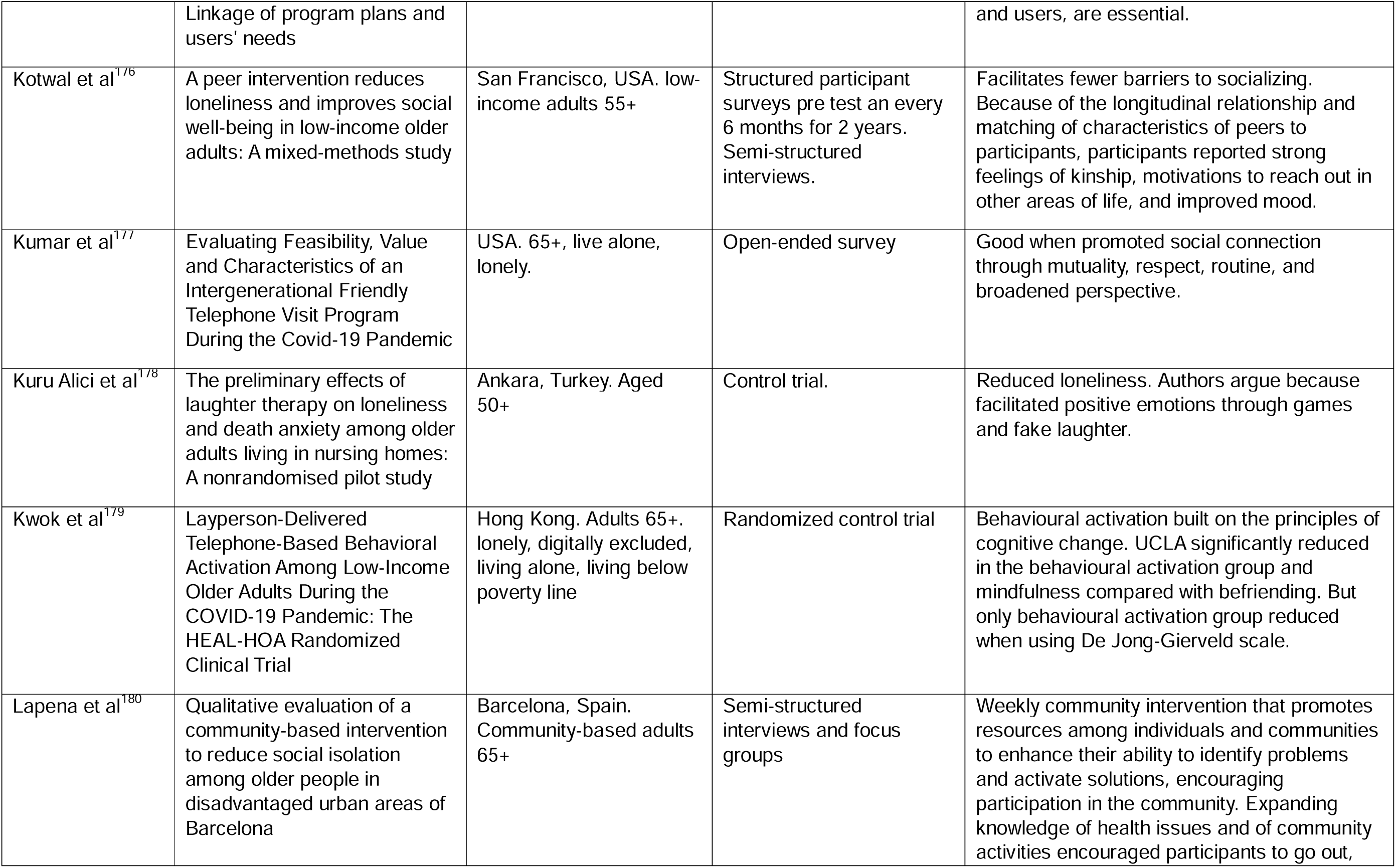
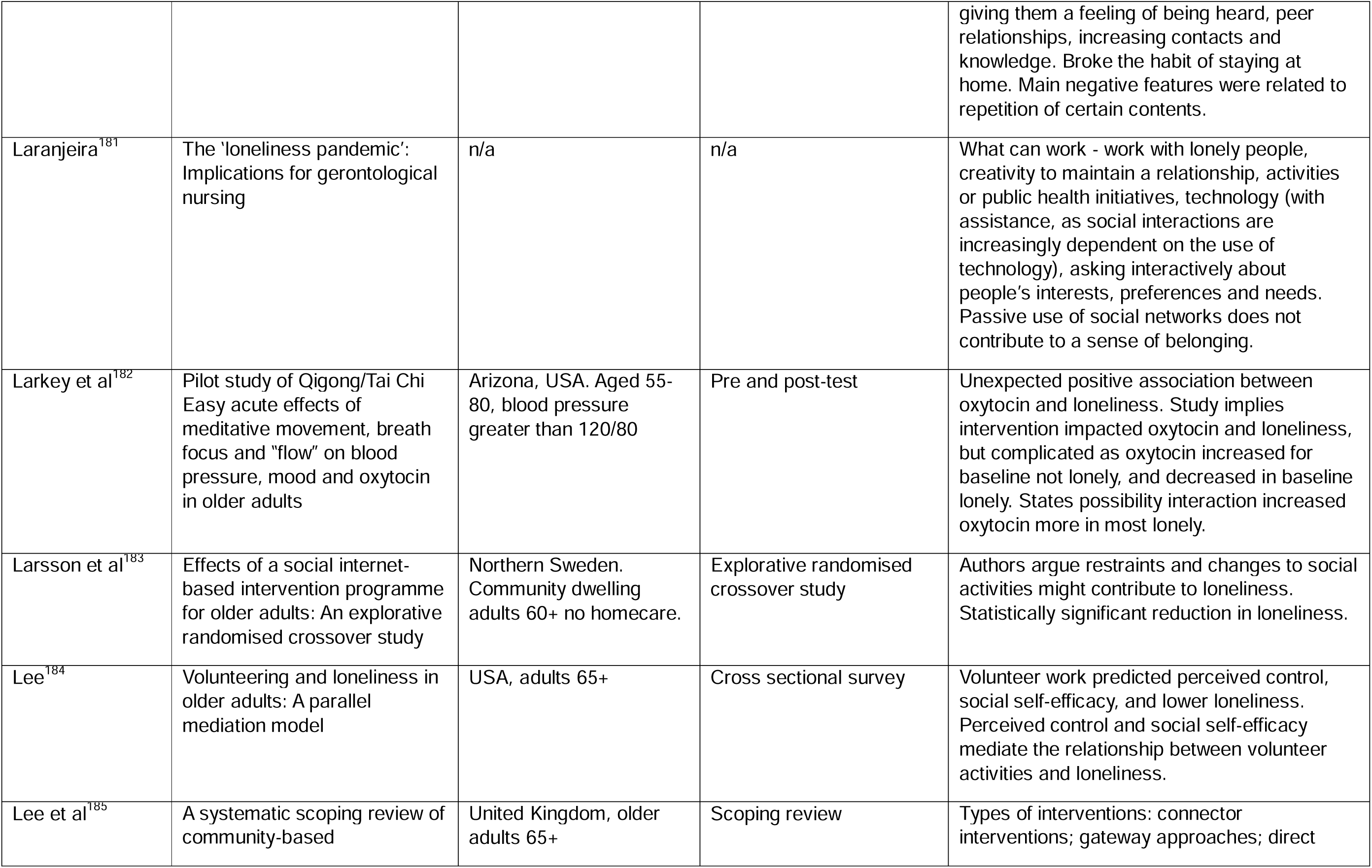
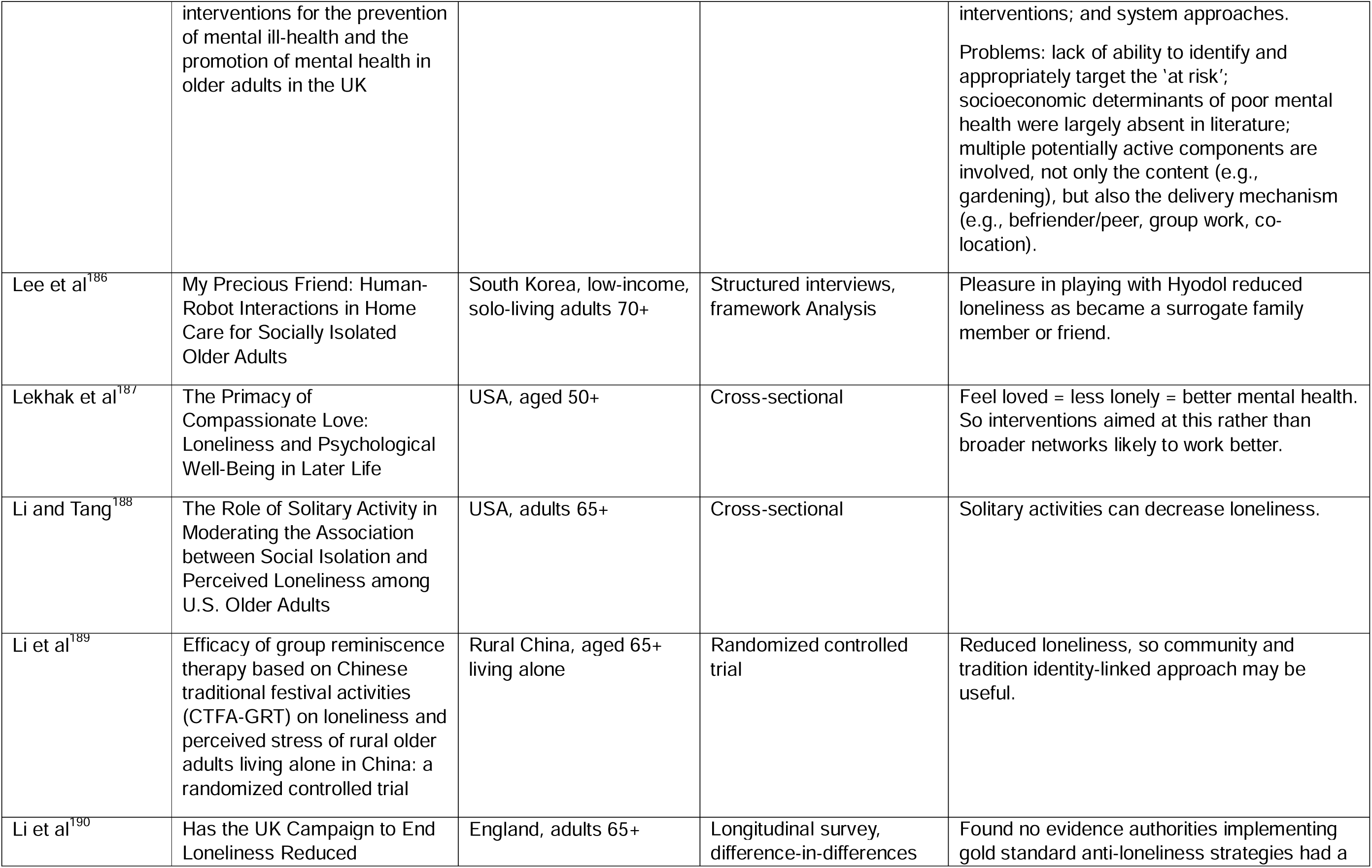
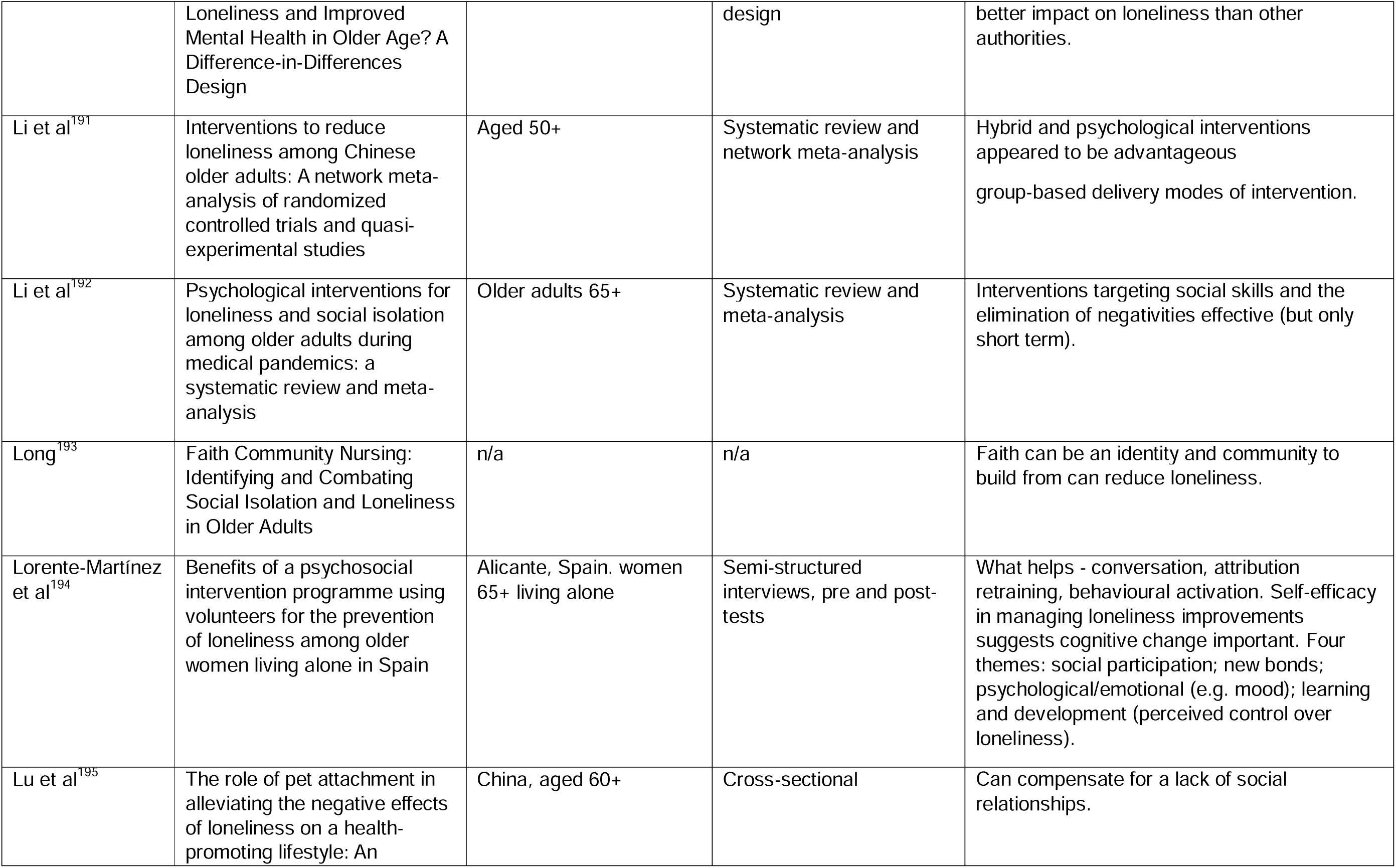
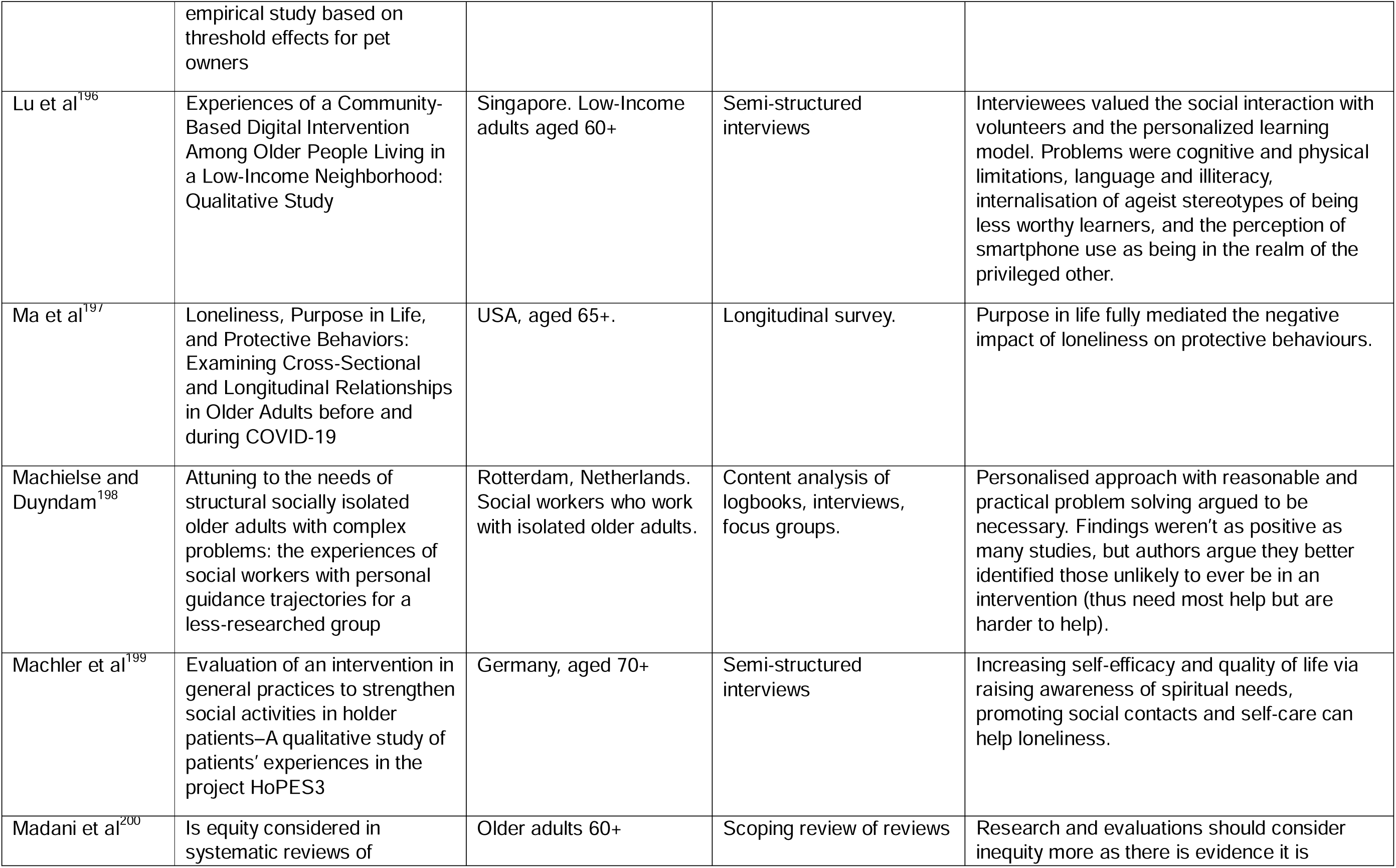
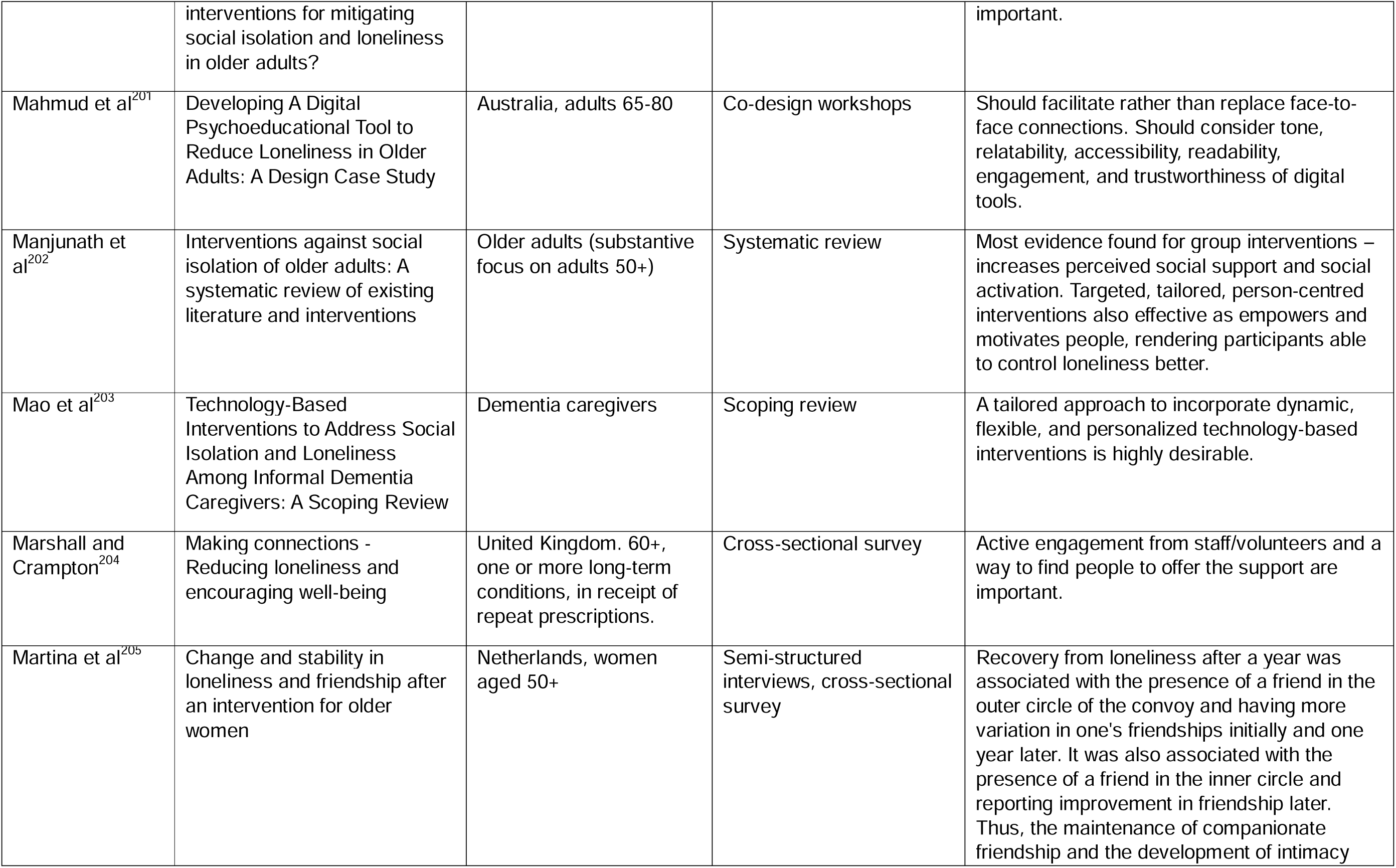
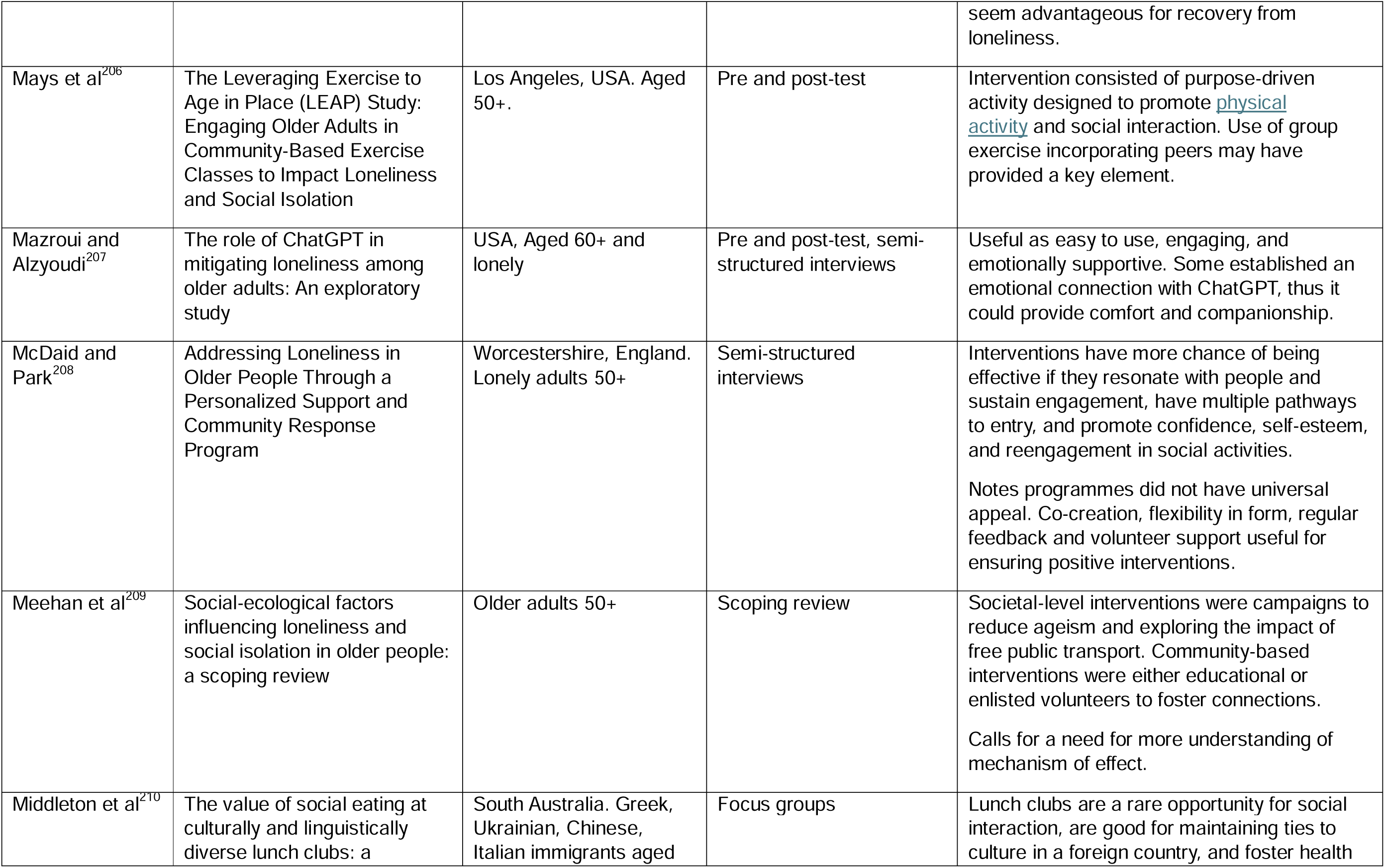
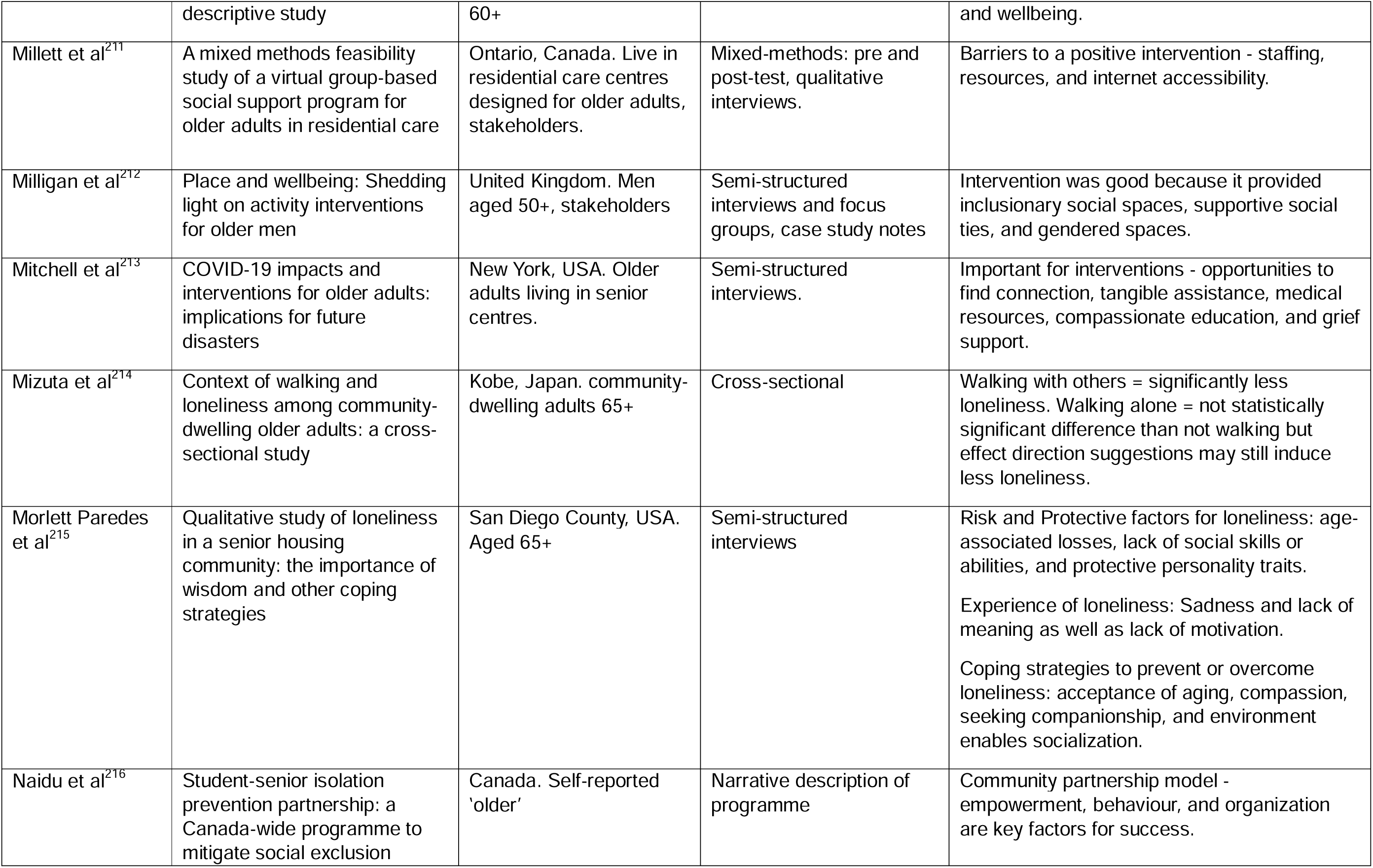
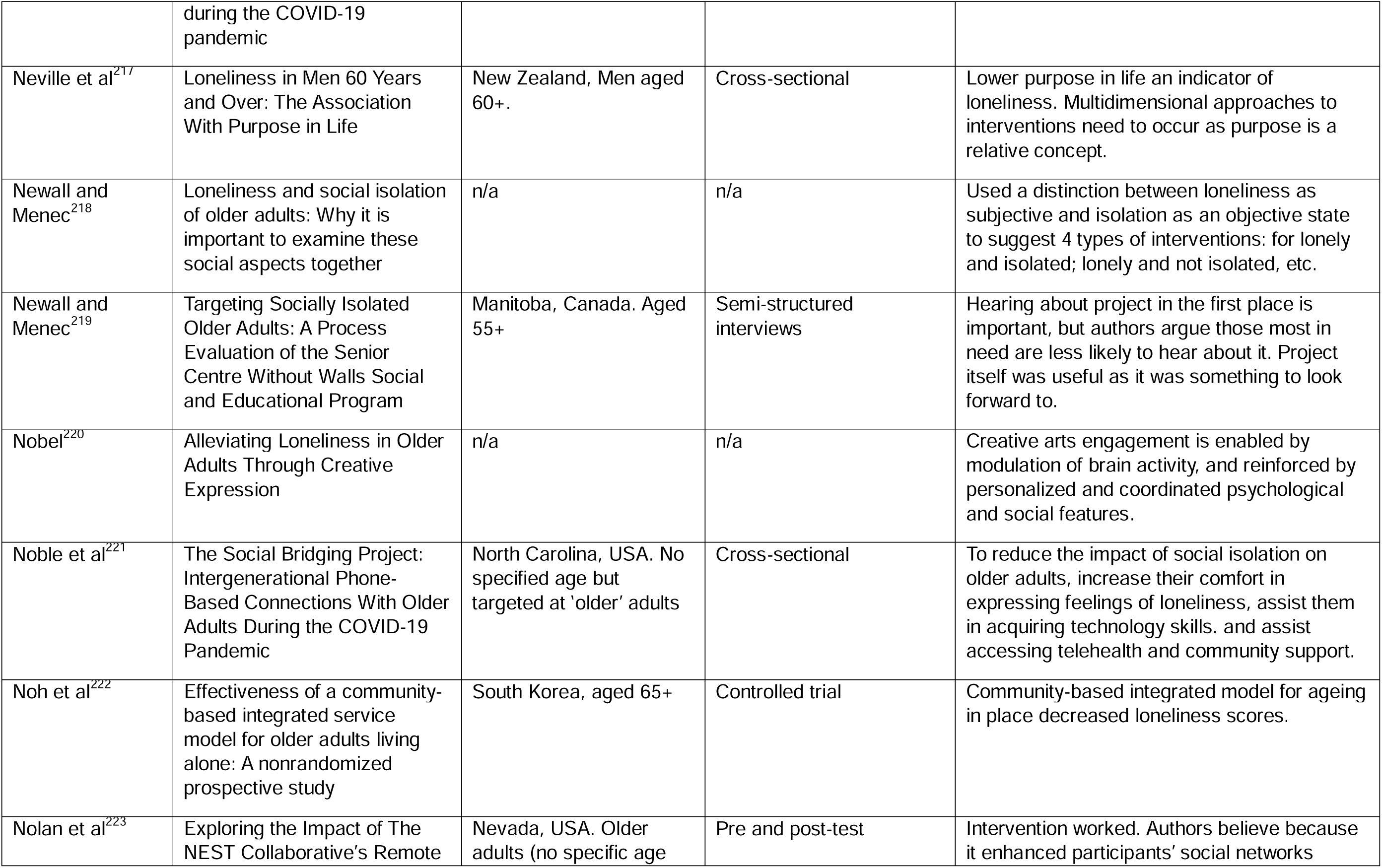
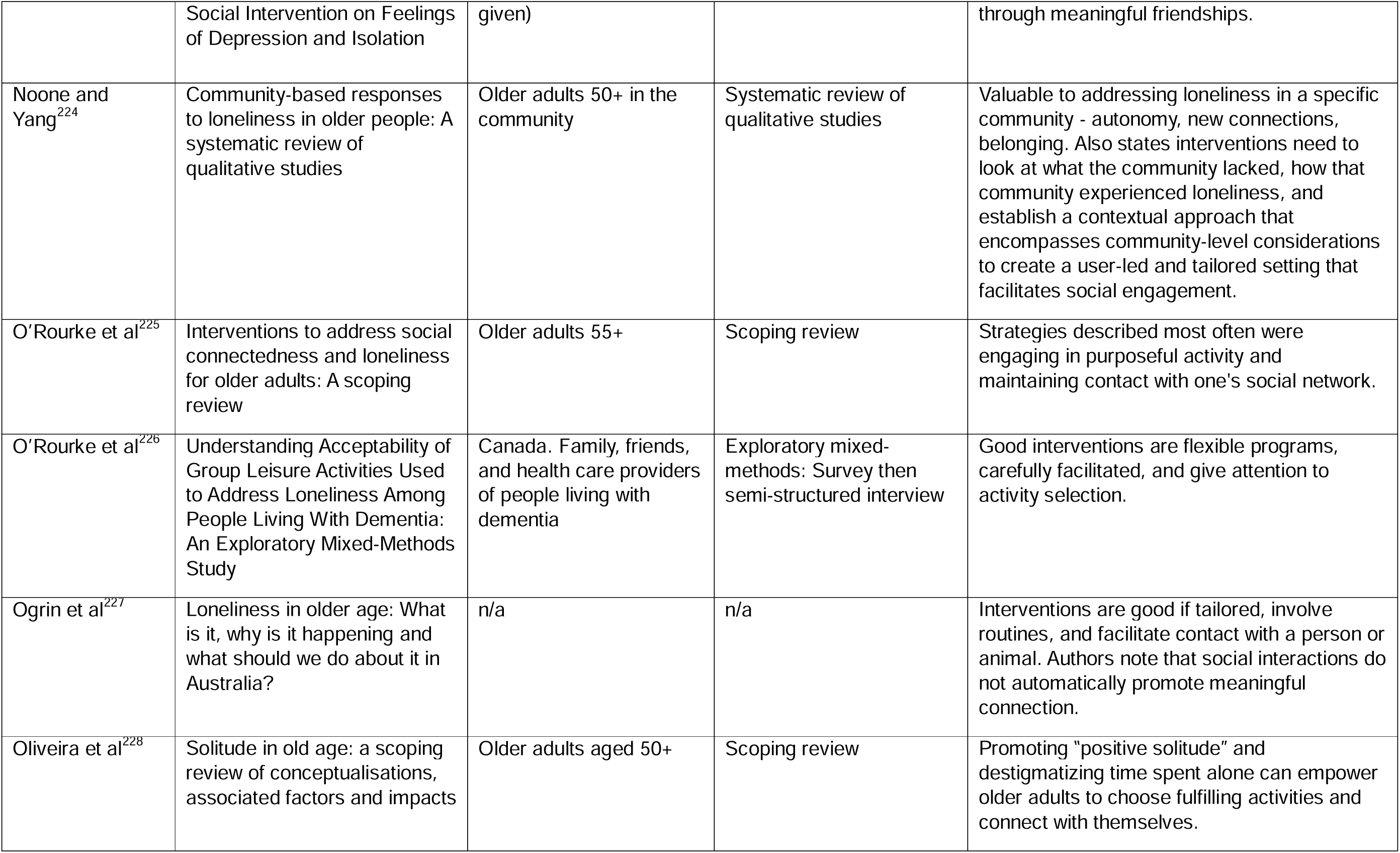
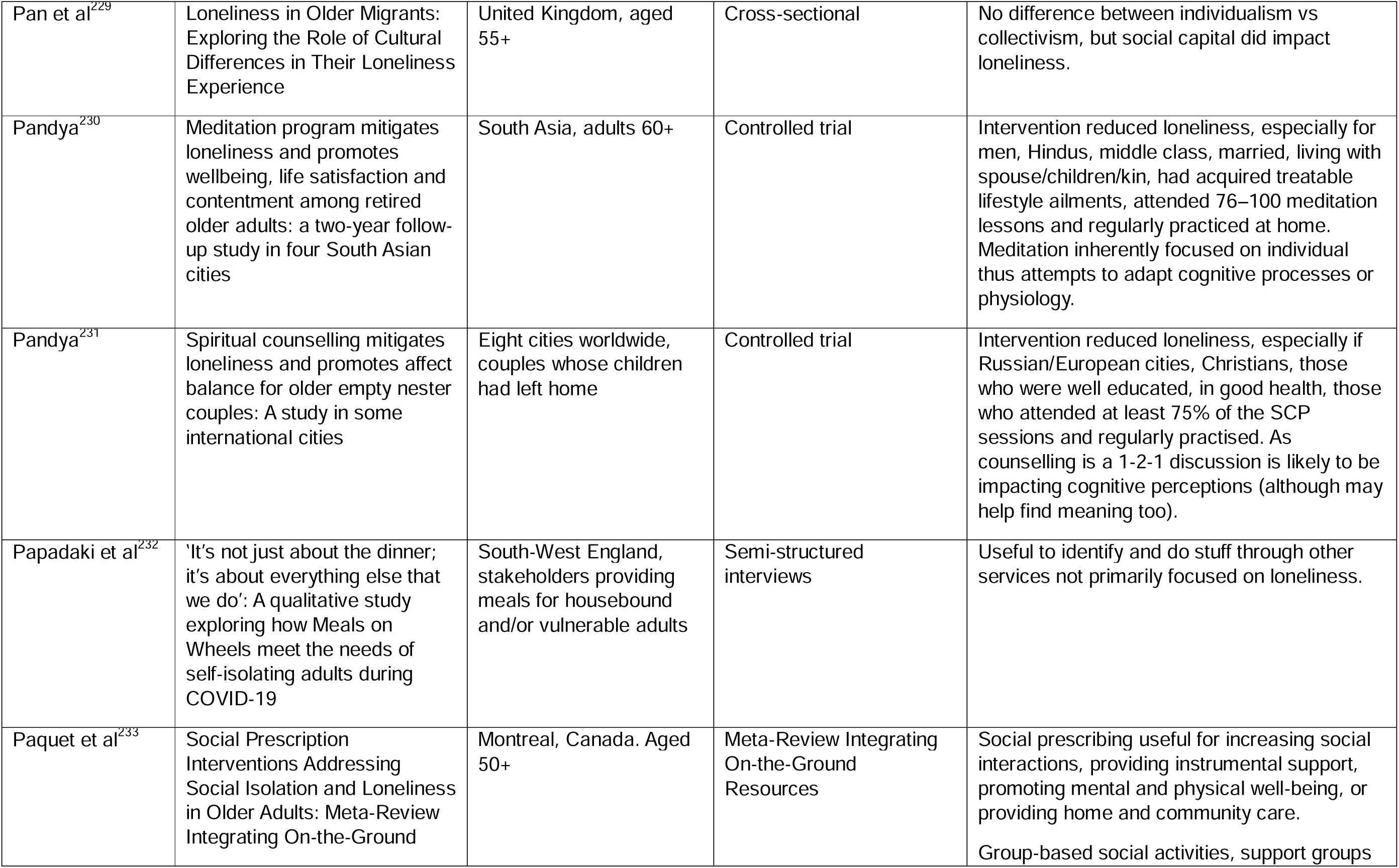
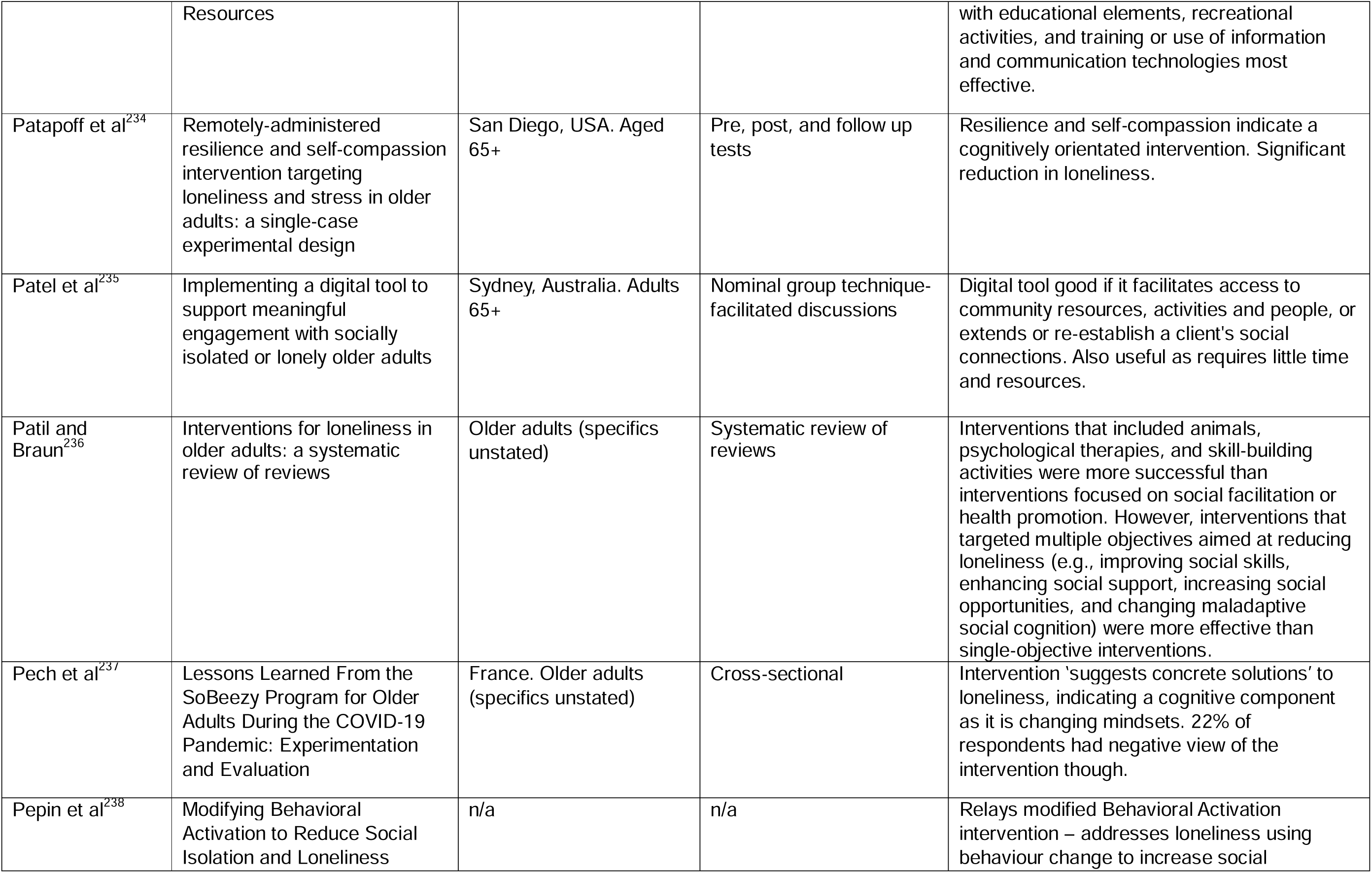
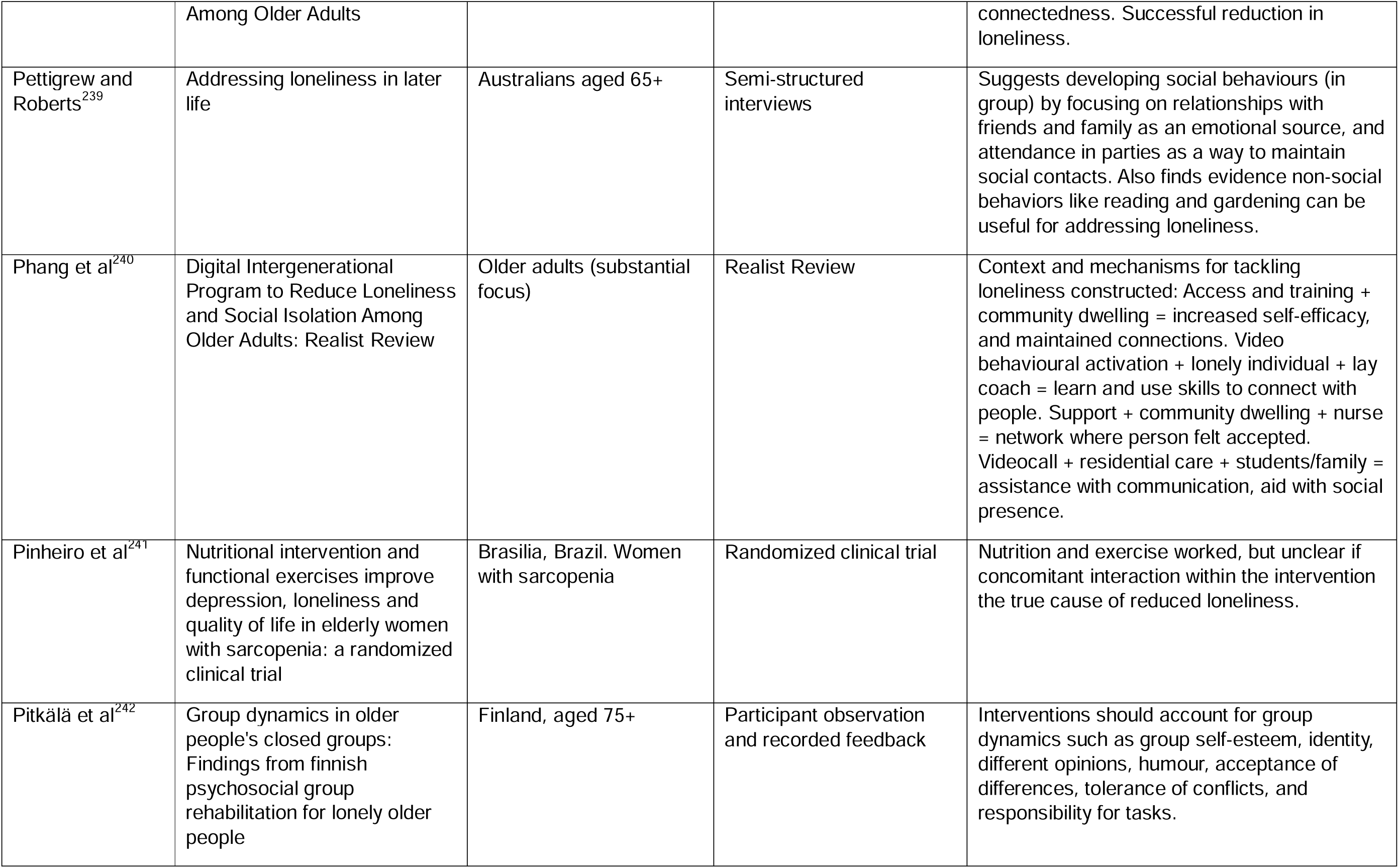
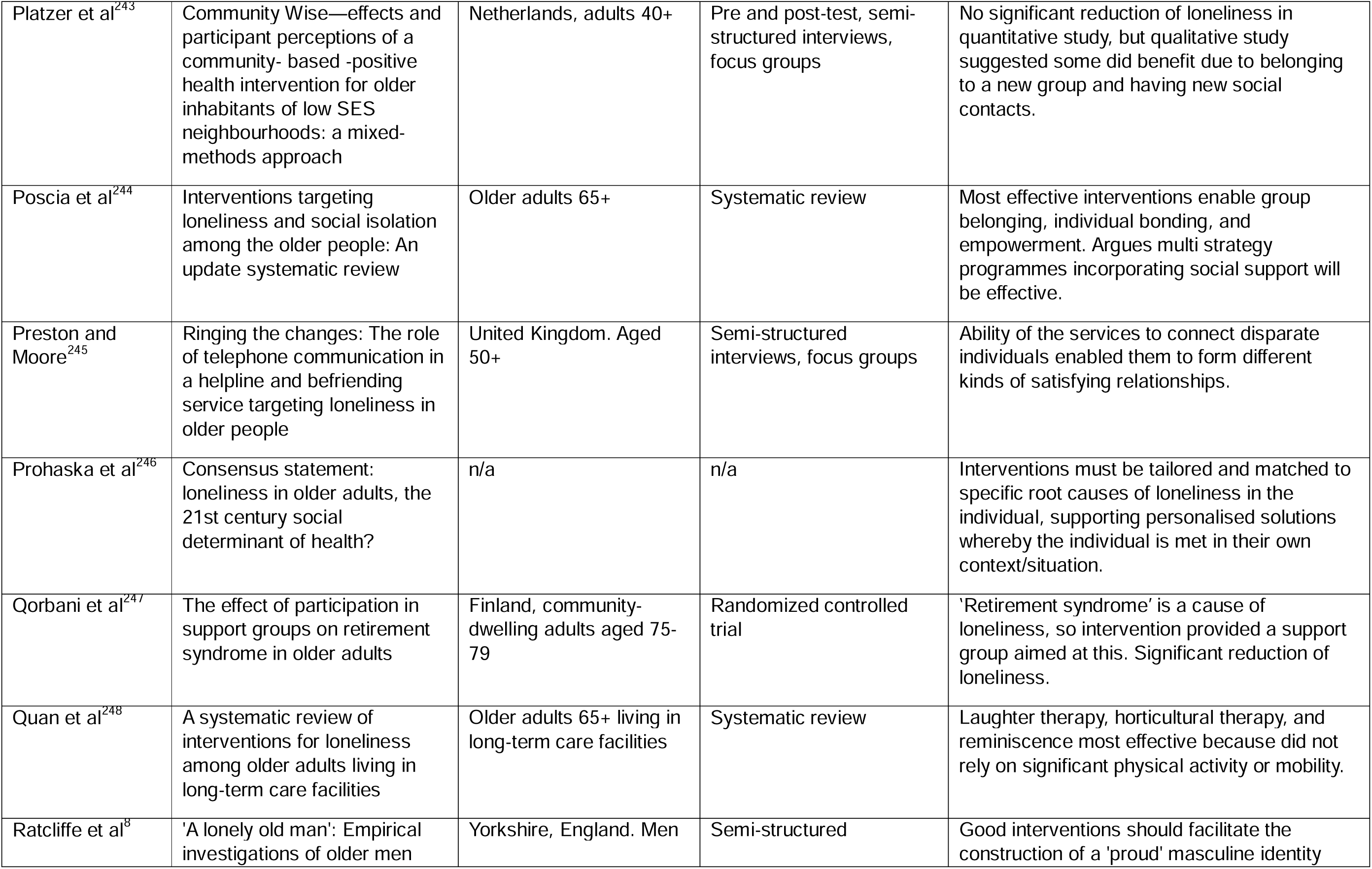
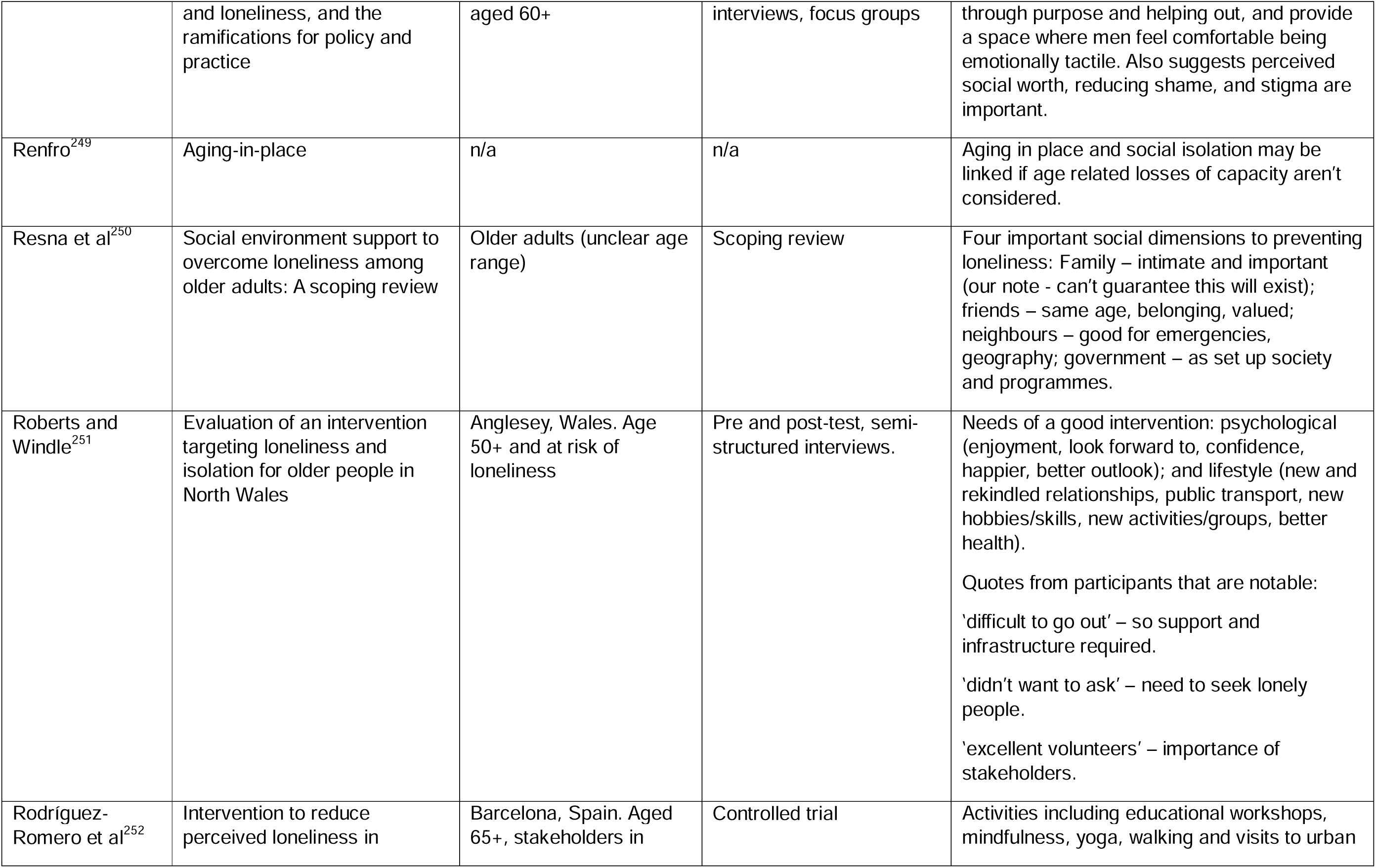
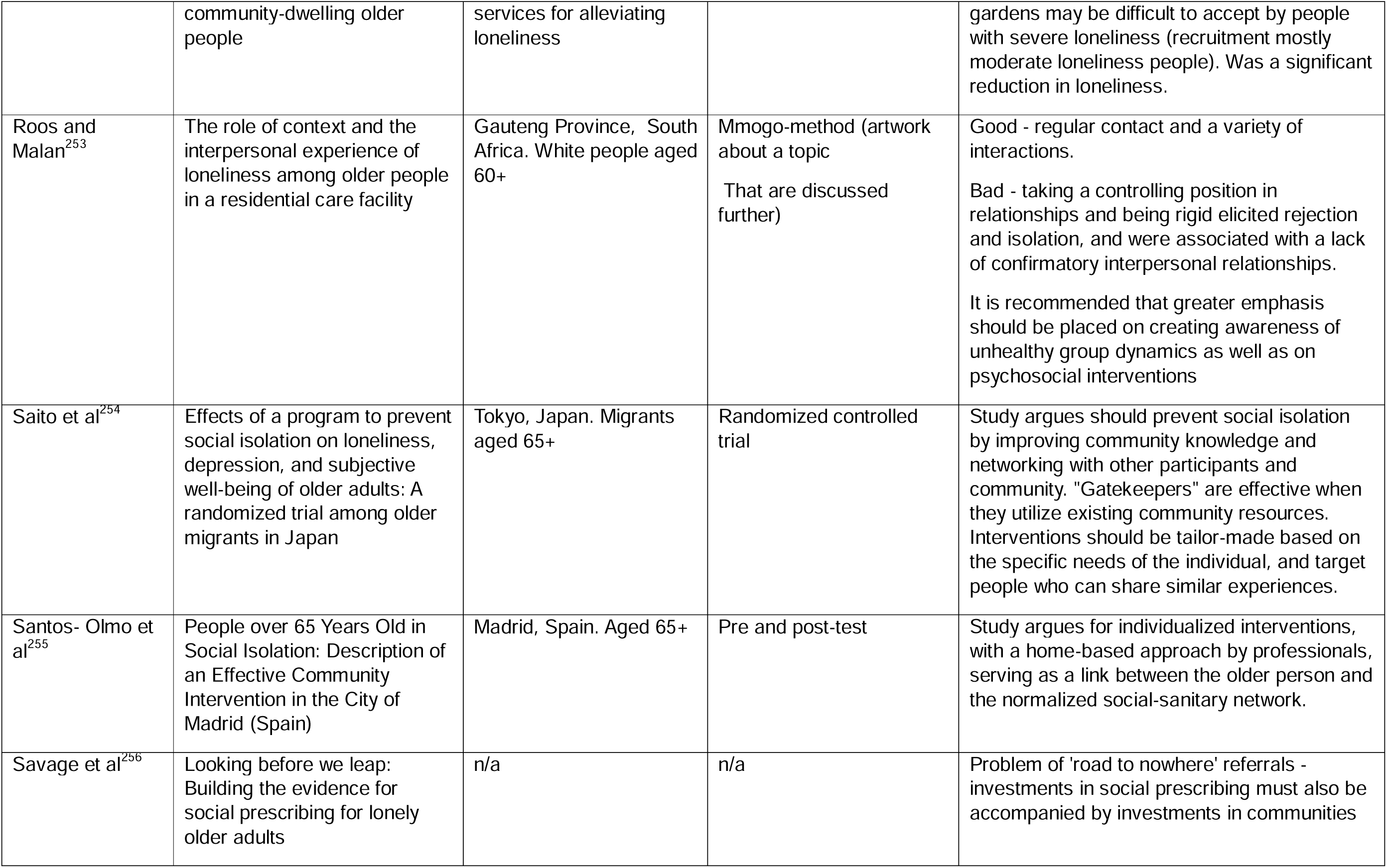
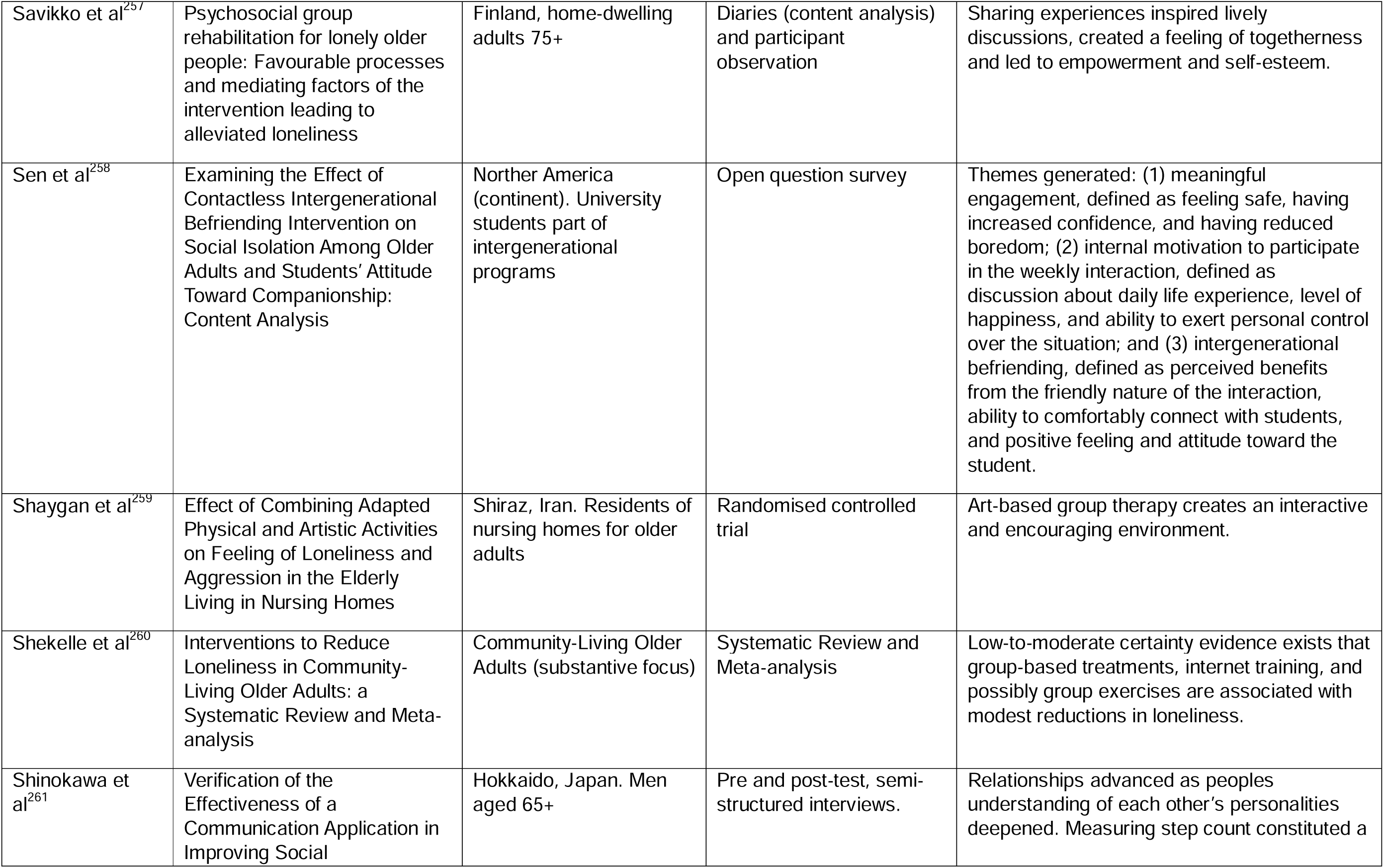
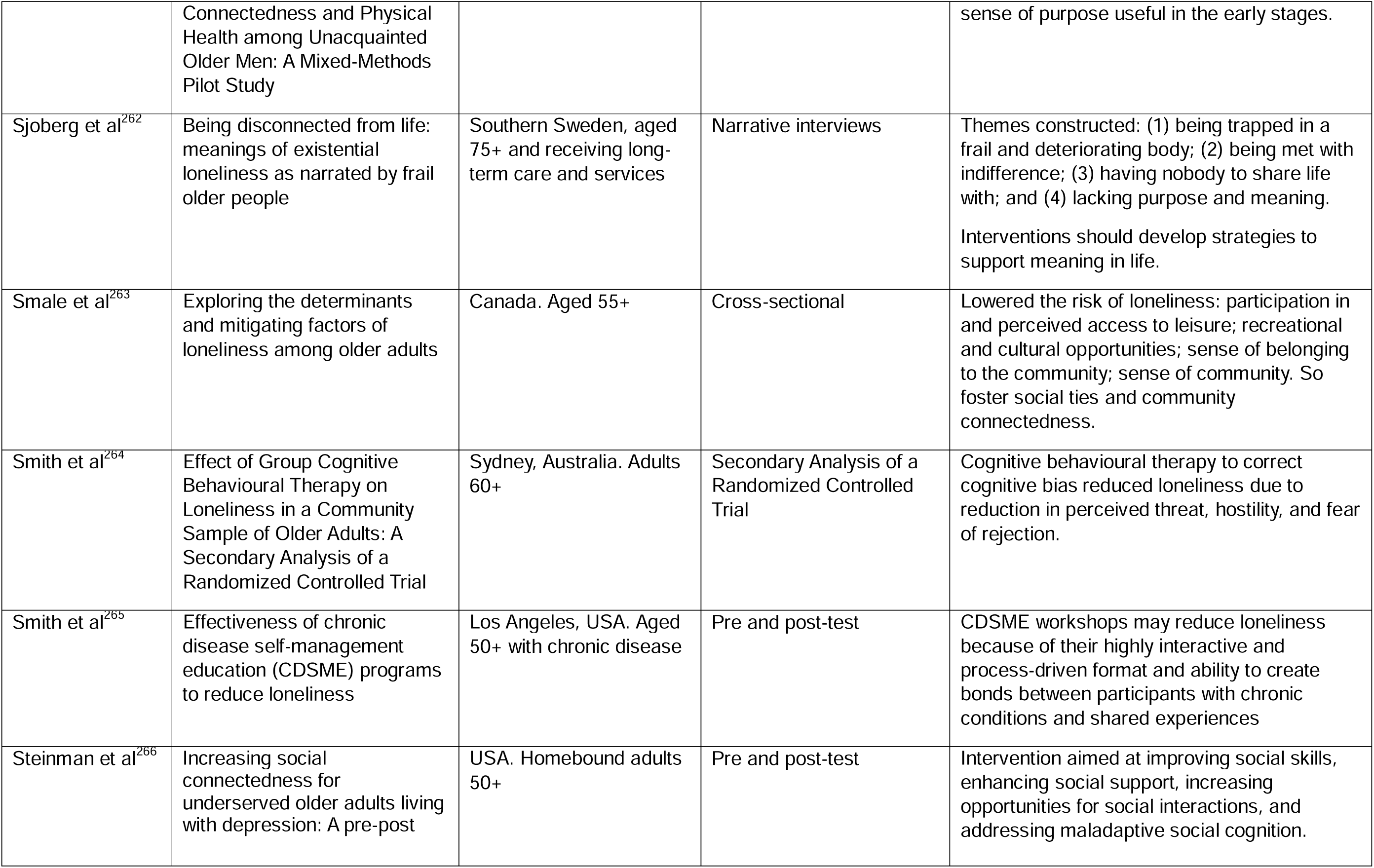
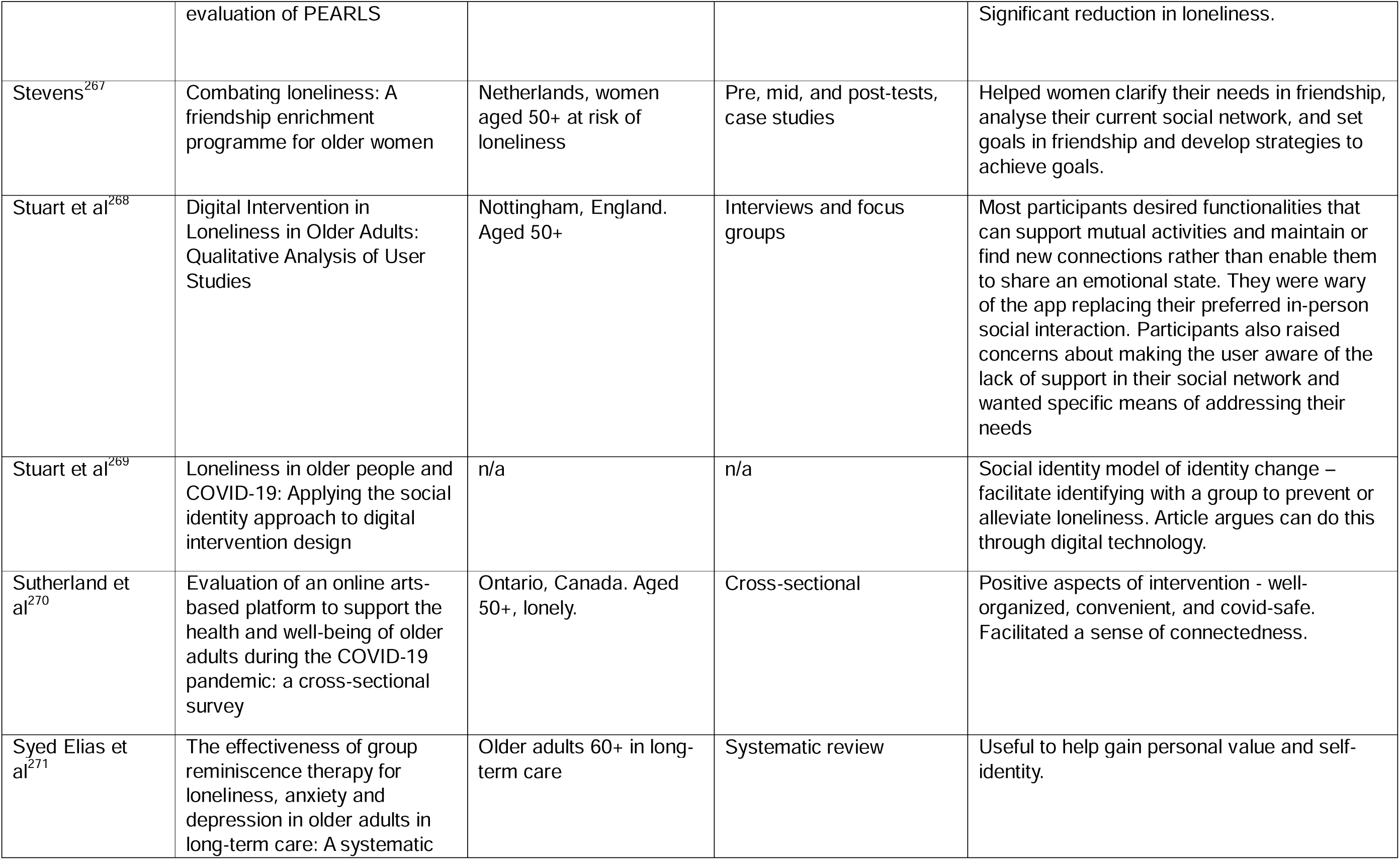
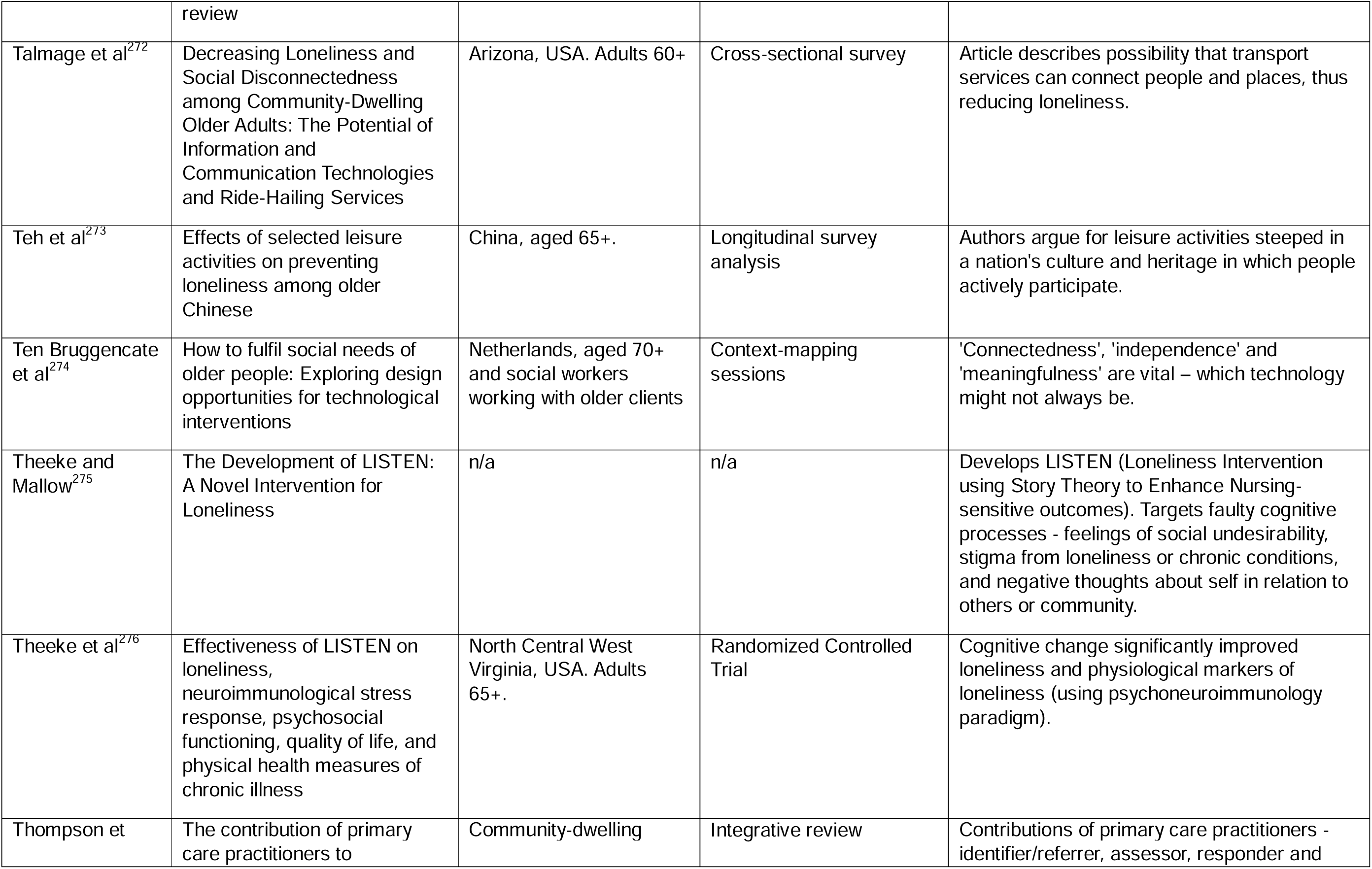
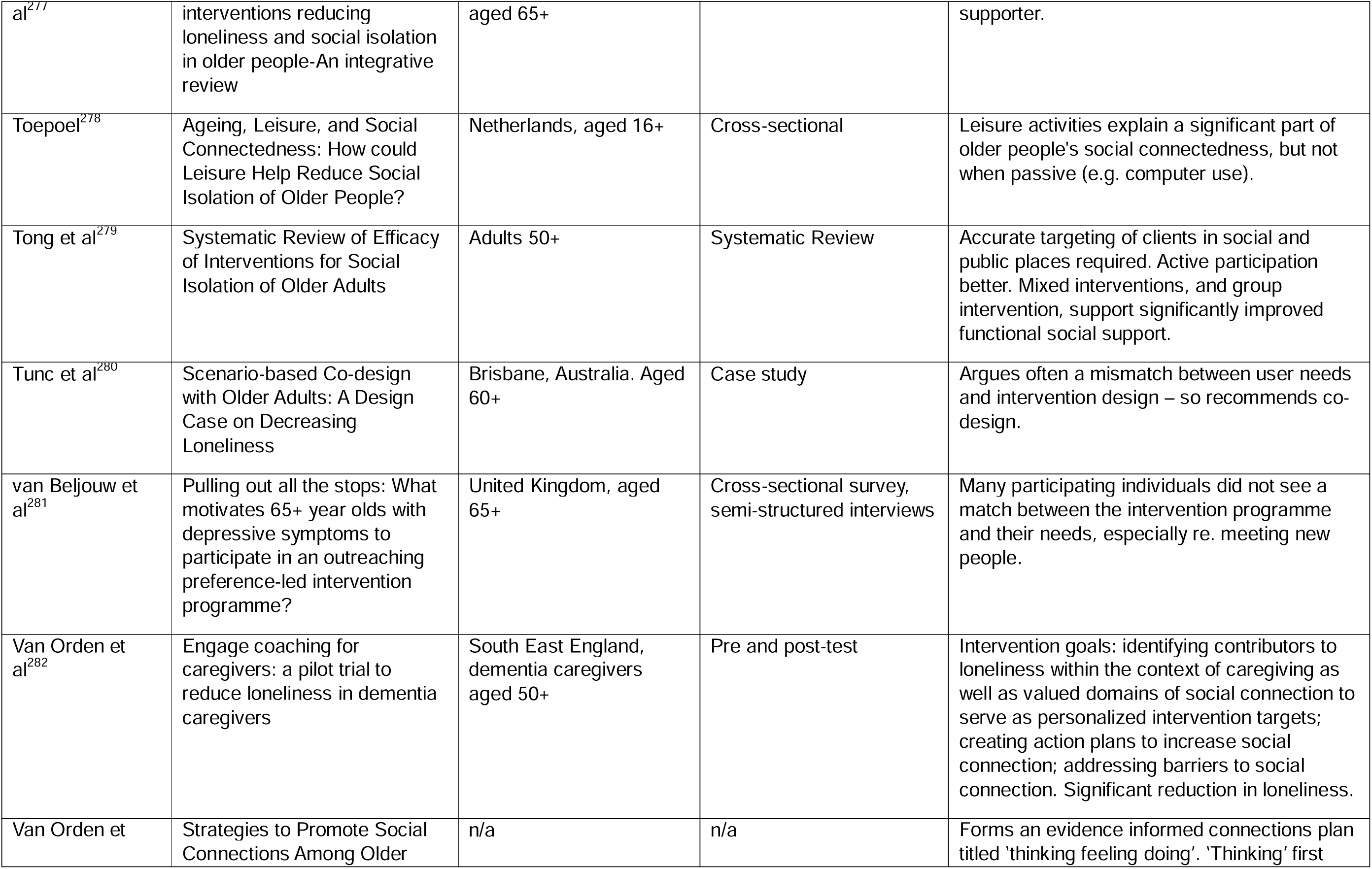
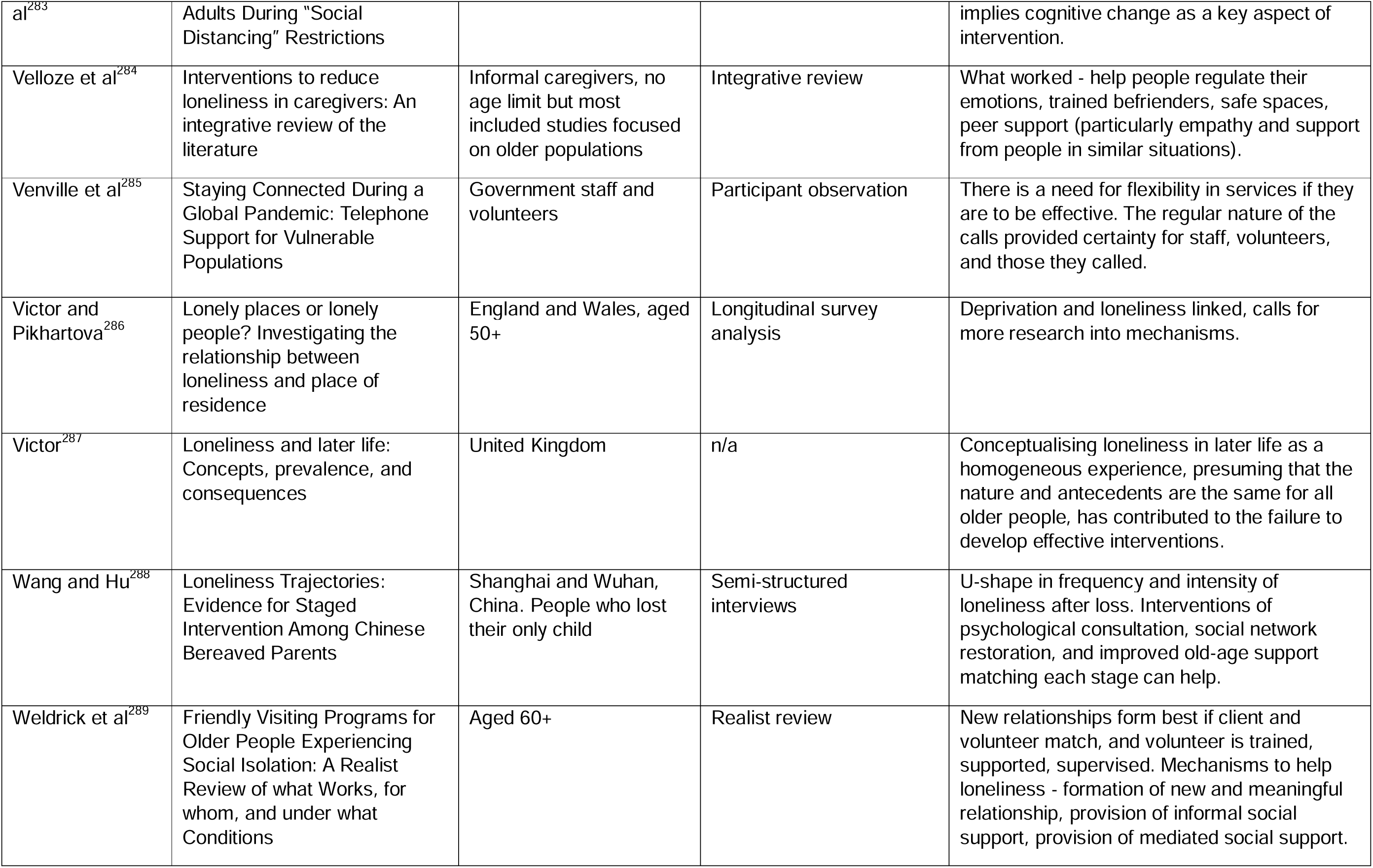
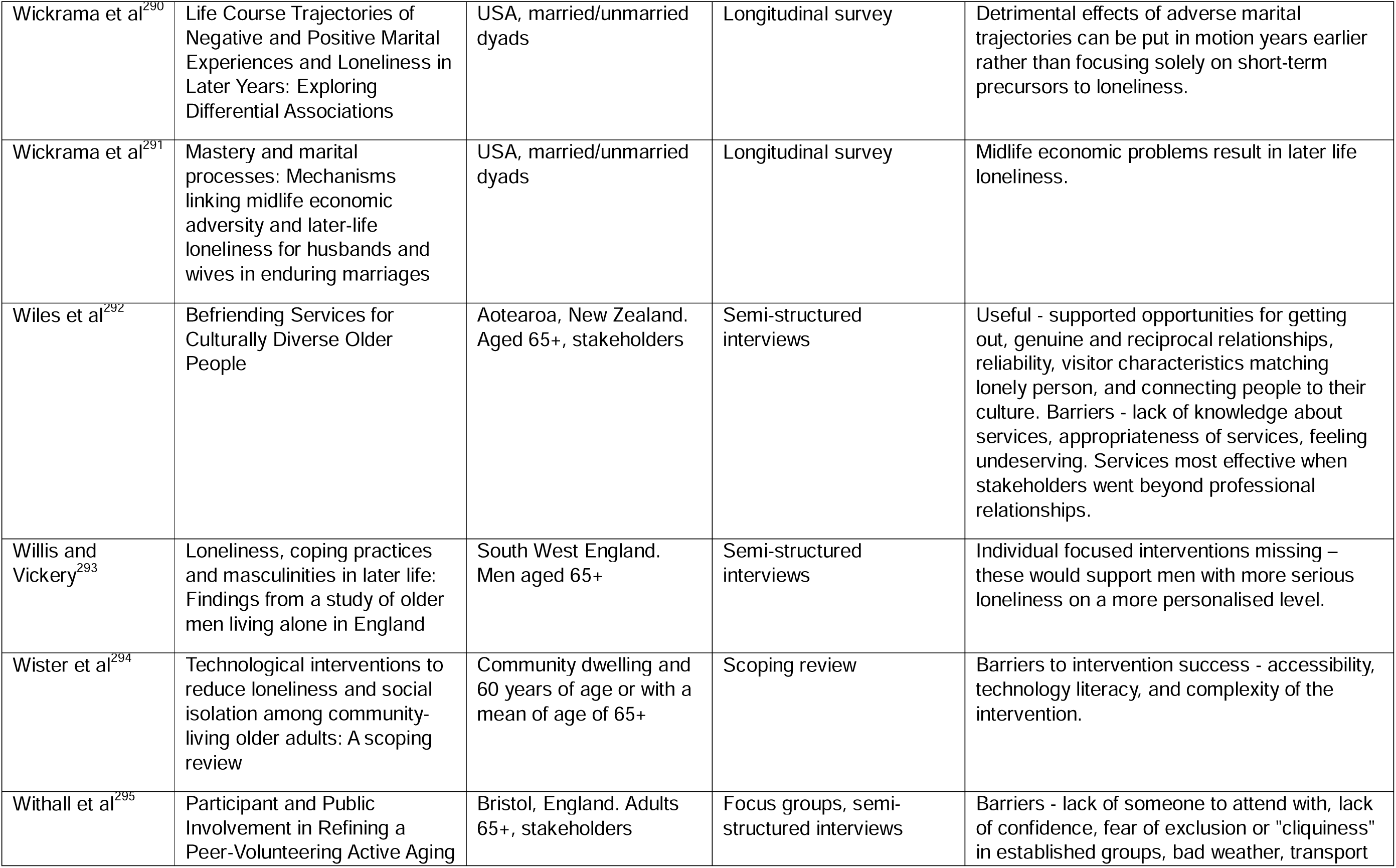
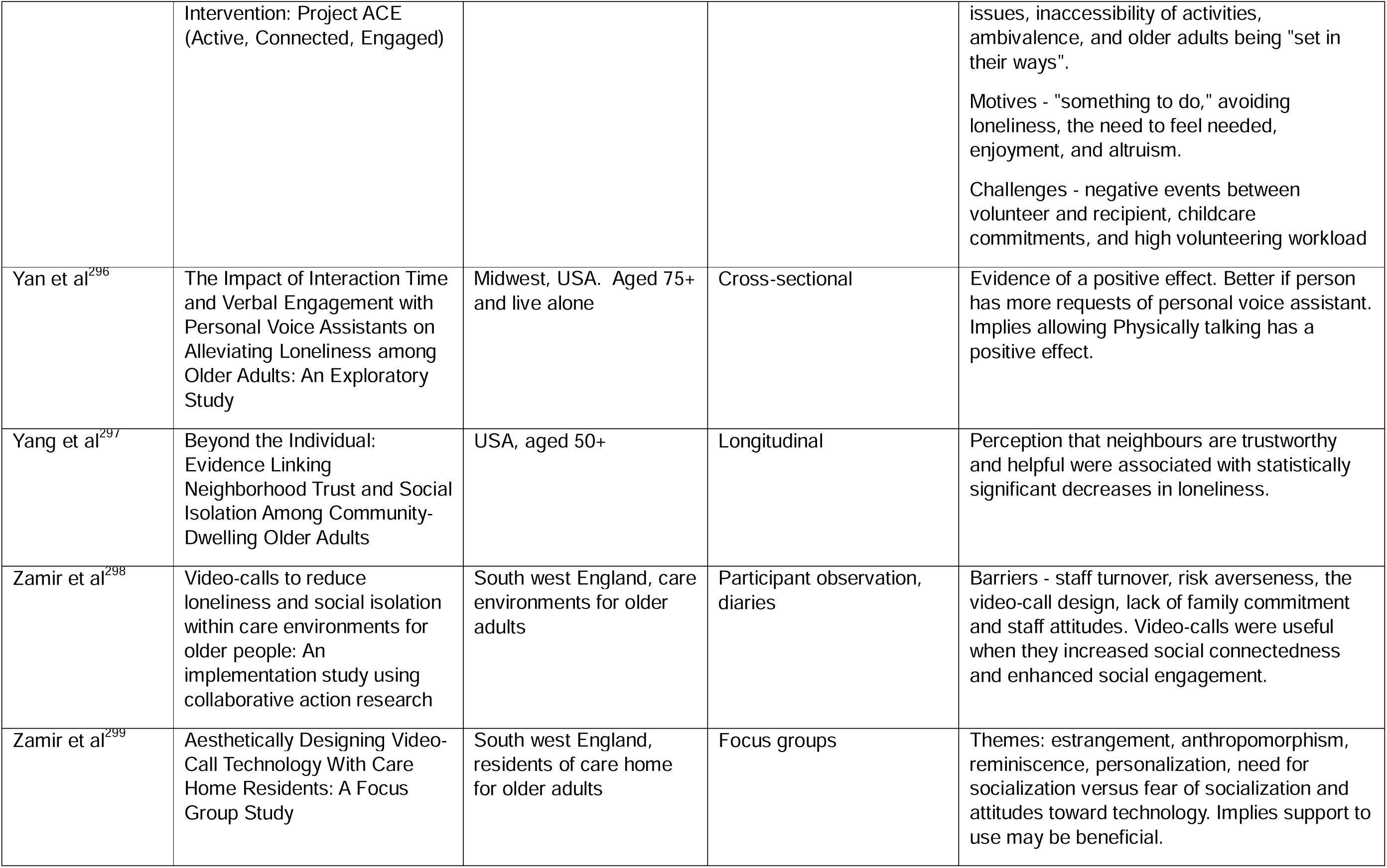
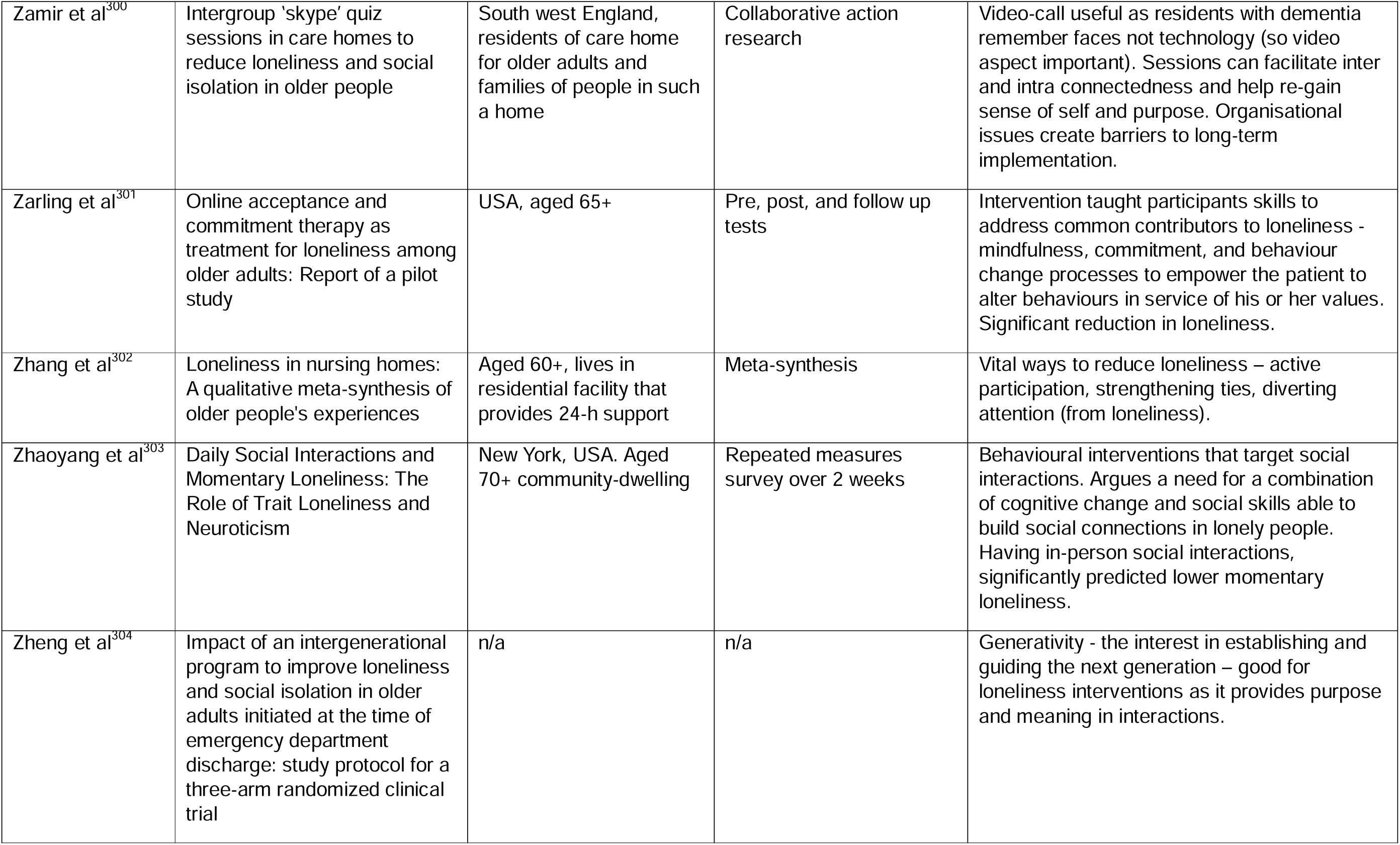
All Included articles and the relevant data/perspectives they provide.

### Synthesising arguments

We constructed four dimensions to defining the theoretical mechanisms and contextual requirements of a loneliness intervention. The *intended outcome* consists of the theoretical mechanism an intervention intends to elicit. *Level of intervention* refers to whether the intervention aims to directly aid an individual (micro), impact the environment around an individual (meso), or have a wider societal impact (macro). *Features of positive interventions* refer to factors that increase the likeliness of a successful intervention. *Emotions representing reduced loneliness* relay specific feelings that could explain a participant’s subjective reduction in loneliness. Figure 2 summarises how the intended outcomes, levels of intervention, and features of positive interventions combined to facilitate different emotions that represented a reduction in loneliness. The following sub-sections relay the sub-components of each dimension, and the evidence for its construction. The supplementary files further describe how the included articles influenced the formation of each dimension and sub-component.

**Figure 2.**
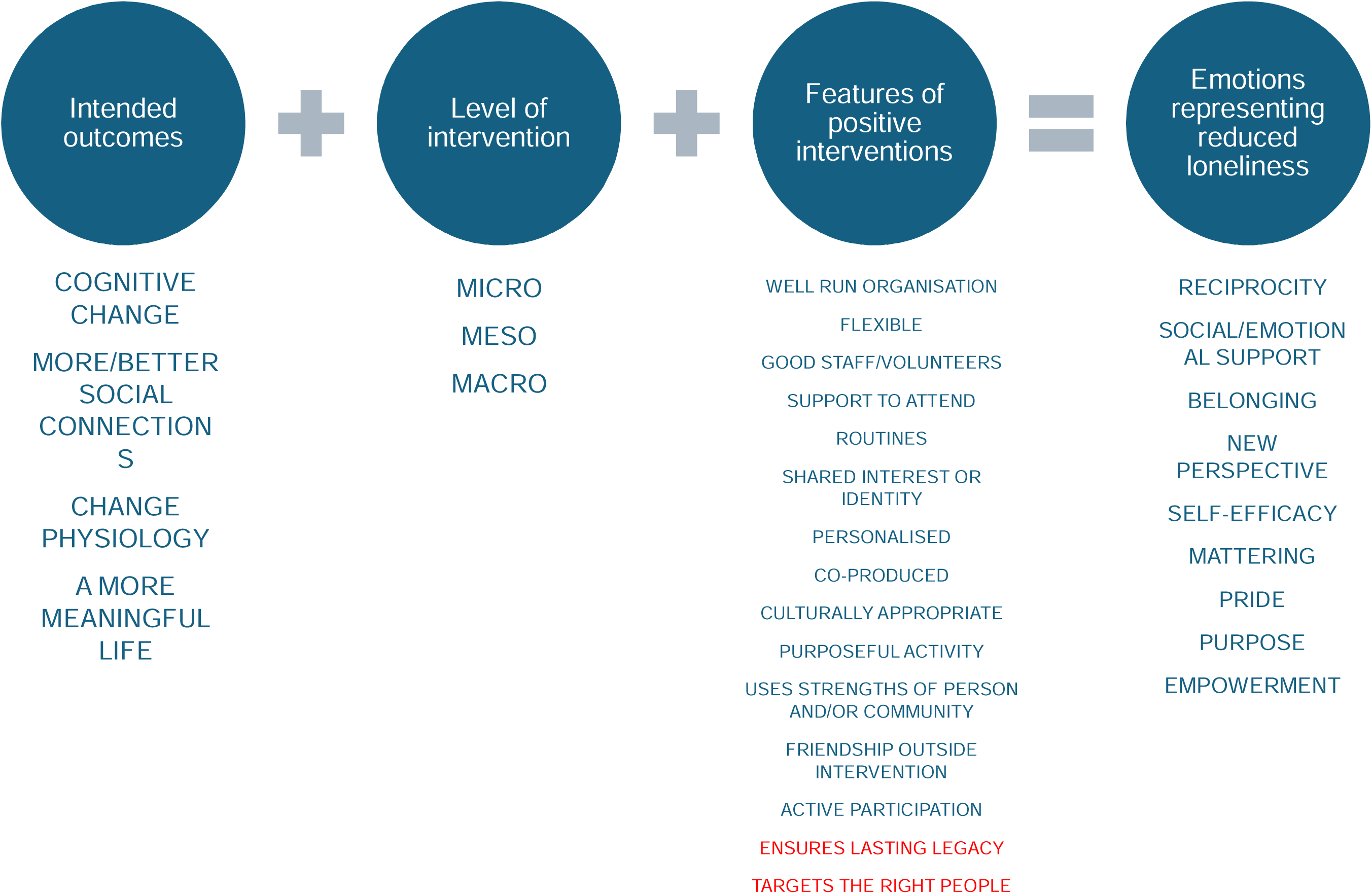
The contexts and mechanisms in interventions for loneliness in older adults.

### Intended outcome

This was constructed to show that interventions consisted of different intended pathways to reduce loneliness. Four categories were identified. These are not mutually exclusive, and articles evidencing the usefulness of ‘multidimensional’ interventions^43,84,147,217,236,244,266^ suggest it may be beneficial to incorporate all four. Interventions intending to induce cognitive change (supplementary file 1) assumed lonely people possess maladaptive social cognitions, therefore they attempted to induce greater satisfaction with existing relationships^116,135,160,217,267^ or address hostile and distrusting attitudes. ^61,179,264^ Cognitive interventions evidenced as effective included behavioural activation,^61,81,132,179^ mindfulness,^135,179^ cognitive behavioural therapy,^161,264^ ‘wisdom’ inducing,^174,215^ resilience-building,^174,234^ and storytelling.^275,276^ The second intended outcome was an improvement in a lonely person’s social connections (supplementary file 2). This could be by facilitating new connections,^44,73,91,174,194,224,243^ strengthening existing connections,^46,55,58,132,182,225,239^ or providing individuals with skills or tools for forming/maintaining connections.^14,15,39,50,111,132,139,154,240^

The third category of intended outcome consisted of interventions that aimed to impact a person’s physiological condition (supplementary file 3). These did not solely intend to change either a person’s social connections or their perception of them, but to elicit a direct neuropsychological change. This was inherent in interventions that indicated loneliness was reduced through exercise, ^39,136,156,241,278,295^ enjoyment,^40,72,148,251,295^ activities,^39,59,188,220,259,278,302^ and interactions with technology.^75,3,106,130,152,186,207^, The notion of a physiologically orientated intervention is consistent with work suggesting reduced oxytocin, a hormone released during moments of intimacy, constitutes the neuropsychological mechanism semiotically described as loneliness.^38^ However, Larkey et al^182^ found a communal Tai Chi intervention showed an inconsistent relationship between oxytocin release and loneliness.

The final intended outcome identified in the literature were attempts to facilitate a more socially meaningful existence (supplementary file 4). This could be through volunteering,^8,67,116,184,297^ activities promoting a sense of achievement,^76,105^ emphasising personal value,^60,271^ and empowerment.^103,114,202,216,244^ Though this represents a cognitive change in a lonely person’s perspective of their social situation, this type of cognitive change required an identifiable socio-structural component around which to define the meaning. They are therefore different to cognitive interventions which only referred to subjective satisfaction of social relationships.

### Level of intervention

This dimension considers whether an intervention aims to tackle loneliness by directly intervening with lonely individuals (micro), in an environment or community where loneliness has been identified (meso), or on a large scale where policies and structures may be changed to reduce loneliness (macro). Most evaluations included in this review assumed a micro scale. However, meso and macro interventions (supplementary file 5) are important as they can change the focus of tackling loneliness from ‘cure’ to ‘prevention’ by emphasising the environment in which loneliness might arise.^94^ Evidence has been found to suggest that loneliness can be addressed through architecture,^41,66,115^ access to transport,^120,251,272^ access to the internet,^211^ and financial resources.^291^ The availability of social resources may also be vital. Savage et al^256^ warn that social prescribing services require concomitant investment in communities lest they become a ‘road to nowhere’. A lack of ‘social capital’, defined as the cultural and structural resources for building social connections, may also provide the context for an individual’s loneliness.^93,104,229^ However, studies evaluating macro level campaigns and policies have found little impact on loneliness, although this may be due to the complexity of creating meaningful statistical comparisons.^47,190^

### Features of positive interventions

This dimension refers to specific factors that increase the likeliness of an intervention being successful. Most referred to how the intervention was organised (supplementary file 6): a well-run administration that is adaptable to user needs;^16,122,148,226,277,208,285,289^ enthusiastic and trained staff/volunteers;^114,129,148,164,172,211,251,284,292,298^ a supported pathway to attendance;^58,129,138,140,226,292,295,299^ routines that can be relied on and looked forward to;^88,97,98,121,134,157,177,183,219,227,285,292^ activities based on shared interests and/or identities;^54,70,92,122,148,173,176,177,193,212,223,265,269,284^ personalised interventions that tailor to people’s specific needs;^10,89,98,103,108,114,133,137,144,198,202,203,224,246,255,281,287,293^ interventions co-produced with the lonely older adults they aim to support;^70,117,129,133,162,175,208,280,281^ culturally appropriate interventions;^8,50,79,102,196,210^ interventions where people are required to actively participate;^16,89,133,147,162,204,259,265,295^ and interventions built on the strengths of the individuals ^8,50,169,248^ and communities ^16,54,100,193,222,254^ that use them. Additionally, though it cannot be specifically actioned, interventions that promote the continuation of friendships outside of the immediate intervention may be beneficial.^205,239,292^

Numerous articles indicated a need for interventions to have a lasting impact (supplementary file 7). ^85,129,192,205,208,261^ Theoretically, an intervention that only reduces loneliness for a short duration is of limited use, therefore lasting impact is given greater emphasis in figure 2 by highlighting it red. Authors have also relayed work emphasising a need to provide targeted interventions (supplementary file 8). Some have argued that those most in need may be relatively ‘hidden’,^71^ or less likely to contact and take part in interventions,^219,251^ therefore interventions need a plan to seek and support the loneliest people.^102^ Tong et al^279^ also conclude that interventions are more effective when they are targeted at specific group needs. Older men, for example, may be more likely to benefit from constructive activities.^8,212^ In figure 2, the broader phrase ‘target the right people’ is highlighted red to emphasise that interventions should target those most in need and those more likely to benefit from the specific intervention being actioned.

### Emotions representing reduced loneliness

This dimension relays specific feelings that interventions have facilitated to reduce loneliness (supplementary files 9-17). Important feelings reported were a sense of reciprocated interactions,^113,116,145,159,177,253^ emotional and social support,^77,98,123,143,146,279,289^ belonging,^63,109,156,173,177,181^ a new perspective on one’s social situation,^116,177,267,271^ self-efficacy,^74,78,184,194,199,232^ mattering,^63,118^ pride,^8,42,78,127,271^ purpose,^60,116,136,206,225,261,262,300^ and empowerment.^147,202,257,267,281,301^ These differ from the intended outcomes in figure 2 as the intended outcomes constitute theoretical perspectives on how the intervention can reduce loneliness, whereas these emotions are mediating subjective states that resulted from the intervention regardless of its intention. In Kikuchi et al^177^, for example, facilitating improved social connections (intended outcome) led to a sense of belonging (the emotion representing reduced loneliness).

Further displaying the relevance of each dimension in figure 2, Kikuchi et al’s work also emphasised a need to consider people’s role within their community (a meso level focus) and facilitate connections through identification with their community (a shared identity). In figure 2, these emotions are not connected to specific intended outcomes, levels, or intervention features because it is theoretically possible for many combinations to occur. Nevertheless, logically, different combinations of the first three dimensions are more likely to result in different emotions. A sense of perspective, for example, would seem more likely to occur from cognitive change interventions. Pride, on the other hand, may be more likely to occur in an intervention aiming to facilitate a more meaningful life. Interventions should consider what specific emotions they wish to elicit, and what combinations of theoretical intention, level, and features are likely to elicit them.

## Discussion

Figure 2 summarises how the intended outcome of a loneliness intervention, the level at which it is conducted, and the positive features it provides, can elicit a variety of emotions that are the foundation to reduced loneliness. This emphasises that the key components for a successful loneliness intervention are multidimensional, will not be the same for different groups and individuals, and may not be related to the activities the intervention forms nor match the assumed goals of interventions. The sub-components of each dimension are not mutually exclusive, and facilitating as many as possible may be helpful for even a small intervention. Policy, practice, and research can use these dimensions and their sub-components as a guide to ensure sufficient focus on what truly matters for tackling loneliness.

Distinguishing between the efficacy of intervention activities, such as by comparing animal assisted therapy, technological interventions, art therapy, etcetera, may be of limited usefulness in the design of interventions. Few of the factors represented in these dimensions represent a specific activity yet each have been evidenced as the cause of positive impacts on loneliness. Rather, the popularity and efficacy of an activity may depend on whether it was personalised and co-produced. At a wider level, it would seem more logical to facilitate a variety of activities able to target different people than identify which activity is ‘best’. Evaluations should focus on the quality of the delivery of the intervention and its relevance to the people involved.

The four ‘intended outcomes’ parallel debates around conceptualising loneliness. Cognitive change and facilitating better social connections both operate from a conceptualisation of loneliness as a subjective lack of social relationships.^19,70^ Changing a person’s physiology, however, assumes that neuropsychological processes are a vital component of loneliness in a manner separate from a lonely individual’s conscious perspectives. Many of the included studies aimed at inducing physiological change provided concomitant social interaction,^136,214,241,248^ rendering the true cause of reduced loneliness difficult to ascertain. However, even if concomitant social interaction is required to alleviate loneliness, the physiological components of these interventions may still be important.

The importance of physiology to loneliness interventions was also partly evidenced by noting that interactions with technology such as robots or ChatGPT were seemingly able to reduce loneliness. In the current review, this was interpreted as indicating that the opportunity to speak and interact may have a neuropsychological impact. This could also be interpreted as representing a mimicked social relationship impacting the person’s subjective perspective of their relationships. Nevertheless, given the established importance of neuropsychology to loneliness,^305^ and that exercise, enjoyment, increased activity, and oxytocin release have been evidenced as able to reduce loneliness, we argue that it is more logical to interpret interactions with technology as able to induce neuropsychological change.

Loneliness research and interventions have been criticised for being overly biomedical and individualised, ignoring the social context in which loneliness arises.^20,306^ Cognitive and physiological interventions are inherently focused on individuals, but the fourth type of intended outcome, facilitating a more meaningful life, is deeply intertwined with social context as it requires something to build meaning from. The second dimension, level, further highlights that interventions should look beyond individuals. Despite the evidence in favour of physiological and cognitive factors, then, the current review supports these criticisms and recommends that researchers and practitioners ensure interventions are designed and evaluated with due consideration of social contexts and inequalities.

A short-term alleviation of loneliness is unlikely to offer long-term benefits to chronically lonely individuals, yet few included studies utilised a longitudinal perspective. Follow up studies, such as those conducted by Bruce et al,^61^ will strengthen the evidence base. Some of the above features of positive interventions, such as ‘routines’ and ‘friendships outside of intervention activities’, indicate a transition from being an intervention activity to being a spontaneous and normalised aspect of life. Interventions that can provide this may be particularly effective for producing long-term impact.^97,105,156^ A macro-level need to consider resources may be necessary where older adults do not have access to opportunities outside of intervention activities.^141,211,256^

The dimensions presented in figure 2 are not inherently age specific, but the impact of age is implicit throughout. The importance of building on the strengths of individuals was highlighted through research emphasising the need for age-friendly activities,^169^ and technological interventions were often framed around overcoming age-related losses in physical capacity^39^ or ensuring continued access to new forms of socialising and belonging.^126,181^ A need to challenge the marginalising aspects of age were highlighted alongside a need for mattering,^118^ pride^8^, and empowerment,^103^ and co-producing culturally aware and personalised interventions can ensure age-suitable interventions.^196^ This may also be beneficial for targeting and supporting people and communities who are less likely to attend or connect with people in mainstream groups and services.

### Limitations

The search strategy was not systematic, therefore the relative importance and efficacy of each dimension and its sub-components cannot be ascertained. Though we included a wide variety of literature, these findings are unlikely to be exhaustive. A wealth of different terminology could arguably be used in place of the terminology constructed in figure 2. ‘Generativity’, for example, has been used in work on ageing and loneliness to represent a need for meaningful contributions to society,^42^ thus could replace ‘pride’, ‘purpose’, and ‘meaning in life’ as they are constructed in this article. Nevertheless, these three phrases represent distinct components of the theory formed in the current review, and are more commonly used terminology thus easier to consider in intervention contexts.

## Conclusions

To our knowledge, this is the most complete construction of the theoretical dimensions relevant to loneliness interventions in older adults to date. Rather than suggesting specific activities are more effective than others, it highlights that interventions must be part of a multidimensional policy and practice response that can work for different groups and individuals with different needs. Future interventions and research should draw on these dimensions to ensure policy and practice is contextualised according to who it aims to benefit, how it will fit within a wider policy and practice response, and how it aims to work.

## Data Availability

All data produced in the present work are contained in the manuscript

## Abbreviations

CIS: Critical Interpretative Synthesis
PRISMA: Preferred Reporting Items for Systematic reviews and Meta-Analyses

## Acknowledgements

Thank you to all the academics that responded to our email request with articles and ideas (as specified in the search strategy):

Dr Samia Akhter-Khan, King’s College London

Dr Lili Golmohammadi, King’s College London

Professor Paul Willis, Cardiff University

Dr Baptiste Brossard, University of York

Dr Kathryn Cunningham, University of Dundee

Dr Christian Langkamp, University of Oxford

And also thanks to the Global Health and Social Medicine ageing group at King’s College London who listened to my presentation and contributed with useful insight and criticism.

