## Supplementary file 1 for "Conceptualising the context and mechanisms for tackling loneliness in older adults through interventions: A Critical Interpretive Synthesis"

Supplementary file 1. Papers indicating cognitive change can elicit reduced loneliness

| **Article title** | **Reason for inclusion** |
| --- | --- |
| A Systematic Literature Review of Loneliness in Community Dwelling Older Adults^101^ | Finds evidence that improving personal and psycho-social resources for older people fared better outcomes than those focused on technology or social connections. |
| An evaluation of a low-intensity cognitive behavioral therapy mhealth-supported intervention to reduce loneliness in older people^161^ | Intervention designed because maladaptive cognitions affect the maintenance and establishment of meaningful social connections. This study implemented and evaluated a low-intensity Cognitive Behavior Therapy mHealth- intervention on whatsapp. Significantly reduced loneliness. |
| Behavioural activation to mitigate the psychological impacts of COVID-19 restrictions on older people in England and Wales (BASIL+): a pragmatic randomised controlled trial^132^ | Behavioural activation sessions facilitate cognitive change encouraging socially reinforcing activities and help maintain social connections. Statistically significant reduction in loneliness. |
| Benefits of a psychosocial intervention programme using volunteers for the prevention of loneliness among older women living alone in Spain^194^ | Improved self-efficacy in managing loneliness improvements, suggesting cognitive change. |
| Changes in Quality of Life and Loneliness Among Middle-Aged and Older Adults Participating in Therapist-Guided Digital Mental Health Intervention^135^ | Mindfulness can be interpreted as cognitive change as it focuses on individual resilience. |
| Combating loneliness: A friendship enrichment programme for older women^267^ | Intervention helped women clarify their needs in friendship, analyse their current social network, set goals in friendship and develop strategies to achieve goals. |
| Commentary: An innovative intervention to reduce perceived stress and loneliness^174^ | Resilience and wisdom inducing – cognitive notions – as vital. |
| Daily Social Interactions and Momentary Loneliness: The Role of Trait Loneliness and Neuroticism^303^ | Behavioural interventions that target social interactions. Argues a need for a combination of cognitive change and social skills able to build social connections in lonely people. |
| Dual-Process Bereavement Group Intervention (DPBGI) for Widowed Older Adults^84^ | DPBGI better than traditional approaches as multidimensional. Its key components, loss-oriented coping, restoration-oriented coping, and oscillation are focused on cognitive change. |
| Effect evaluation of a two-year complex intervention to reduce loneliness in non-institutionalised elderly Dutch people^151^ | Psychosocial group courses, alongside mass media campaign, information meetings, social activities, and training , were implemented– but no effect on social support and loneliness, only on ‘loneliness literacy’ (suggesting some favourable cognitive change without it improving wellbeing). |
| Effect of Group Cognitive Behavioural Therapy on Loneliness in a Community Sample of Older Adults: A Secondary Analysis of a Randomized Controlled Trial^264^ | Cognitive behavioural therapy to correct cognitive bias reduced loneliness due to reduction in perceived threat, hostility, and fear of rejection. |
| Effect of psychosocial care model applied in an 'elderly day care center' on loneliness, depression, quality of life, and elderly attitude^112^ | Significant reduction of loneliness. Studies argues is because intervention meets psychosocial care needs. Does so via 9 weekly sessions, implies cognitive change and a little interaction is the cause. |
| Effectiveness of LISTEN on loneliness, neuroimmunological stress response, psychosocial functioning, quality of life, and physical health measures of chronic illness^276^ | Cognitive change significantly improved loneliness and physiological markers of loneliness (using psychoneuroimmunology paradigm). |
| Engage coaching for caregivers: a pilot trial to reduce loneliness in dementia caregivers^282^ | Creating action plans to increase social connection and addressing barriers to social connection part of the coaching |
| Evaluation of an intervention in general practices to strengthen social activities in older patients - A qualitative study of patients' experiences in the project HoPES3^199^ | Increasing self-efficacy and quality of life via raising awareness of spiritual needs, promoting social contacts and self-care – suggests a combination of cognitive change and need for social connections. |
| Facing the Next “Geriatric Giant”—A Systematic Literature Review and Meta-Analysis of Interventions Tackling Loneliness and Social Isolation Among Older Adults^107^ | Addressing social cognition one of the most effective intervention types. |
| Feasibility, acceptability, and preliminary efficacy of an internet-based CBT intervention for loneliness in older adults: A pilot RCT^108^ | Argues “despite evidence showing that addressing maladaptive social cognition is the most effective intervention strategy for reducing loneliness, most existing programs aimed at older individuals do not use that method” |
| Improving Social Connectedness for Homebound Older Adults: Randomized Controlled Trial of Tele-Delivered Behavioral Activation Versus Tele-Delivered Friendly Visits^81^ | Tele behavioural activation (a form of cognitive intervention) more successful than ‘friendly visits’ (which are less overtly cognitively orientated). |
| Increasing social connectedness for underserved older adults living with depression: A pre-post evaluation of PEARLS^266^ | Includes improving social skills, enhancing social support, increasing opportunities for social interactions, and addressing maladaptive social cognition*.* So multidimensional but specific focus on need for cognitive change. |
| Interventions against social isolation of older adults: A systematic review of existing literature and interventions^202^ | Social activation, empowering and motivating interventions, and interventions where participants able to control loneliness better the cognitively orientated findings. |
| Interventions for loneliness in older adults: a systematic review of reviews^236^ | Psychological therapies and skill-building activities were more successful than interventions focused on social facilitation or health promotion. However, interventions that targeted multiple objectives aimed at reducing loneliness (e.g., improving social skills, enhancing social support, increasing social opportunities, and changing maladaptive social cognition) were more effective than single-objective interventions. |
| Interventions that have potential to help older adults living with social frailty: a systematic scoping review^170^ | Self-management the best way to reduce loneliness. |
| Interventions to reduce loneliness among Chinese older adults: A network meta-analysis of randomized controlled trials and quasi-experimental studies^191^ | Hybrid and psychological interventions appeared to be advantageous for interventions, indicating a need for a partially or wholly cognitive orientation. |
| Interventions to reduce loneliness in caregivers: An integrative review of the literature^284^ | Interventions that attempt to help people regulate their emotions were among the most effective type. |
| It Gave Me Somebody Else to Think About Besides Myself: Caring Callers Volunteer Experiences With a Telephone-Based Reassurance Program for Socially Isolated Older Adults^116^ | Gaining perspective, i.e., a more balanced and forgiving view of one’s interpersonal relationships, among the key factors for the success of the intervention. |
| Layperson-Delivered Telephone-Based Behavioral Activation Among Low-Income Older Adults During the COVID-19 Pandemic: The HEAL-HOA Randomized Clinical Trial^179^ | Behaveoural activation built on principles of cognitive change. UCLA significantly reduced in the behavioural activation group and mindfulness compared with befriending. But only behavioural activation group reduced when using De Jong-Gierveld scale. |
| Lessons Learned From the SoBeezy Program for Older Adults During the COVID-19 Pandemic: Experimentation and Evaluation^237^ | Intervention ‘suggests concrete solutions’ to loneliness, indicating a cognitive component as it is changing mindsets. 22% had negative view of the intervention though. |
| Loneliness and social isolation of older adults: Why it is important to examine these social aspects together^218^ | Using a distinction between loneliness as subjective and isolation as objective state to suggests 4 types of interventions. Intervening for people that are lonely but not isolated suggests a need for cognitive change given that such people have social connections. |
| Loneliness in Old Age: Interventions to Curb Loneliness in Long-Term Care Facilities^59^ | Interventions aiming to change cognition found to be effective. |
| Loneliness in older persons: A theoretical model and empirical findings^91^ | Model argues loneliness is interaction of cognitive processes and environmental events. |
| Loneliness matters: a theoretical and empirical review of consequences and mechanisms^146^ | Addressing of maladaptive social cognitions presented as a key component to tackling loneliness. |
| Managing loneliness: a qualitative study of older people’s views^171^ | Notion of ‘positive attitude’ mirrors a need for cognitive. |
| Meditation program mitigates loneliness and promotes wellbeing, life satisfaction and contentment among retired older adults: a two-year follow-up study in four South Asian cities^230^ | Meditation inherently attempts to adapt cognitive processes. |
| Modifying Behavioral Activation to Reduce Social Isolation and Loneliness Among Older Adults^238^ | Behavioral Activation is a cognitive intervention aimed at then changing behaviour. Successful reduction in loneliness. |
| One Year Impact on Social Connectedness for Homebound Older Adults: Randomized Controlled Trial of Tele-delivered Behavioral Activation Versus Tele-delivered Friendly Visits^61^ | Behavioral Activation is a cognitive intervention aimed at then changing behaviour. Improvements maintained at 1 year follow up. |
| Online acceptance and commitment therapy as treatment for loneliness among older adults: Report of a pilot study^301^ | Teach participants skills to address common contributors to loneliness - mindfulness, commitment, and behaviour change processes to empower the patient to alter behaviours in service of his or her values. Significant reduction in loneliness. |
| Qualitative study of loneliness in a senior housing community: the importance of wisdom and other coping strategies^215^ | Some coping strategies to prevent or overcome loneliness had a cognitive orientation (acceptance of aging, compassion) |
| Remotely-administered resilience and self-compassion intervention targeting loneliness and stress in older adults: a single-case experimental design^234^ | Resilience and self-compassion are cognitive notions – worked vs pretest (no control group). |
| Screening — An Important Starting Point for Effective Loneliness Interventions among Older Adults^160^ | Strong focus on perception of negative relationships in article’s recommendations -  1) increasing contacts of an older adult and reducing one’s perceived discrepancy between actual and desired relationships, 2) decreasing relationship expectations to meet realities, and 3) reducing the effect of the discrepancy by coping with experiences of loneliness. |
| Self-affirmation training can relieve negative emotions by improving self-integrity among older adults^95^ | Self-affirmation didn’t impact loneliness (just other negative emotions). So cognitive aspect not effective for loneliness here. |
| Social and physical effects of a pedometer and communication application among older men: a mixed-methods, pre/post pilot study^36^ | According to authors, part of social connectedness is ‘constructing reliance’, ‘Remembering to consider friends’, ‘Awareness of friends’ daily lives and encouragement for me’, and ‘Taking an interest in friends’ – all suggests a cognitive awareness of friends as part of what is important. |
| Spiritual counselling mitigates loneliness and promotes affect balance for older empty nester couples: A study in some international cities^231^ | Intervention reduced loneliness. As counselling is a wholly 1-2-1 experience is likely to be impacting cognitive perceptions (although may help find meaning too). |
| Strategies to Promote Social Connections Among Older Adults During “Social Distancing” Restrictions^283^ | Evidence informed Connections Plan – thinking feeling doing. ‘Thinking’ first implies cognitive change as key aspect of intervention. |
| Tellegacy: An Intergenerational Wellness and Health Promotion Project to Reduce Social Isolation and Loneliness in Older Adults: A Feasibility Study^150^ | Trained in goal setting, mindfulness, and listening. These imply a change in individual mentality, and mindfulness in particular a cognitive change. |
| The Development of LISTEN: A Novel Intervention for Loneliness^275^ | LISTEN (Loneliness Intervention using Story Theory to Enhance Nursing-sensitive outcomes). Target faulty cognitive processes - feelings of social undesirability, stigma from loneliness or chronic conditions, and negative thoughts about self in relation to others or community. |
| The Effect of a Multi-Strategy Program on Developing Social Behaviors Based on Pender’s Health Promotion Model to Prevent Loneliness of Old Women Referred to Gonabad Urban Health Centers^43^ | Improving social relationships and efficacy will improve cognitive-perception and then reduce loneliness. Implies cognitive important, but arises from relationships. Study indicates success. |
| The meanings of loneliness for older persons^87^ | Barriers included cognitive issues of cognitive sets, rejection of others, and cognitive limitations. |
| Volunteering and loneliness in older adults: A parallel mediation model^184^ | Volunteer work predicted perceived control, social self-efficacy, and lower loneliness. Perceived control and social self-efficacy mediate the relationship between volunteer activities and loneliness. Self-efficacy an individual’s perspective thus represents cognitive change. |
