## Supplementary file 2 for "Conceptualising the context and mechanisms for tackling loneliness in older adults through interventions: A Critical Interpretive Synthesis"

Supplementary file 2. Papers indicating improved social connections/relationships can elicit reduced loneliness

| **Article title** | **Reason for inclusion** |
| --- | --- |
| ‘I want him to tell me he loves me’: A smart audio device, Tochie, for resident-family connection in long-term care^154^ | Provided new opportunities for maintaining family connections. |
| “Circles of support”: social isolation, targeted assistance, and the value of “ageing in place” for older people^58^ | Helped to rebuild social networks by providing targeted support. |
| “I Didn’t Think I Needed It. But I Find I Look Forward to It Very Much”: Social Connectedness and Physical Health through the Eyes of Older Adults^120^ | Physical activity interventions that also aim to enhance social connectedness decreased loneliness. |
| A Randomized Waitlist-Controlled Trial of an Intergenerational Arts and Heritage-Based Intervention in Singapore: Project ARTISAN^148^ | Connect with someone otherwise wouldn’t have considered part of the reason for reduction of loneliness in trial. |
| A realist evaluation of loneliness interventions for older people^98^ | ‘Enhancing social connectivity’ one of the beneficial components of successful interventions. |
| A review of the impact of hearing interventions on social isolation and loneliness in older people with hearing loss^111^ | Hearing loss a barrier to forming and maintaining connections, so physical aid vital to help build and rebuild connections. |
| A systematic review of interventions for loneliness among older adults living in long-term care facilities^248^ | Laughter therapy, horticultural therapy, and reminiscence were the most effective interventions. Authors conclude are effective because they did not rely on significant physical activity or mobility, and the former 2 usually included concomitant social interactions which is argued to be a key mechanism for reducing loneliness. |
| A Systematic Review on Technology-Supported Interventions to Improve Old-Age Social Wellbeing: Loneliness, Social Isolation, and Connectedness^15^ | Review found that a lack of social relationships and infrequent contacts is the most commonly addressed factor in technology-supported interventions. |
| Addressing loneliness and isolation in retirement housing^137^ | Generating social contacts may provide a pathway to avoid loneliness, and can be done through housing design. |
| Addressing loneliness and social isolation through the involvement of primary and secondary informal caregivers in nursing homes: a scoping review^49^ | The quantity of social interactions, the perception of these encounters, and biographical changes in social relationships create loneliness. To prevent - increasing opportunities for social contact, creating meaningful encounters, maintaining existing relationships with primary informal caregivers and establishing new ones with secondary informal caregivers. |
| Addressing loneliness in later life^239^ | Recommends developing social behaviors (in group) by focusing on relationships with friends and family as an emotional source, and attendance in parties as a way to maintain social contacts. |
| Addressing Loneliness in Older People Through a Personalized Support and Community Response Program^208^ | Interventions have more chance of being effective if they resonate with people and sustain engagement |
| Aging and social isolation: An integrative review^54^ | One of the factors that results in effective interventions is to target people who can share similar experiences. |
| Associations Between Changes in Loneliness and Social Connections, and Mental Health During the COVID-19 Pandemic: The Women’s Health Initiative^136^ | ‘Disrupted social connections’ one of the factors that induce loneliness in later life. |
| Befriending Services for Culturally Diverse Older People^292^ | Supported opportunities for getting out and genuine and reciprocal relationships two factors in a positive befriending intervention. |
| Behavioural activation to mitigate the psychological impacts of COVID-19 restrictions on older people in England and Wales (BASIL+): a pragmatic randomised controlled trial^132^ | Behavioural activation sessions to maintain social connections and encourage socially reinforcing activities (so is cognitive change specifically aimed at facilitating connections). |
| Benefits of a psychosocial intervention programme using volunteers for the prevention of loneliness among older women living alone in Spain^194^ | Social participation and new bonds concluded to be among the ways the intervention reduced loneliness. |
| Beyond the Individual: Evidence Linking Neighborhood Trust and Social Isolation Among Community-Dwelling Older Adults^297^ | Perception that neighbours are trustworthy and helpful were associated with statistically significant decreases in loneliness. |
| Can digital technology enhance social connectedness among older adults? A feasibility study^50^ | Use increased perceived amount of social interaction with existing connections, but increased social connectedness was only reported by participants with geographically distant relatives. |
| Community Wise—effects and participant perceptions of a community- based -positive health intervention for older inhabitants of low SES neighbourhoods: a mixed-methods approach^243^ | ‘Belonging to a new group’ and ‘having new social contacts’ found to be important in qualitative arm of study. |
| Community-based responses to loneliness in older people: A systematic review of qualitative studies^224^ | Valuable to addressing loneliness in a specific community - new connections, belonging.  Facilitating social engagement recommended for interventions. |
| Daily Social Interactions and Momentary Loneliness: The Role of Trait Loneliness and Neuroticism^303^ | Consists of behavioral interventions that target social interactions – combination of cognitive change and social skills able to build social connections. |
| Design as Mediation for Social Connection Against Loneliness of Older People^105^ | Reciprocal partnerships argued to be important.  Facilities can mediate function, helping social interactions change from intentional to natural and from compulsory to spontaneous. |
| Digital Intergenerational Program to Reduce Loneliness and Social Isolation Among Older Adults: Realist Review^240^ | Maintaining connections a useful result of digital intervention. Do this by assisting with communication and aiding with social presence. |
| Digital Intervention in Loneliness in Older Adults: Qualitative Analysis of User Studies^268^ | Most desired functionalities that can support mutual activities and maintain or find new connections rather than enable them to share an emotional state. They were wary of the app replacing their preferred in-person social interaction. Participants also raised concerns about making the user aware of the lack of support in their social network and wanted specific means of addressing their needs. |
| Digital Storytelling as an Intervention for Older Adults: A Scoping Review^72^ | Created connections as they enjoyed it. |
| Effectiveness of interventions to address different types of vulnerabilities in community-dwelling older adults: An umbrella review^39^ | Interventions using digital technologies may decrease social isolation and loneliness as they facilitate connections. |
| Effects of selected leisure activities on preventing loneliness among older Chinese^273^ | A leisure activity steeped in a nation's culture and heritage facilitated active participation and therefore less loneliness. |
| Effects of tai chi qigong on psychosocial well-being among hidden elderly, using elderly neighborhood volunteer approach: A pilot randomized controlled trial^71^ | Social reinforcement from participation argued to have social benefits that counter social isolation and create friendships. |
| Escape loneliness by going digital: A quantitative and qualitative evaluation of a Dutch experiment in using ECT to overcome loneliness among older adults^14^ | E-mail was found to facilitate social contact.  Opportunity to develop new contacts and maintain contacts among the reasons was considered effective. Increased social connectedness and social engagement also important. |
| Evaluation of an intervention targeting loneliness and isolation for older people in North Wales^251^ | New and rekindled relationships concluded to be part of the reason the intervention was successful. |
| Examining the Effect of Contactless Intergenerational Befriending Intervention on Social Isolation Among Older Adults and Students’ Attitude Toward Companionship: Content Analysis^258^ | Intergenerational befriending, defined as perceived benefits from the friendly nature of the interaction, ability to comfortably connect with students, and positive feeling and attitude toward the student, were beneficial to loneliness. |
| Experiences of a Community-Based Digital Intervention Among Older People Living in a Low-Income Neighborhood: Qualitative Study^196^ | Valued the social interaction with volunteers. |
| Exploring existential loneliness among frail older people as a basis for an intervention: Protocol for the development phase of the LONE study^109^ | Older people highlighted a longing for a deeper connectedness among their problems. |
| Exploring the determinants and mitigating factors of loneliness among older adults^263^ | Participation in and perceived access to leisure, recreational and cultural opportunities lower the risk of loneliness in older adults. Sense of belonging to the community and especially overall sense of community. So foster social ties and community connectedness to alleviate loneliness. |
| Exploring the impact of information and communication technologies on loneliness and social isolation in community-dwelling older adults: a scoping review of reviews^139^ | Technology enabling open communication, fostering close relationships, improved social support is positive (but some technology interventions had negative effects). |
| Exploring the Impact of The NEST Collaborative’s Remote Social Intervention on Feelings of Depression and Isolation^223^ | Successfully sought to enhance social networks through meaningful friendships. |
| Family Environment, Loneliness, Hope, and Subjective Well-Being of Asian Older Adults^82^ | Concludes family environments are vital so we should bolster family environments through policy. |
| Feasibility, Acceptance, and Initial Evaluation of a Telephone-Based Program Designed to Increase Socialization in Older Veterans^166^ | Structured program with interesting topics to spend time on and the opportunity to socialize, exchange ideas, and connect. Evidence it could reduce loneliness, but individual challenges (e.g., hearing difficulty) and program-level challenges (e.g., complicated procedures) could prevent connections forming. |
| Findings From Talking Tech: A Technology Training Pilot Intervention to Reduce Loneliness and Social Isolation Among Homebound Older Adults^126^ | Can promote digital literacy and participation, which reduces loneliness, through individualised training and via specific technology. |
| Friends from the future: A scoping review of research into robots and computer agents to combat loneliness in older people^130^ | Can act as a companion, be catalyst for social interaction, facilitate remote communication with others, and remind users of upcoming social engagements. |
| Friends or frenemies? The role of social technology in the lives of older people^62^ | When social needs are not fulfilled, loneliness and social isolation can occur. Strengthens existing relationships and social structures, but depth and fun to the social contacts needed. Also a problem if stands in the way of real human contact. |
| Holistic Wellness Coaching for Older Adults: Preliminary Evidence for a Novel Wellness Intervention in Senior Living Communities^124^ | Sustained changes associated relationships formed. |
| Implementing a digital tool to support meaningful engagement with socially isolated or lonely older adults^235^ | Facilitates access to community resources, activities and people to extend or re-establish a client's social connections. |
| Increasing social connectedness for underserved older adults living with depression: A pre-post evaluation of PEARLS^266^ | Increasing opportunities for social interactions among reasons for successful reduction of loneliness. |
| Intergenerational Group Reminiscence: A Potentially Effective Intervention to Enhance Elderly Psychosocial Wellbeing and to Improve Children's Perception of Aging^127^ | Provided participants with feelings of togetherness and intimacy. |
| Interventions for alleviating loneliness among older persons: A critical review^90^ | Educational interventions focused on social networks maintenance were successful. |
| Interventions targeting loneliness and social isolation among the older people: An update systematic review^244^ | Group belonging and individual bonding common to successful interventions, but multi strategy programmes incorporating social support argued to be most effective. |
| Interventions that have potential to help older adults living with social frailty: a systematic scoping review^170^ | Social prescribing found to be effective. |
| Interventions to address social connectedness and loneliness for older adults: a scoping review^225^ | Strategies described most often were engaging in purposeful activity and maintaining contact with one’s social network. |
| Loneliness and social isolation of older adults: Why it is important to examine these social aspects together^218^ | Using a distinction between loneliness as subjective and isolation as objective state to suggests 4 types of interventions (indicates isolated and lonely people require support for isolation). |
| Loneliness and social support of older people living alone in a county of Shanghai, China^77^ | Social support may fill the need for social contacts, Potential interventions include encouraging more frequent contacts from children, the development of one-to-one 'befriending' and group activity programmes. |
| Loneliness in nursing homes: A qualitative meta-synthesis of older people's experiences^302^ | Participation and strengthening ties among the best ways to alleviate loneliness. |
| Loneliness in older persons: A theoretical model and empirical findings^91^ | One of the things interventions should target is new social contacts for lonely person. |
| Loneliness matters: a theoretical and empirical review of consequences and mechanisms^146^ | Increasing opportunities for social interaction among the vital goals for interventions. |
| Loneliness Trajectories: Evidence for Staged Intervention Among Chinese Bereaved Parents^288^ | Social network restoration important for preventing loneliness. |
| Loneliness, older people and a proposed social work response^142^ | The maintenance of meaningful existing relationships and, if required, potential introduction of new social support, are important. |
| Long-term subjective loneliness in adults after hearing loss treatment^46^ | Physical issues contribute to worsening loneliness as harder to maintain contacts. |
| Mental Health of Elders in Retirement Communities: Is Loneliness a Key Factor?^53^ | Develop gratifying interpersonal relationships to tackle loneliness. |
| Minor positive effects of health-promoting senior meetings for older community-dwelling persons on loneliness, social network, and social support^141^ | Didn’t work as lack of accessible social resources to meet participants' identified needs. I.e., can’t form connections as lack of opportunity. |
| Modifying Behavioral Activation to Reduce Social Isolation and Loneliness Among Older Adults^238^ | Modified Behavioral Activation to address social connectedness. Found a significant reduction in loneliness. |
| Multi-cultural perspectives on group singing among diverse older adults^44^ | Group singing helped loneliness. That it’s a ‘group’ important to why. |
| New Older Users’ Attitudes Toward Social Networking Sites and Loneliness: The Case of the Oldest-Old Residents in a Small Italian City^68^ | Technology mainly for contact with existing family. |
| Nutritional intervention and functional exercises improve depression, loneliness and quality of life in elderly women with sarcopenia: a randomized clinical trial^241^ | Nutrition and exercise worked, but unclear if due to concomitant social interaction. |
| Older family carers in rural areas: Experiences from using caregiver support services based on Information and Communication Technology (ICT)^55^ | Strengthening relationships with relatives and people wouldn’t otherwise access. |
| One Year Impact on Social Connectedness for Homebound Older Adults: Randomized Controlled Trial of Tele-delivered Behavioral Activation Versus Tele-delivered Friendly Visits^61^ | Help participants develop enduring skills (both resilience and social skills for forming connections). Improvements maintained at 1 year follow up. |
| Online acceptance and commitment therapy as treatment for loneliness among older adults: Report of a pilot study^301^ | Largely cognitive intervention but implies it aims to build skills for forming connections. |
| Online Social Networking and Mental Health among Older Adults: A Scoping Review^78^ | Enhanced communication with family and friends and creation of online communities among the important factors for reducing loneliness. |
| Preventing social isolation in later life: Findings and insights from a pilot Queensland intervention study^52^ | Community development approach allowed a wide range of group activities and services to create meaningful social networks and close relationships among older people in the district |
| Primary care-based interventions addressing social isolation and loneliness in older people: a scoping review^129^ | All findings assume connections as the intended pathway to reduced loneliness. |
| Promoting social capital to alleviate loneliness and improve health among older people in Spain^93^ | Increasing social capital reduces loneliness (as facilitates opportunities for meaningful connections). |
| Psychological interventions for loneliness and social isolation among older adults during medical pandemics: a systematic review and meta-analysis^192^ | Interventions targeting social skills effective. |
| Qualitative evaluation of a community-based intervention to reduce social isolation among older people in disadvantaged urban areas of Barcelona^180^ | Weekly community intervention that promotes resources among individuals and communities in order to enhance their ability to identify problems and activate solutions, encouraging participation in the community  expanding knowledge of health issues and of community activities encouraging participants to go out, giving them a feeling of being heard (loneliness not isolation?), and peer relationships, increasing participants' contacts and knowledge while the main negative features were related to repetition of certain contents.  breaking the habit of staying at home. |
| Qualitative study of loneliness in a senior housing community: the importance of wisdom and other coping strategies^215^ | Lack of social skills or abilities a cause of loneliness. Seeking companionship, and environments that enable socialising help. |
| Ringing the changes: The role of telephone communication in a helpline and befriending service targeting loneliness in older people^245^ | Ability of the services to connect disparate individuals enabled them to form different kinds of satisfying relationships. |
| Screening — An Important Starting Point for Effective Loneliness Interventions among Older Adults^160^ | Argues that increasing the contacts of an older adult can help loneliness. |
| Shared Interest Groups (SHIGs) in low-income independent living facilities^92^ | Shared interest groups are effective as point for bonding. |
| Social and physical effects of a pedometer and communication application among older men: a mixed-methods, pre/post pilot study^36^ | Parts of the concept of social connectedness are  ‘constructing reliance’, ‘remembering to consider friends’, ‘awareness of friends’ daily lives and encouragement for me’, and ‘Taking an interest in friends’ – all indicate a role for friends. |
| Social Isolation in Older Adults: A Qualitative Study on the Social Dimensions of Group Outdoor Health Walks^156^ | Fosters casual interpersonal interactions through spontaneous mixing during and after the walk. This programmatic structure counters loneliness, en-genders pleasurable anticipation of regular contact with others, supports physical activity, and fosters group cohesion. These in turn contribute to individual social wellbeing, including expanding social networks, meaningful relationships, a sense of belonging, and acting on empathy for others. |
| Social Prescription Interventions Addressing Social Isolation and Loneliness in Older Adults: Meta-Review Integrating On-the-Ground Resources^233^ | Increasing social interactions, Group-based social activities, support groups with educational elements, recreational activities, and communication technologies among most effective interventions. |
| Strategies to Promote Social Connections Among Older Adults During “Social Distancing” Restrictions^283^ | Evidence informed Connections Plan – thinking feeling doing. Constructs a need for connections as positive to overall well-being. |
| The ‘loneliness pandemic’: Implications for gerontological nursing^182^ | Maintain a relationship as important. Interactions are increasingly dependent on the use of technology. However, passive use of social networks does not contribute to a sense of belonging. |
| The Effectiveness of a Thanks, Sorry, Love, and Farewell Board Game in Older People in Taiwan: A Quasi-Experimental Study^74^ | Playing the game created improvements in scores on interpersonal communication, self-efficacy, and loneliness. Implies was an activity to bond around. |
| The link between social anxiety and intimate loneliness is stronger for older adults than for younger adults^149^ | Intervention decreased social anxiety. Older age especially a problem due to ‘pruning’ (focusing on smaller no.s of relationships). |
| The meanings of loneliness for older persons^87^ | Alone evening/weekends part of the problem though still had relationship preferences.  Barriers included social skills deficits, technological illiteracy, and physical, sensory, and cognitive limitations, economic hardship, and community programming which failed to promote socialization. |
| The role of assistive technology in addressing social isolation, loneliness and health inequities among older adults during the COVID-19 pandemic^168^ | Animal therapy can reduce social isolation and therefore loneliness too. |
| The Social Bridging Project: Intergenerational Phone-Based Connections With Older Adults During the COVID-19 Pandemic^221^ | Reduce the impact of social isolation on older adults, assist them in acquiring technology skills, assist accessing telehealth community supports all help reduce loneliness. |
| The value of social eating at culturally and linguistically diverse lunch clubs: a descriptive study^210^ | A rare opportunity for social interaction and helps maintain ties to culture in a foreign country. |
| Virtual Intergenerational Reverse-Mentoring Program Reduces Loneliness among Older Adults: Results from a Pilot Evaluation^167^ | Social capital and intergroup contact theorised as important. Learning new ways to interact with friends and family concluded as helpful. |
