## Supplementary file 3 for "Conceptualising the context and mechanisms for tackling loneliness in older adults through interventions: A Critical Interpretive Synthesis"

Supplementary file 3. Papers indicating changed physiological condition can elicit reduced loneliness

| **Article title** | **Reason for inclusion** |
| --- | --- |
| A peer intervention reduces loneliness and improves social well-being in low-income older adults: A mixed-methods study^176^ | Improved mood reported as a cause of less loneliness arsing from the intervention. |
| A Randomized Waitlist-Controlled Trial of an Intergenerational Arts and Heritage-Based Intervention in Singapore: Project ARTISAN^148^ | Enjoyment of intervention as part of what reduced loneliness. |
| A social robot intervention on depression, loneliness, and quality of life for Taiwanese older adults in long-term care^75^ | Companionship with the robot (not people) able to help – indicates something in the individual rather than their perception of their social relationships. |
| A systematic review of interventions for loneliness among older adults living in long-term care facilities^248^ | Laughter therapy able to reduce loneliness (implicitly due to impact on mood). |
| A Systematic Review of Research on Pet Ownership and Animal Interactions among Older Adults^131^ | Included as pets can help even though not a connection with a person (though arguably connection to an animal can count as a social connection). |
| Ageing, Leisure, and Social Connectedness: How could Leisure Help Reduce Social Isolation of Older People?^278^ | Leisure activities explain a significant part of older people's social connectedness, but not when passive (eg computer use), suggesting the physical activity is important. |
| Alleviating Loneliness in Older Adults Through Creative Expression^220^ | Creative arts engagement is enabled by modulation of brain activity, and reinforced by personalized and coordinated psychological and social features. |
| Associations Between Changes in Loneliness and Social Connections, and Mental Health During the COVID-19 Pandemic: The Women’s Health Initiative^136^ | Engaging in more frequent physical activity identified as reducing loneliness. |
| Bonding With Bot: User Feedback on a Chatbot for Social Isolation^106^ | Not a social connection, but still able to help. An be interpreted as a neuropsychological benefit related to physical act of speaking. |
| ChatGPT: A Companion for Dementia Care^152^ | Not a social connection, but still able to help - Offers a way to talk which may increase psychoneurological stimulation. |
| Context of walking and loneliness among community-dwelling older adults: a cross-sectional study^214^ | Walking with others = less loneliness, walking alone no significant difference to not walking (though direction still positive, suggesting may help a bit but needs more research). |
| Dance Intervention Affects Social Connections and Body Appreciation Among Older Adults in the Long Term Despite COVID-19 Social Isolation: A Mixed Methods Pilot Study^15^ | Increased connection partly through a shift to experiencing the body as responsive. |
| Development and evaluation of the information and communication technology-based Loneliness Alleviation Program for community-dwelling older adults: A pilot study and randomized controlled trial^40^ | Psychological interventions - enhance positive emotions, such as feelings of comfort and happiness, thereby alleviating loneliness. As can be enjoyed, loneliness reduced, willingness to participate increased. |
| Digital Storytelling as an Intervention for Older Adults: A Scoping Review^72^ | Created connections as they enjoyed it. |
| Dog-assisted interventions and outcomes for older adults in residential long-term care facilities: A systematic review and meta-analysis^157^ | Supplemented missing interaction as therapeutic including touch and ‘spirituality’. |
| Effect of Combining Adapted Physical and Artistic Activities on Feeling of Loneliness and Aggression in the Elderly Living in Nursing Homes^259^ | Just taking part in activity part of the benefit to loneliness (regardless of social relationships). |
| Effectiveness of interventions to address different types of vulnerabilities in community-dwelling older adults: An umbrella review^39^ | Physical activity may improve social functioning and interventions using digital technologies may decrease social isolation and loneliness. |
| Effectiveness of LISTEN on loneliness, neuroimmunological stress response, psychosocial functioning, quality of life, and physical health measures of chronic illness^276^ | Cognitive change significantly improved loneliness and physiological markers of loneliness (using psychoneuroimmunology paradigm). |
| Effects of Video-Assisted Leisure Education on Leisure, Loneliness, and Affect of Older Adults^96^ | Leisure creates opportunity for connection and thus less loneliness. Impact on amount of leisure and loneliness small, but increased knowledge reduced overall negative effects. |
| Establishing routines to cope with the loneliness associated with widowhood: a narrative analysis^97^ | Routines can alleviate loneliness as everyday chit chat that just happens (but aren’t thought of as social relationships, so arguable that main impact is stimulation through social interaction). |
| Evaluation of an intervention targeting loneliness and isolation for older people in North Wales^251^ | Enjoyment, happiness, and better health part of efficacy of programme. |
| Examining the Effect of Contactless Intergenerational Befriending Intervention on Social Isolation Among Older Adults and Students’ Attitude Toward Companionship: Content Analysis^258^ | Reduced boredom and greater level of happiness as part of overall cause of reduction in loneliness. |
| Friends from the future: A scoping review of research into robots and computer agents to combat loneliness in older people^130^ | Can act as a direct companion, but as it isn’t a person cannot impact loneliness defined as a subjective lack of relationships. |
| Loneliness in nursing homes: A qualitative meta-synthesis of older people's experiences^302^ | Diverting attention constructed as a cause of reduced loneliness. Implies can have real change to loneliness through focus on an activity. |
| Loneliness in Old Age: Interventions to Curb Loneliness in Long-Term Care Facilities^59^ | Increase activity and social interaction – more interaction was thing that helped, but activity an important anchor to do it. |
| Loneliness matters: a theoretical and empirical review of consequences and mechanisms^146^ | Argues loneliness a regulatory loop consisting of cognitive, behavioral, and physiological cycles. |
| Managing loneliness: a qualitative study of older people’s views^171^ | Physical engagement with the world beyond the home and distraction can be used to alleviate loneliness. Among other things, people use interests and hobbies and alcohol, which indicate a physiological component. |
| Mechanisms through which befriending services may impact the health of older adults: A dyadic qualitative investigation^143^ | Improving mood, getting cognitive (neuropsychological) stimulation and novelty (something different and interesting to pique interest) among the findings on what can be helpful in interventions. |
| My Precious Friend: Human-Robot Interactions in Home Care for Socially Isolated Older Adults^186^ | Pleasure in playing with Hyodol reduced loneliness. Acted as a surrogate family member or friend, but not able to impact perception of social relationships. |
| Nutritional intervention and functional exercises improve depression, loneliness and quality of life in elderly women with sarcopenia: a randomized clinical trial^241^ | Nutrition and exercise worked, but unclear if due to physiological change or concomitant social interaction. |
| Online Social Networking and Mental Health among Older Adults: A Scoping Review^78^ | If social media had positive associations with well-being and life satisfaction was more successful. |
| Participant and Public Involvement in Refining a Peer-Volunteering Active Aging Intervention: Project ACE (Active, Connected, Engaged)^295^ | Intervention helped some as it was ‘something to do’ that could facilitate ‘enjoyment’. |
| Physical impairments disrupt the association between physical activity and loneliness: A longitudinal study^57^ | Improvements in moderate to vigorous PA were associated with decreases in loneliness (but impairments prevented this relationship) |
| Pilot study of Qigong/Tai Chi Easy acute effects of meditative movement, breath focus and “flow” on blood pressure, mood and oxytocin in older adults^182^ | Found an unexpected positive association between oxytocin and loneliness. Study implies intervention impacted oxytocin and loneliness, but complicated as oxytocin increased for baseline not lonely, and decreased in baseline lonely. |
| Reducing Loneliness Among Aging Adults: The Roles of Personal Voice Assistants and Anthropomorphic Interactions^165^ | Found a successful reduction in loneliness despite no attempt to impact subjective perspective of social relationships or meaning in life. |
| Regional brain volumes moderate, but do not mediate, the effects of group-based exercise training on reductions in loneliness in older adults^110^ | Individuals with larger baseline amygdalae experienced greater decreases in loneliness due to greater reductions in stress. |
| Social Isolation in Older Adults: A Qualitative Study on the Social Dimensions of Group Outdoor Health Walks^156^ | Fosters casual interpersonal interactions, en-genders pleasurable anticipation, and supports physical activity. |
| The Impact of Interaction Time and Verbal Engagement with Personal Voice Assistants on Alleviating Loneliness among Older Adults: An Exploratory Study^296^ | The act of speaking, even with a PVA, can reduced loneliness, suggesting a neuropsychological benefit not related to subjective perspective of social relationships or meaning in life. |
| The preliminary effects of laughter therapy on loneliness and death anxiety among older adults living in nursing homes: A nonrandomised pilot study^178^ | Facilitating positive emotions through games and fake laughter reduced loneliness. |
| The role of oxytocin in regulating loneliness in old age^38^ | Good interactions created oxytocin response and less loneliness. Indicates social interactions are a mediating factor in a neuropsychological process we describe as ‘loneliness’. |
| The role of ChatGPT in mitigating loneliness among older adults: An exploratory study^207^ | Users established an emotional connection with ChatGPT, thus potential to provide comfort and companionship. Again may signify that chance to speak reduces loneliness regardless of subjective perspective of social relationships or meaning in life. |
| The role of pet attachment in alleviating the negative effects of loneliness on a health-promoting lifestyle: An empirical study based on threshold effects for pet owners^195^ | Can compensate for a lack of social relationships, but can’t impact of subjective perspective of social relationships or meaning in life directly. However, animals may be able to constitute a social connection. |
| The Role of Solitary Activity in Moderating the Association between Social Isolation and Perceived Loneliness among U.S. Older Adults^188^ | Solitary activities can decrease loneliness. |
| Understanding and Addressing Older Adults’ Loneliness: The Social Relationship Expectations Framework^42^ | Need for enjoyment highlighted. |
| User-Friendly Chatbot to Mitigate the Psychological Stress of Older Adults During the COVID-19 Pandemic: Development and Usability Study^83^ | Companionship from older adult designed chatbox may signify that chance to speak reduces loneliness regardless of subjective perspective of social relationships or meaning in life. Didn’t work in younger people who, though they may be lonely, generally have more chances to interact with others. |
