## Supplementary file 4 for "Conceptualising the context and mechanisms for tackling loneliness in older adults through interventions: A Critical Interpretive Synthesis"

Supplementary file 4. Papers indicating a more meaningful life can elicit reduced loneliness

| **Article title** | **Reason for inclusion** |
| --- | --- |
| “It makes me feel not so alone”: features of the Choose to Move physical activity intervention that reduce loneliness in older adults^121^ | ‘Opportunity to share information and experiences and learn from others’ identified as a positive feature of interventions. |
| 'A lonely old man': Empirical investigations of older men and loneliness, and the ramifications for policy and practice^8^ | Facilitate the construction of a 'proud' masculine identity (purpose, helping out), perceived social worth, and reducing shame and stigma all highlighted as useful for reducing loneliness. |
| A Systematic Review of Research on Pet Ownership and Animal Interactions among Older Adults^131^ | Opportunity to nurture and care for another being highlighted as a possible reason for efficacy of pet ownership in reducing loneliness. |
| Addressing loneliness in later life^239^ | Spending time in activities constructed as ‘constructive’, such as reading and gardening, can be helpful. |
| Aging and Feeling Valued Versus Expendable During the COVID-19 Pandemic and Beyond: a Review and Commentary of Why Mattering Is Fundamental to the Health and Well-Being of Older Adults^118^ | A feeling of ‘mattering’ as key to not lonely. |
| Associations Between Changes in Loneliness and Social Connections, and Mental Health During the COVID-19 Pandemic: The Women’s Health Initiative^136^ | Having a higher purpose in life associated with less loneliness. |
| Becoming immune to loneliness helping the elderly fill a void^48^ | Three relevant arguments by author:  1. Persons immune to loneliness have characteristics that permit growth and constructive behaviors when faced with a loss or change.  2. Loneliness in old age occurs because interactions and communications from others are ignored and are not accepted by others.  3. Knowledge of an individual's level of creative potential is relevant for nurses planning interventions for mobilizing a support system for the lonely elderly. |
| Being disconnected from life: meanings of existential loneliness as narrated by frail older people^262^ | Being met with indifference and lacking purpose and meaning can increase loneliness. Interventions should develop strategies to support meaning in life. |
| Design as Mediation for Social Connection Against Loneliness of Older People^105^ | Generative role for a sense of accomplishment as important for reducing loneliness. |
| Does Becoming A Volunteer Attenuate Loneliness among Recently Widowed Older Adults?^67^ | Volunteering 2+ hours a week related to purpose thus less loneliness. |
| Effects of horticultural therapy on psychosocial health in older nursing home residents: A preliminary study^76^ | A sense of achievement identified as able to reduce loneliness. |
| Facing the Next “Geriatric Giant”—A Systematic Literature Review and Meta-Analysis of Interventions Tackling Loneliness and Social Isolation Among Older Adults^107^ | Intervention involving ‘transferring knowledge and skills’ by lonely person to another can reduce loneliness. |
| Group dynamics in older people's closed groups: Findings from Finnish psychosocial group rehabilitation for lonely older people^242^ | Group self-esteem argued to be important for reducing loneliness. |
| Holistic Wellness Coaching for Older Adults: Preliminary Evidence for a Novel Wellness Intervention in Senior Living Communities^124^ | Sustained changes to loneliness associated with goal attainment. |
| Impact of an intergenerational program to improve loneliness and social isolation in older adults initiated at the time of emergency department discharge: study protocol for a three-arm randomized clinical trial^304^ | Generativity - the interest in establishing and guiding the next generation – good for loneliness interventions as it provides purpose and meaning in interactions. |
| Intergroup ‘skype’ quiz sessions in care homes to reduce loneliness and social isolation in older people^300^ | Re-gaining sense of self and purpose can occurred in these sessions and thus reduced loneliness. |
| Interventions against social isolation of older adults: A systematic review of existing literature and interventions^202^ | Targeted, tailored person-centred interventions also effective as empowers and motivate people, and participants able to control loneliness better. |
| Interventions aimed at Alleviating Loneliness and Social Isolation among the Older Population: Perspectives of Service Providers^114^ | Empowerment of service users and staff/volunteers among the most helpful factors. |
| Interventions targeting loneliness and social isolation among the older people: An update systematic review^244^ | Most effective factors - group belonging, individual bonding, and empowerment. |
| It Gave Me Somebody Else to Think About Besides Myself: Caring Callers Volunteer Experiences With a Telephone-Based Reassurance Program for Socially Isolated Older Adults^116^ | Purposeful use of time and learning new skills were reasons for the successful components of the intervention. |
| Loneliness in Men 60 Years and Over: The Association With Purpose in Life^217^ | Lower purpose in life is an indicator of loneliness. A multidimensional approach to any intervention needs to occur as purpose is a relative concept. |
| Loneliness, Purpose in Life, and Protective Behaviors: Examining Cross-Sectional and Longitudinal Relationships in Older Adults before and during COVID-19^197^ | Purpose in life fully mediated the negative impact of loneliness on protective behaviours. |
| Participant and Public Involvement in Refining a Peer-Volunteering Active Aging Intervention: Project ACE (Active, Connected, Engaged)^297^ | Altruism a motive to get involved in something that can alleviate loneliness. |
| Promoting the empowerment and emancipation of community-dwelling older adults with chronic multimorbidity through a home visiting programme: a hermeneutical study^103^ | Personalising interventions via trusting relationships with home visitors empowers people thus alleviates loneliness. Implies cognitive change within individual but use of terms empowerment and emancipation also suggest a social impact. |
| Social Isolation and Psychological Distress Among Older Adults Related to COVID-19: A Narrative Review of Remotely-Delivered Interventions and Recommendations^133^ | Interventions that promote active engagement and are goal-directed are part of the best type of interventions. |
| Social Isolation in Older Adults: A Qualitative Study on the Social Dimensions of Group Outdoor Health Walks^156^ | Acting on empathy for others purported to be part of why the walks reduced loneliness. |
| Solitude in old age: a scoping review of conceptualisations, associated factors and impacts^228^ | Destigmatizing time spent alone can empower older adults to choose fulfilling activities and connect with themselves, thus reducing loneliness. |
| Student-senior isolation prevention partnership: a Canada-wide programme to mitigate social exclusion during the COVID-19 pandemic^216^ | Community partnership model - empowerment, behaviour, and organization are key factors for success. |
| The effectiveness of group reminiscence therapy for loneliness, anxiety and depression in older adults in long-term care: A systematic review^271^ | The gain in personal value and self-identity are key to success of reminiscence therapy. |
| The Impact of Robotic Companion Pets on Depression and Loneliness for Older Adults with Dementia During the COVID-19 Pandemic^119^ | Qualitative data suggests looking after the robot pet a purposeful thing to do, but unclear whether this can be permanent. |
| The Leveraging Exercise to Age in Place (LEAP) Study: Engaging Older Adults in Community-Based Exercise Classes to Impact Loneliness and Social Isolation^206^ | Purpose-driven activity designed to promote physical activity and social interaction. Successful reduction in loneliness. |
| The Management of Loneliness in Aged Care Residents: An Important Therapeutic Target for Gerontological Nursing^60^ | Article argues human beings have an intrinsic need for contributing to family, social, community, and institutional life, which preserves the feeling of being valued and helpful, and still serving a purpose. This means interventions like reminiscence and gardening can be effective regardless of social connections formed. |
| The Role of Solitary Activity in Moderating the Association between Social Isolation and Perceived Loneliness among U.S. Older Adults^188^ | Solitary activities can decrease loneliness. |
| Understanding and Addressing Older Adults’ Loneliness: The Social Relationship Expectations Framework^42^ | Needs from relationships include generativity and contribution, and being respected and valued. |
| Verification of the Effectiveness of a Communication Application in Improving Social Connectedness and Physical Health among Unacquainted Older Men: A Mixed-Methods Pilot Study^261^ | Measuring step count as part of the intervention added purpose increasing efficacy, continuation, and subsequent reduction of loneliness. |
| Volunteering and loneliness in older adults: A parallel mediation model^184^ | Volunteer work predicted perceived control, social self-efficacy, and lower loneliness. Perceived control and social self-efficacy mediate the relationship between volunteer activities and loneliness. |
