## Supplementary file 5 for "Conceptualising the context and mechanisms for tackling loneliness in older adults through interventions: A Critical Interpretive Synthesis"

Supplementary file 5. Articles indicating a need to consider meso and macro perspectives in loneliness interventions

| **Article title** | **Reason for inclusion** |
| --- | --- |
| “Circles of support”: social isolation, targeted assistance, and the value of “ageing in place” for older people^58^ | “Ageing in place” (i.e., at home) can result in social isolation as people lose capacity to maintain connections. |
| “I Didn’t Think I Needed It. But I Find I Look Forward to It Very Much”: Social Connectedness and Physical Health through the Eyes of Older Adults^120^ | The natural environment and transportation can be factors in whether someone is lonely. |
| A mixed methods feasibility study of a virtual group-based social support program for older adults in residential care^211^ | staffing, resources, and internet accessibility are important to tackling loneliness. |
| A systematic scoping review of community-based interventions for the prevention of mental ill-health and the promotion of mental health in older adults in the UK^185^ | ‘System approaches’ – those involving structural and interpersonal factors - identified as a potential type of intervention. Socioeconomic determinants of poor mental health were largely absent from research though. |
| Aging-in-place^249^ | Ageing in place, particularly in rural areas, can result in social isolation. |
| Decreasing Loneliness and Social Disconnectedness among Community-Dwelling Older Adults: The Potential of Information and Communication Technologies and Ride-Hailing Services^272^ | Possibility that transport services can connect to people/places thus less loneliness. |
| Design as Mediation for Social Connection Against Loneliness of Older People^105^ | Notion of design as mediating function emphasises role of physical environment. |
| Does household help prevent loneliness among the elderly? An evaluation of a policy reform in the Netherlands^47^ | Couldn’t find evidence housing policy impacted loneliness. May be too complex to find at macro level. |
| Effects of Video-Assisted Leisure Education on Leisure, Loneliness, and Affect of Older Adults^96^ | Implies need for leisure infrastructure, although this study didn’t find a successful reduction in loneliness. |
| Effectiveness of a community-based integrated service model for older adults living alone: A nonrandomized prospective study^222^ | Community-based integrated model for ageing in place decreased loneliness scores (need for infrastructure to do so in community). |
| Evaluation of an intervention targeting loneliness and isolation for older people in North Wales^251^ | Good public transport a useful factor in tackling loneliness. |
| Experiences of a Community-Based Digital Intervention Among Older People Living in a Low-Income Neighborhood: Qualitative Study^196^ | Study noted need for financial comfort to purchase a phone to then take part in their intervention. |
| Exploring the determinants and mitigating factors of loneliness among older adults^263^ | Recreational and cultural opportunities lower the risk of loneliness in older adults. |
| Has the UK Campaign to End Loneliness Reduced Loneliness and Improved Mental Health in Older Age? A Difference-in-Differences Design^190^ | Found no evidence authorities implementing gold standard antiloneliness strategies had a better impact on loneliness than other authorities. Suggests could be too much complexity and lack of meaningful difference between comparison groups. |
| Housing Unit Type and Perceived Social Isolation Among Senior Housing Community Residents^66^ | Consider housing type as part of loneliness policy – physical environment can impact loneliness. |
| Implementing a digital tool to support meaningful engagement with socially isolated or lonely older adults^235^ | Time and resources for intervention providers were found to be important for success. |
| Interventions Aimed at Alleviating Loneliness and Social Isolation among the Older Population: Perspectives of Service Providers^114^ | Characteristics of the wider community that impacted intervention were where services were locate; education and awareness of services; family and community structures; and accessible, affordable, and safe public transport |
| Loneliness and social support of older people living alone in a county of Shanghai, China^77^ | Living alone may increase the risk of loneliness of older people, especially for those in China where collectivism and filial piety are emphasised. |
| Loneliness in Older Migrants: Exploring the Role of Cultural Differences in Their Loneliness Experience^229^ | No difference between individualism vs collectivism in odds of loneliness but social capital did impact odds. |
| Loneliness in older persons: A theoretical model and empirical findings^91^ | Proposes that loneliness is interaction of cognitive processes and environmental events. Considering mobility and financial resources important for reducing loneliness. |
| Lonely places or lonely people? Investigating the relationship between loneliness and place of residence^286^ | Finds deprivation and loneliness are linked. |
| Looking before we leap: Building the evidence for social prescribing for lonely older adults^256^ | Problem of 'road to nowhere' referrals - investments in social prescribing must also be accompanied by investments in communities. |
| Mastery and marital processes: Mechanisms linking midlife economic adversity and later-life loneliness for husbands and wives in enduring marriages^291^ | Midlife economic problems predicts later life loneliness. |
| Minor positive effects of health-promoting senior meetings for older community-dwelling persons on loneliness, social network, and social support^141^ | Authors believe intervention didn’t work for loneliness as there was a lack of accessible social resources to meet participants' identified needs. |
| Participant and Public Involvement in Refining a Peer-Volunteering Active Aging Intervention: Project ACE (Active, Connected, Engaged)^295^ | Childcare commitments and high volunteering workload barriers to positive interventions. |
| Preventing social isolation in older people^94^ | Argues there is a need for a cultural change from ‘cure’ to ‘prevention’. |
| Promoting social capital to alleviate loneliness and improve health among older people in Spain^93^ | Increasing social capital reduces loneliness. |
| Qualitative study of loneliness in a senior housing community: the importance of wisdom and other coping strategies^215^ | Finds evidence environment enables socialising. |
| Redesigning Memory Care in the COVID-19 Era: Interdisciplinary Spatial Design Interventions to Minimize Social Isolation in Older Adults^115^ | Finds evidence can minimise isolation through spatial design. |
| Social environment support to overcome loneliness among older adults: A scoping review^250^ | Two of the dimensions they find important to loneliness: Neighbours – helps in emergencies, close geography. Government – sets up society and programmes. |
| Social-ecological factors influencing loneliness and social isolation in older people: a scoping review^209^ | Three societal (macro) -level interventions were found: two campaigns to reduce ageism and one which explored the impact of free public transport. Community-based interventions (meso) were either educational or enlisted volunteers to foster connections. |
| Subjective social isolation or loneliness in older adults residing in social housing in Ontario: a cross-sectional study^41^ | Structural barriers found prevent engagement in social activities or maintenance of social support. |
| Tackling social disconnection: an umbrella review of RCT-based interventions targeting social isolation and loneliness^144^ | Finds a need for preventive structural interventions. |
| Technological interventions to reduce loneliness and social isolation among community-living older adults: A scoping review^294^ | Accessibility and technology literacy were found to be barriers to the success of an intervention. |
| The effect of participation in support groups on retirement syndrome in older adults^247^ | ‘Retirement syndrome’ argued to be a cause of loneliness, so provided a support group aimed at this. Successfully reduced loneliness. |
| The role of socio-economic status and neighborhood social capital on loneliness among older adults: evidence from the Sant Boi Aging Study^104^ | Neighbourhood social capital found to have a greater impact on loneliness than individual social capital, suggesting social structures need changing more than individuals. |
