## Supplementary file 6 for "Conceptualising the context and mechanisms for tackling loneliness in older adults through interventions: A Critical Interpretive Synthesis"

Supplementary file 6. Articles relaying practical ways to provide a successful intervention

| **Article title** | **Reason for inclusion (refers to all practical recommends written in blue in figure 2).** |
| --- | --- |
| “Circles of support”: social isolation, targeted assistance, and the value of “ageing in place” for older people^58^ | Recommends practical interventions, e.g. help to attend appointments and financial advice. Requires a well-run organisation. |
| “I Didn’t Think I Needed It. But I Find I Look Forward to It Very Much”: Social Connectedness and Physical Health through the Eyes of Older Adults^120^ | Looking forward to activity implies a need for reliable routines. |
| “It makes me feel not so alone”: features of the Choose to Move physical activity intervention that reduce loneliness in older adults^121^ | Good features highlighted included engagement with others who share similar/familiar experiences. |
| 'A lonely old man': Empirical investigations of older men and loneliness, and the ramifications for policy and practice^8^ | Gender appropriate activities, which partly constituted shared interests/activities, were recommended as useful. |
| A mixed methods feasibility study of a virtual group-based social support program for older adults in residential care^211^ | Sufficient and well-trained staff, resources, and internet accessibility noted as helpful. |
| A peer intervention reduces loneliness and improves social well-being in low-income older adults: A mixed-methods study^176^ | Matching of characteristics of peers to participants is useful, especially when resulted in participants reporting strong feelings of kinship. Shared identity a way to bond. |
| A Randomized Waitlist-Controlled Trial of an Intergenerational Arts and Heritage-Based Intervention in Singapore: Project ARTISAN^148^ | Shared nationhood (identity) key to projects success. |
| A realist evaluation of loneliness interventions for older people^98^ | Recurring interventions useful as provide a routine. Personalised approaches necessary as individuals have specific needs. |
| A systematic review of interventions for loneliness among older adults living in long-term care facilities^248^ | Laughter therapy, horticultural therapy, and reminiscence were most effective because they do not rely on significant physical activity or mobility (so build on strengths). |
| A Technology Training Program to Alleviate Social Isolation and Loneliness Among Homebound Older Adults: A Community Case Study^164^ | Lauds having an existing relationship with clientele, an established infrastructure to deliver community-based interventions, the alignment of intervention goals with broader organizational aims, and highlights a need for funding to support dedicated program staff (so consists of community strengths approach and well-run organisation). |
| Addressing loneliness and isolation in retirement housing^137^ | Individualising services according to why a person is lonely necessary. |
| Addressing loneliness in later life^239^ | Highlights engaging in eating and drinking rituals in a routinised way as a means of maintaining social contacts. |
| Addressing Loneliness in Older People Through a Personalized Support and Community Response Program^208^ | Highlights: interventions have more chance of being effective if they resonate with and sustain engagement (partly through shared identity); a need for multiple pathways to entry (support to attend); the usefulness of co-creation; a need for flexibility in form, regular feedback and volunteer support (i.e., being a well-run and adaptable organisation). |
| Adequacy of Web-Based Activities as a Substitute for In-Person Activities for Older Persons during the COVID-19 Pandemic: Survey Study^88^ | Highlights routine as important for tackling loneliness. |
| Aesthetically Designing Video-Call Technology With Care Home Residents: A Focus Group Study^299^ | Personalization and a need to consider the for socialization versus fear of socialization (implies a need for support to attend intervention). |
| Aging and social isolation: An integrative review^54^ | Interventions that are most effective utilize existing resources in the community, are tailor-made according to the needs of the individual, and target people who can share similar experiences. |
| Association between loneliness and acceptance of using robots and pets as companions among older Chinese immigrants during the COVID-19 pandemic^79^ | Interventions tailored for older people with specific cultural requirements to address loneliness are needed. |
| Attitudes of Israelis toward family caregivers assisted by a robot in the delivery of care to older people: The roles of collectivism and individualism^37^ | Cultural acceptance of robot increased likeliness of efficacy (so cultural awareness is important, but not always in immediately obvious ways). |
| Attributions of Loneliness—Life Story Interviews with Older Mental Health Service Users^65^ | Refers to ‘personalised’ as indicates a need to consider specific situation of person. |
| Attuning to the needs of structural socially isolated older adults with complex problems: the experiences of social workers with personal guidance trajectories for a less-researched group^198^ | Suggests interventions need a personalised approach with reasonable and practical problem solving. |
| Befriending Services for Culturally Diverse Older People^292^ | Services should ensure: reliability (routine); positive befriender characteristics; connect people to their culture. Are most effective when relationships with staff/volunteers went beyond professional. |
| Being cut off from social identity resources has shaped loneliness during the coronavirus pandemic: A longitudinal interview study with medically vulnerable older adults from the United Kingdom^145^ | Social identification (shared identity) with other attendees is the important characteristic for efficacy. |
| Can Digital Technology Enhance Social Connectedness Among Older Adults? A Feasibility Study^50^ | Valuing of the traditional community defined by geographical place helped facilitate a successful use of technology. Created some feelings of inadequacy and awareness of frailty, suggesting technology focusing on what the person can’t do rather than strengths is a problem. |
| Change and stability in loneliness and friendship after an intervention for older women^205^ | Recovery from loneliness after a year was associated with the presence of a friend in the outer circle of the convoy and having more variation in one's friendships initially and one year later (so highlights need for friendship outside of immediate intervention activities). |
| Community-based responses to loneliness in older people: A systematic review of qualitative studies^224^ | It is valuable to establish a user-led and tailored setting that facilitates social engagement. |
| Conditions for Feasibility of a Multicomponent Intervention to Reduce Social Isolation and Loneliness in Noninstitutionalized Older Adults^147^ | Positive features: a specialized intervention agent (to support to attend and ensure a well-run organisation), a clear intervention purpose, facilitating active participation, adequate management. |
| Consensus statement: loneliness in older adults, the 21st century social determinant of health?^246^ | Recommends interventions are tailored and matched to the specific root causes of loneliness in the individual. |
| Delivering a psychosocial program for older people living in retirement homes during the Covid-19 pandemic: A process evaluation and recommendations for community interventions^162^ | Active participation is good, but only if co-produced too (because of power dynamics not just general preferences). |
| Determinants of Implementing an Information and Communication Technology Tool for Social Interaction Among Older People: Qualitative Content Analysis of Social Services Personnel Perspectives^122^ | Determinants of successful intervention all represented a well-run intervention: good organizational structure; leadership; personnel skills and attitudes; resources; accessibility; legal liability. |
| Developing A Digital Psychoeducational Tool to Reduce Loneliness in Older Adults: A Design Case Study^201^ | Determinants of a successful intervention: tone towards participants, relatability (shared identity), accessibility, readability, engagement, and trustworthiness. |
| Dog-assisted interventions and outcomes for older adults in residential long-term care facilities: A systematic review and meta-analysis^157^ | Structure and flexibility of staff and organisation vital |
| Effect of Combining Adapted Physical and Artistic Activities on Feeling of Loneliness and Aggression in the Elderly Living in Nursing Homes^259^ | Active participation in art-based group therapy creates an interactive and encouraging environment. |
| Effectiveness of a community-based integrated service model for older adults living alone: A nonrandomized prospective study^222^ | Community-based integrated model for ageing in place decreased loneliness scores (strengths of community drawn on). |
| Effectiveness of chronic disease self-management education (CDSME) programs to reduce loneliness^265^ | May reduce loneliness because of its highly interactive and process-driven format (active participation) and ability to create bonds through shared experiences. |
| Effects of a ‘social activity program that encourages interaction’ on rural older people's psychosocial health: Mixed-methods research^173^ | Themes ‘stimulation brought about by relationships with peers,’ ‘realization as to where they feel they belong,’ ‘rethinking of oneself in the community,’ and ‘awareness of attachment to and coexistence with the community’ all represent shared interests and identity. |
| Effects of a program to prevent social isolation on loneliness, depression, and subjective well-being of older adults: A randomized trial among older migrants in Japan^254^ | Effective to utilize existing community resources, tailor-make services according to the specific needs of the individual, and target people who can share similar experiences. |
| Effects of a social internet-based intervention programme for older adults: An explorative randomised crossover study^183^ | Restraints and changes to social activities contribute to loneliness (lost routines). |
| Effects of selected leisure activities on preventing loneliness among older Chinese^273^ | A leisure activity steeped in ‘culture and heritage’ is useful. Represents shared identity and cultural awareness. |
| Efficacy of group reminiscence therapy based on Chinese traditional festival activities (CTFA-GRT) on loneliness and perceived stress of rural older adults living alone in China: a randomized controlled trial^189^ | Community and tradition linked approach worked (cultural awareness). |
| Efficacy of the I-SOCIAL intervention for loneliness in old age: Lessons from a randomized controlled trial^89^ | Intervention worked as individualised treatment options. Passive activities were not suitable (active participation needed). |
| Engage coaching for caregivers: a pilot trial to reduce loneliness in dementia caregivers^282^ | Personalized intervention targets were helpful. |
| Establishing routines to cope with the loneliness associated with widowhood: a narrative analysis^97^ | Routines can alleviate loneliness as they provide everyday chit chat that just happens. |
| Evaluating Feasibility, Value and Characteristics of an Intergenerational Friendly Telephone Visit Program During the Covid-19 Pandemic^177^ | Mutuality (often representing shared identity/interest) and routine among the determinants of success. |
| Evaluation of a culturally adapted reminiscence therapy intervention: Improving mood, family and community connectedness in Spanish- and Vietnamese-speaking older adults^102^ | Culturally adapting service worked well, suggesting importance of cultural awareness. |
| Evaluation of an intervention targeting loneliness and isolation for older people in North Wales^251^ | Service was good as it was something to look forward to (routine) and the volunteers were excellent (suggesting well-run organisation). |
| Evaluation of an online arts-based platform to support the health and well-being of older adults during the COVID-19 pandemic: a cross-sectional survey^270^ | Intervention may have been good because it was well-organized and convenient (the latter implying a routine). |
| Experiences of a Community-Based Digital Intervention Among Older People Living in a Low-Income Neighborhood: Qualitative Study^196^ | Findings valued the personalized learning model and suggest a need to recognise the cultural situation of older adults in design of digital interventions (co-produce to ensure culturally aware). |
| Exploring the determinants and mitigating factors of loneliness among older adults^263^ | Sense of belonging to the community was good (shared identity). |
| Exploring the Impact of The NEST Collaborative’s Remote Social Intervention on Feelings of Depression and Isolation^223^ | one-to-one empathy-based phone calls were good, implying a shared identity/experience was useful. |
| Faith Community Nursing: Identifying and Combating Social Isolation and Loneliness in Older Adults^193^ | Faith as an identity and community to build from. |
| Feasibility, acceptability, and preliminary efficacy of an internet-based CBT intervention for loneliness in older adults: A pilot RCT^108^ | Individualisation and interactivity needed for effective cognitive change. Negatives - Individual challenges (not built on individual strengths) and program-level challenges (not well run). |
| Friendly Visiting Programs for Older People Experiencing Social Isolation: A Realist Review of what Works, for whom, and under what Conditions^289^ | New relationships form best if client volunteer match, and are trained, supported, supervised (well run). |
| Group dynamics in older people's closed groups: Findings from finnish psychosocial group rehabilitation for lonely older people^242^ | Interventions should account for group dynamics, including considering whether are shared identities and responsibility for tasks (well run). |
| Implementation of Social Isolation Screening and an Integrated Community Resource Referral Platform^138^ | Study implies people need a clear and supported pathway from referral (support to attend). |
| Integrating community participation in the transition of older adults from hospital to home: a scoping review^134^ | Transition services to back home needed as patients have lost routines and with this the capacity for maintaining social connections. |
| Intermediates' satisfaction with a loneliness intervention program aimed at older adults: Linkage of program plans and users' needs^175^ | Democratic linkage strategies, like needs assessments, local action plans and two-way communication between program designers and users, are essential to support attendance to an intervention. |
| Interventions against social isolation of older adults: A systematic review of existing literature and interventions^202^ | Targeted, tailored, person-centred interventions effective. |
| Interventions Aimed at Alleviating Loneliness and Social Isolation among the Older Population: Perspectives of Service Providers^114^ | Important characteristics of the service - person-centred approach (personalisation), empowerment of service users and staff/volunteers (potentially through co-production), personal qualities of staff and volunteers*,* and funding (well run),  family and community structures, and accessible, affordable, and safe public transport (built on strengths of community). |
| Interventions to reduce loneliness in caregivers: An integrative review of the literature^284^ | Trained befrienders needed (thus part of a well-run organisation). Peer support needed to facilitate empathy and support from people in similar situations (shared experiences). |
| Interventions to reduce social isolation and loneliness among older people: an integrative review^16^ | Adaptability (as part of a well-run organisation), a community development approach, and productive engagement (active participation) are best for successful interventions. |
| Learning through Practice: How Can We Address Loneliness among Older People?^100^ | Community asset based approaches recommended. Also need to understand the nature of an individual’s loneliness and developing a personalised response, and  support lonely individuals to access services. |
| Loneliness and congregational social work^73^ | Interventions involving older adults in meaningful social activities within their communities may build and enhance social networks. |
| Loneliness and later life: Concepts, prevalence, and consequences^287^ | Not acknowledging loneliness in later life as a heterogeneous experience has contributed to the failure to develop effective interventions (as no personalisation). |
| Loneliness and social isolation interventions for older adults: A scoping review of reviews^10^ | Concludes there is no one-size-fits-all approach to interventions. Should instead consider what works for whom, in what particular context and how (so related to personalisation and cultural awareness). |
| Loneliness in older age: What is it, why is it happening and what should we do about it in Australia?^227^ | Interventions that are tailored (personalised), routine, and facilitate contact with a person or animal show most promise. |
| Loneliness in older people and COVID-19: Applying the social identity approach to digital intervention design^269^ | Social Identity Model of Identity Change – facilitate identifying with a group to prevent or alleviate loneliness. |
| Loneliness, coping practices and masculinities in later life: Findings from a study of older men living alone in England^293^ | Individual focused interventions missing – these would support men with more serious loneliness on a more personalised level. |
| Loneliness, older people and a proposed social work response^142^ | Direct assessment of needs required for effective intervention (support to attend) |
| Making connections - Reducing loneliness and encouraging well-being^204^ | Active engagement from staff/volunteers and  a way to find people and offer support most vital. |
| Older Adults and Social Isolation and Loneliness During the COVID-19 Pandemic: An Integrated Review of Patterns, Effects, and Interventions^169^ | Highlights need for age-friendly interventions (individual strengths) and strength-based approaches (community). |
| Older Adults’ Use of a Research-Based Web Platform for Social Interaction^140^ | Involvement of social services personnel in recruitment and support to attend was important. |
| Participant and Public Involvement in Refining a Peer-Volunteering Active Aging Intervention: Project ACE (Active, Connected, Engaged)^295^ | Barriers - lack of someone to attend with, lack of confidence, fear of exclusion or "cliquiness" in established groups, bad weather, transport issues, inaccessibility of activities, ambivalence, and older adults being "set in their ways" (all indicate support to attend). |
| People over 65 Years Old in Social Isolation: Description of an Effective Community Intervention in the City of Madrid (Spain)^255^ | Described need for individualized interventions, with a home-based approach by professionals, serving as a link between the older person and the normalized social-sanitary network. |
| Place and wellbeing: Shedding light on activity interventions for older men^212^ | Inclusionary social spaces and supportive social ties (culturally aware and shared identity). |
| Preventing social isolation and loneliness among older people: A systematic review of health promotion interventions^70^ | Good - volunteer visitor and the ‘service recipient’ belong to the same generation, have common interests, and share a common culture and social background (shared identity and well-run organisation). Programmes that enable older people to be involved in planning, developing and delivering activities are most likely to be effective (co-produced). |
| Primary care-based interventions addressing social isolation and loneliness in older people: a scoping review^129^ | Barriers – workload (not well run), lack of interest (no shared interest), ageing-related (not built on strengths of individual). Good - well-defined pathways, accessible (support to attend), collaborative designs (co-produced). |
| Promoting the empowerment and emancipation of community-dwelling older adults with chronic multimorbidity through a home visiting programme: a hermeneutical study^103^ | Personalising via trusting relationship with home visitor empowers to alleviate loneliness. |
| Psychosocial group rehabilitation for lonely older people: Favourable processes and mediating factors of the intervention leading to alleviated loneliness^257^ | Sharing experiences inspired lively discussions created a feeling of togetherness. |
| Pulling out all the stops: What motivates 65+ year olds with depressive symptoms to participate in an outreaching preference-led intervention programme?^281^ | Many participating individuals did not see a match between the intervention programme and their needs, especially re. meeting new people (so need to be personalised and co-produced). |
| Scenario-based Co-design with Older Adults: A Design Case on Decreasing Loneliness^280^ | Can be a mismatch between user needs and intervention design (so should co-produce). |
| Shared Interest Groups (SHIGs) in low-income independent living facilities^92^ | Shared interest groups effective as point for bonding. |
| Sing4Health: Randomised controlled trial of the effects of a singing group program on the subjective and social well-being of older adults^128^ | Participants identified with group thus less loneliness. |
| Smart Speaker and ICT Use in Relationship with Social Connectedness during the Pandemic: Loneliness and Social Isolation Found in Older Adults in Low-Income Housing^86^ | Found a strong desire among attendees to build community together. |
| Social Isolation and Psychological Distress Among Older Adults Related to COVID-19: A Narrative Review of Remotely-Delivered Interventions and Recommendations^133^ | Recomends services consisting of: active engagement; flexibility (among staff); individualised (personalised); involvement of older adults in development and implementation (co-production). |
| Social Isolation in Older Adults: A Qualitative Study on the Social Dimensions of Group Outdoor Health Walks^156^ | Engenders pleasurable anticipation of regular contact with others (routine), facilitates empathy for others (shared identity through conversation). |
| Staying Connected During a Global Pandemic: Telephone Support for Vulnerable Populations^285^ | The need for flexibility to make service change (well-run organisation). The regular calls became part of the rhythm of life providing certainty (routine). |
| Student-senior isolation prevention partnership: a Canada-wide programme to mitigate social exclusion during the COVID-19 pandemic^216^ | Community partnership model – organization of group important (well-run). |
| Tackling Loneliness and Isolation in Older Adults With Virtual Reality: How do We Move Forward?^117^ | Co-creation vital. |
| Tackling social disconnection: an umbrella review of RCT-based interventions targeting social isolation and loneliness^144^ | Tailored mitigating strategies (personalised) that address specific types and causes of loneliness are recommended. |
| Targeting Socially Isolated Older Adults: A Process Evaluation of the Senior Centre Without Walls Social and Educational Program^219^ | Hearing about project in the first place (support to attend) an important consideration. Positive as provides something to look forward to (routine). |
| Technology-Based Interventions to Address Social Isolation and Loneliness Among Informal Dementia Caregivers: A Scoping Review^203^ | A tailored approach to incorporate dynamic, flexible, and personalized technology-based interventions are desirable. |
| The contribution of primary care practitioners to interventions reducing loneliness and social isolation in older people-An integrative review^277^ | Identifier/referrer, assessor, responder and supporter as key components of support to attend. |
| The role of context and the interpersonal experience of loneliness among older people in a residential care facility^253^ | Good impacts of intervention - regular contact (routine). Bad – some attendees taking a controlling position (so should co-produce and personalise for all attendees). |
| The value of social eating at culturally and linguistically diverse lunch clubs: a descriptive study^210^ | Clubs were a rare opportunity for social interaction, maintaining ties to culture in a foreign country, and fostering health and wellbeing. The lunch club is a routine. |
| Understanding Acceptability of Group Leisure Activities Used to Address Loneliness Among People Living With Dementia: An Exploratory Mixed-Methods Study^226^ | Required for success interventions - flexible programs, attention to activity selection,  careful facilitation (support to attend). |
| Video-calls to reduce loneliness and social isolation within care environments for older people: An implementation study using collaborative action research^298^ | Barriers to success included staff turnover, risk averseness, staff attitudes (all represent whether a well-run organisation). |
| What do older people experiencing loneliness think about primary care or community based interventions to reduce loneliness? A qualitative study in England^172^ | Shared interest groups are effective. Loneliness when older is a bigger problem as practitioners lack understanding (indicating ineffective staff, so linked to well-run organisation). |
