## Supplementary file 7 for "Conceptualising the context and mechanisms for tackling loneliness in older adults through interventions: A Critical Interpretive Synthesis"

Supplementary file 7. Articles indicating a need for interventions to have a lasting impact.

| **Article title** | **Reason for inclusion** |
| --- | --- |
| Addressing Loneliness in Older People Through a Personalized Support and Community Response Program^208^ | Interventions have more chance of being effective if they resonate with and sustain engagement. |
| Effectiveness of home-based interventions in improving loneliness and social connectedness among older adults: a systematic review and meta-analysis^85^ | Interventions which lasted more than three months and were delivered using mixed platforms were more favourable. |
| Primary care-based interventions addressing social isolation and loneliness in older people: a scoping review^129^ | Emphasises sustainability as vital. |
| Psychological interventions for loneliness and social isolation among older adults during medical pandemics: a systematic review and meta-analysis^192^ | Interventions targeting social skills and the elimination of negativities were effective but only short term. |
| Change and stability in loneliness and friendship after an intervention for older women^205^ | Recovery from loneliness after a year was associated with the presence of a friend in the outer circle of the convoy and having more variation in one's friendships initially and one year later. It was also associated with the presence of a friend in the inner circle and reporting improvement in friendship. The maintenance of companionate friendship and the development of intimacy in one's friendships are therefore advantageous for recovery from loneliness. |
| Verification of the Effectiveness of a Communication Application in Improving Social Connectedness and Physical Health among Unacquainted Older Men: A Mixed-Methods Pilot Study^261^ | Relationships advanced as attendees’ understanding of each other’s personalities deepened. |
