## Supplementary file 8 for "Conceptualising the context and mechanisms for tackling loneliness in older adults through interventions: A Critical Interpretive Synthesis"

Supplementary file 8. Articles indicating a need to target interventions appropriately.

| **Article title** | **Reason for inclusion** |
| --- | --- |
| A systematic scoping review of community-based interventions for the prevention of mental ill-health and the promotion of mental health in older adults in the UK^185^ | Highlights a lack of ability to identify and appropriately target the ‘at risk’ among existing literature. |
| Attuning to the needs of structural socially isolated older adults with complex problems: the experiences of social workers with personal guidance trajectories for a less-researched group^198^ | Intervention not as positive results as many studies, but authors argue it may be better at identifying people particularly in need who are unlikely to attend many interventions. |
| Effects of tai chi qigong on psychosocial well-being among hidden elderly, using elderly neighborhood volunteer approach: A pilot randomized controlled trial^71^ | Argues a need to find the ‘hidden elderly’ (albeit the extent to which they managed to do this is unclear). |
| Evaluation of an intervention targeting loneliness and isolation for older people in North Wales^251^ | Notes that lonely participants often ‘didn’t want to ask’ (for help or anything related to their loneliness). |
| Interventions Aimed at Alleviating Loneliness and Social Isolation among the Older Population: Perspectives of Service Providers^114^ | Characteristics of the wider community were services were located were important as it could be hard to identify and/or reach lonely people. Suggests need for: education and awareness of services; family and community structures; and accessible, affordable, and safe public transport. |
| Learning through Practice: How Can We Address Loneliness among Older People?^100^ | Argues interventions should consider how best to reach, identify and support older people. |
| Loneliness: Possibilities for intervention^163^ | Argues the attendance the truly isolated and lonely person rarely attends interventions. |
| Making connections - Reducing loneliness and encouraging well-being^204^ | A way to find people and offer support is vital. |
| Systematic Review of Efficacy of Interventions for Social Isolation of Older Adults^279^ | Accurate targeting of clients in social and public places is required. |
| Targeting Socially Isolated Older Adults: A Process Evaluation of the Senior Centre Without Walls Social and Educational Program^219^ | Hearing about project in the first place is vital for lonely individuals. |
| The contribution of primary care practitioners to interventions reducing loneliness and social isolation in older people-An integrative review^277^ | Primary care practitioners can contribute by being an identifier/referrer, assessor, responder and supporter. |
| The role of assistive technology in addressing social isolation, loneliness and health inequities among older adults during the COVID-19 pandemic^168^ | Assistive technology may benefit the least in need first as it requires trust in and access to technology. |
