## Supplementary file 9 for "Conceptualising the context and mechanisms for tackling loneliness in older adults through interventions: A Critical Interpretive Synthesis"

Supplementary file 9. Articles evidencing that reciprocity in relationships results in reduced loneliness

| **Article title** | **Reason for inclusion** |
| --- | --- |
| Aging and social isolation: An integrative review^54^ | Most effective when target people who can share similar experiences with each other. |
| Being cut off from social identity resources has shaped loneliness during the coronavirus pandemic: A longitudinal interview study with medically vulnerable older adults from the United Kingdom^145^ | Reciprocal contributions important for successful intervention. |
| Evaluating Feasibility, Value and Characteristics of an Intergenerational Friendly Telephone Visit Program During the Covid-19 Pandemic^177^ | ‘Mutuality’ important for successful intervention. |
| Friendly Visiting Programs for Older People Experiencing Social Isolation: A Realist Review of what Works, for whom, and under what Conditions^289^ | New relationships form best if client volunteer match and people can form meaningful relationships. |
| How do befriending interventions alleviate loneliness and social isolation among older people? A realist evaluation study^113^ | Reciprocity highlighted as important for relationship building. |
| Intervention to reduce loneliness among older adults in the community: Making friends with volunteers^153^ | Volunteers who were considered friends resulted in less loneliness. |
| It Gave Me Somebody Else to Think About Besides Myself: Caring Callers Volunteer Experiences With a Telephone-Based Reassurance Program for Socially Isolated Older Adults^116^ | Reciprocity highlighted. |
| Key Elements and Mechanisms of a Peer-Support Intervention to Reduce Loneliness and Isolation among Low-Income Older Adults: A Qualitative Implementation Science Study^125^ | Staff perceived as friends resulted in more effective interventions. |
| Older Adults’ Views on Social Interactions and Online Socializing Games–A Qualitative Study^159^ | ‘Reciprocity’, ‘in-person contact’, and ‘personal connection’ interrelated useful factors for reducing loneliness. |
| The Primacy of Compassionate Love: Loneliness and Psychological Well-Being in Later Life^187^ | Feel loved = less lonely. So interventions aimed at this rather than broader networks likely to work better. |
| The role of context and the interpersonal experience of loneliness among older people in a residential care facility^253^ | A lack of confirmatory interpersonal relationships result in an unsuccessful intervention. It is recommended that greater emphasis should be placed on creating awareness of unhealthy group dynamics. |
