## Supplementary file 10 for "Conceptualising the context and mechanisms for tackling loneliness in older adults through interventions: A Critical Interpretive Synthesis"

Supplementary file 10. Articles evidencing that emotional or social support results in reduced loneliness

| **Article title** | **Reason for inclusion** |
| --- | --- |
| A realist evaluation of loneliness interventions for older people^98^ | Providing emotional support as part of what facilitates a successful intervention. |
| COVID-19 impacts and interventions for older adults: implications for future disasters^213^ | Grief support as part of what facilitates a successful intervention. |
| Friendly Visiting Programs for Older People Experiencing Social Isolation: A Realist Review of what Works, for whom, and under what Conditions^289^ | Provision of informal social support and provision of mediated social support as part of what facilitates a successful intervention. |
| Loneliness and social support of older people living alone in a county of Shanghai, China^77^ | Found a negative correlations between loneliness and overall social support and its three dimensions (objective support, subjective support, support utilisation). |
| Loneliness matters: a theoretical and empirical review of consequences and mechanisms^146^ | Provision of social support part of what is needed for a successful reduction in loneliness. |
| Mechanisms through which befriending services may impact the health of older adults: A dyadic qualitative investigation^143^ | Providing emotional support part of what facilitates a successful intervention. |
| Systematic Review of Efficacy of Interventions for Social Isolation of Older Adults^279^ | Group intervention activities and individual intervention interviews were effective in improving social support. |
| The effectiveness of remote delivered intervention for loneliness reduction in older adults: A systematic review and meta-analysis^123^ | Facilitating social support was an important component of successful remote interventions. |
| Understanding and Addressing Older Adults’ Loneliness: The Social Relationship Expectations Framework^42^ | Receiving care and support an important need from relationships. |
