## Supplementary file 11 for "Conceptualising the context and mechanisms for tackling loneliness in older adults through interventions: A Critical Interpretive Synthesis"

Supplementary file 11. Articles evidencing that a sense of belonging results in reduced loneliness.

| **Article title** | **Reason for inclusion** |
| --- | --- |
| “It makes me feel not so alone”: features of the Choose to Move physical activity intervention that reduce loneliness in older adults^121^ | Articles concludes opportunity to share information and experiences and learn from others, engagement with others who share similar/familiar experiences, and increased opportunity for meaningful interaction had a positive impact on loneliness. |
| Effects of a ‘social activity program that encourages interaction’ on rural older people's psychosocial health: Mixed-methods research^173^ | Articles concludes ‘stimulation brought about by relationships with peers,’ ‘realization as to where they feel they belong,’ ‘rethinking of oneself in the community,’ and ‘awareness of attachment to and coexistence with the community’ had a positive impact on loneliness. |
| Evaluating Feasibility, Value and Characteristics of an Intergenerational Friendly Telephone Visit Program During the Covid-19 Pandemic^177^ | Mutuality and respect have a positive impact on loneliness. |
| Exploring existential loneliness among frail older people as a basis for an intervention: Protocol for the development phase of the LONE study^109^ | Loneliness portrayed as a feeling of being fundamentally separated from others (thus represents the opposite of belonging). |
| Piloting the use of football club community Trust's to create social Hubs for older adults^63^ | Study concludes it can work because people comfortable being there as it is a familiar and welcoming environment. |
| Reminiscence interventions for loneliness reduction in older adults: a systematic review^69^ | In group reminiscence interventions, the levels of loneliness might be reduced partly as a result of the group effect rather than the effect of the reminiscence itself. |
| Smart Speaker and ICT Use in Relationship with Social Connectedness during the Pandemic: Loneliness and Social Isolation Found in Older Adults in Low-Income Housing^86^ | Found a strong desire to build community together in their facilities and highlighted the potential role of smart speakers in making meaningful social connections. |
| Social Isolation in Older Adults: A Qualitative Study on the Social Dimensions of Group Outdoor Health Walks^156^ | Fosters casual interpersonal interactions and group cohesion. These contribute to expanding social networks, meaningful relationships, and a sense of belonging. |
| The ‘loneliness pandemic’: Implications for gerontological nursing^181^ | Passive use of social networks not effective as it does not contribute to the sense of belonging. |
| The role of context and the interpersonal experience of loneliness among older people in a residential care facility^253^ | A lack of confirmatory interpersonal relationships (a lack of belonging) a barrier to positive interventions. Greater emphasis should be placed on awareness of unhealthy Group dynamics. |
