## Supplementary file 12 for "Conceptualising the context and mechanisms for tackling loneliness in older adults through interventions: A Critical Interpretive Synthesis"

Supplementary file 12. Articles evidencing that gaining a sense of perspective results in reduced loneliness

| **Article title** | **Reason for inclusion** |
| --- | --- |
| Evaluating Feasibility, Value and Characteristics of an Intergenerational Friendly Telephone Visit Program During the Covid-19 Pandemic^177^ | Broadened perspective highlighted by participants as useful for reducing loneliness. |
| It Gave Me Somebody Else to Think About Besides Myself: Caring Callers Volunteer Experiences With a Telephone-Based Reassurance Program for Socially Isolated Older Adults^116^ | Gaining perspective highlighted by participants as useful for reducing loneliness. |
| The effectiveness of group reminiscence therapy for loneliness, anxiety and depression in older adults in long-term care: A systematic review^271^ | Helped people gain personal value and self-identity. |
| Combating loneliness: A friendship enrichment programme for older women^267^ | Helped women clarify their needs in friendship and analyse their current social network. |
