## Supplementary file 13 for "Conceptualising the context and mechanisms for tackling loneliness in older adults through interventions: A Critical Interpretive Synthesis"

Supplementary file 13. Articles evidencing that increased an increased sense of self-efficacy results in reduced loneliness.

| **Article title** | **Reason for inclusion** |
| --- | --- |
| ‘It’s not just about the dinner; it’s about everything else that we do’: A qualitative study exploring how Meals on Wheels meet the needs of self-isolating adults during COVID-19^232^ | Service encourages independence which is purported to be indirectly beneficial as it allows for greater self-perception. |
| Benefits of a psychosocial intervention programme using volunteers for the prevention of loneliness among older women living alone in Spain^194^ | Attribution retraining and behavioural activation facilitated self-efficacy in managing loneliness. |
| Evaluation of an intervention in general practices to strengthen social activities in older patients - A qualitative study of patients' experiences in the project HoPES3^199^ | Increasing self-efficacy and quality of life via raising awareness of spiritual needs, promoting social contacts and self-care. Discussion section suggests are inter-related factors. |
| Online Social Networking and Mental Health among Older Adults: A Scoping Review^78^ | Greater independence and self-efficacy portrayed as a positive impact of online social networking. |
| The Effectiveness of a Thanks, Sorry, Love, and Farewell Board Game in Older People in Taiwan: A Quasi-Experimental Study^74^ | Playing the game led to improvements in scores on interpersonal communication, self-efficacy, and loneliness. |
| Volunteering and loneliness in older adults: A parallel mediation model^184^ | Volunteer work predicted perceived control, social self-efficacy, and lower loneliness. Perceived control and social self-efficacy mediate the relationship between volunteer activities and loneliness. |
