## Supplementary file 14 for "Conceptualising the context and mechanisms for tackling loneliness in older adults through interventions: A Critical Interpretive Synthesis"

Supplementary file 14. Articles evidencing that a feeling of mattering results in reduced loneliness

| **Article title** | **Reason for inclusion** |
| --- | --- |
| Effects of a ‘social activity program that encourages interaction’ on rural older people's psychosocial health: Mixed-methods research^173^ | Themes titled ‘rethinking of oneself in the community,’ and ‘awareness of attachment to and coexistence with the community’ indicate a sense of mattering to people. |
| Evaluating Feasibility, Value and Characteristics of an Intergenerational Friendly Telephone Visit Program During the Covid-19 Pandemic^177^ | Themes of mutuality and respect indicate a sense of mattering to people. |
| Piloting the use of football club community Trust's to create social Hubs for older adults^63^ | Uses quote of a person saying ‘you feel wanted, they ask after you’ to emphasise positive components of intervention. |
| Qualitative evaluation of a community-based intervention to reduce social isolation among older people in disadvantaged urban areas of Barcelona^180^ | Giving people a feeling of being heard was highlighted as a positive component of the intervention. |
| The Primacy of Compassionate Love: Loneliness and Psychological Well-Being in Later Life^187^ | Feel loved = less lonely. Logically, if you feel loved, you also feel you matter to the person that loves you. |
| The role of context and the interpersonal experience of loneliness among older people in a residential care facility^253^ | Taking a controlling position in relationships and being rigid elicited feelings of rejection (thus not mattering). It is recommended that greater emphasis should be placed on creating awareness of unhealthy group dynamics. |
