## Supplementary file 15 for "Conceptualising the context and mechanisms for tackling loneliness in older adults through interventions: A Critical Interpretive Synthesis"

Supplementary file 15. Articles evidencing that pride results in reduced loneliness

| **Article title** | **Reason for inclusion** |
| --- | --- |
| 'A lonely old man': Empirical investigations of older men and loneliness, and the ramifications for policy and practice^8^ | Good interventions should facilitate the construction of a 'proud' masculine identity through purpose and helping out, and provide a space where men feel comfortable being emotionally tactile. Also suggests perceived social worth, reducing shame, and stigma are important |
| Being cut off from social identity resources has shaped loneliness during the coronavirus pandemic: A longitudinal interview study with medically vulnerable older adults from the United Kingdom^145^ | Recommends addressing age-based stereotypes to tackle loneliness. |
| Developmental Study on “Smart Silver Care”: A Mobile Application to Alleviate Loneliness in Older Adults within the Community^80^ | Expected to help older adults achieve their goals by promoting participation. Achieving and non-loneliness constructed as interrelated. |
| Effects of horticultural therapy on psychosocial health in older nursing home residents: A preliminary study^76^ | A sense of achievement found to be a positive result of intervention. Not directly linked to loneliness, but study also had a positive impact on loneliness. |
| Intergenerational Group Reminiscence: A Potentially Effective Intervention to Enhance Elderly Psychosocial Wellbeing and to Improve Children's Perception of Aging^127^ | Sharing stories of the past and meaningful life events provided participants with feelings of togetherness and intimacy. |
| Online Social Networking and Mental Health among Older Adults: A Scoping Review^78^ | Loneliness had positive associations with well-being and life satisfaction |
| Psychosocial group rehabilitation for lonely older people: Favourable processes and mediating factors of the intervention leading to alleviated loneliness^257^ | Sharing experiences inspired lively discussions, created a feeling of togetherness and led to empowerment and self-esteem. |
| The effectiveness of group reminiscence therapy for loneliness, anxiety and depression in older adults in long-term care: A systematic review^271^ | Intervention led to gains in personal value and self-identity. |
| Understanding and Addressing Older Adults’ Loneliness: The Social Relationship Expectations Framework^42^ | Highlights generativity and contribution, and being respected and valued, as needed from relationships to reduce loneliness. |
