## Supplementary file 16 for "Conceptualising the context and mechanisms for tackling loneliness in older adults through interventions: A Critical Interpretive Synthesis"

Supplementary file 16. Articles evidencing that a sense of purpose results in reduced loneliness

| **Article title** | **Reason for inclusion** |
| --- | --- |
| Associations Between Changes in Loneliness and Social Connections, and Mental Health During the COVID-19 Pandemic: The Women’s Health Initiative^136^ | Having higher purpose in life statistically associated with less loneliness. |
| Being disconnected from life: meanings of existential loneliness as narrated by frail older people^262^ | Being met with indifference and lacking purpose and meaning linked to loneliness. Interventions should develop strategies to support meaning in life. |
| Effect of Combining Adapted Physical and Artistic Activities on Feeling of Loneliness and Aggression in the Elderly Living in Nursing Homes^259^ | Art-based group therapy creates an interactive and encouraging environment. Art creation can be interpreted as a purpose facilitating the reductions in loneliness. |
| Intergroup ‘skype’ quiz sessions in care homes to reduce loneliness and social isolation in older people^300^ | Re-gaining sense of self and purpose one of the benefits of the intervention. |
| Interventions to address social connectedness and loneliness for older adults: a scoping review^225^ | One of the strategies described most often were engaging in purposeful activity. |
| It Gave Me Somebody Else to Think About Besides Myself: Caring Callers Volunteer Experiences With a Telephone-Based Reassurance Program for Socially Isolated Older Adults^116^ | purposeful use of time and learning new skills highlighted as helpful. |
| Managing loneliness: a qualitative study of older people’s views^171^ | Engagement with purpose in an ‘outside world’ highlighted as helpful. |
| The Impact of Robotic Companion Pets on Depression and Loneliness for Older Adults with Dementia During the COVID-19 Pandemic^119^ | Qualitative data suggests was a purposeful thing to do as people engaged with caring for the robot. |
| The Leveraging Exercise to Age in Place (LEAP) Study: Engaging Older Adults in Community-Based Exercise Classes to Impact Loneliness and Social Isolation^206^ | Purpose-driven activity designed to promote physical activity and social interaction.  Showed a statistically significant reduction in loneliness. |
| The Management of Loneliness in Aged Care Residents: An Important Therapeutic Target for Gerontological Nursing^60^ | Contributing to family, social, community, and institutional life preserves the feeling of being valued and helpful, and still serving a purpose. |
| Voluntary sector interventions to address loneliness and mental health in older people: taking account of emotional, psychological and social wellbeing^99^ | Groups are better for forming networks and feeling person contributes to a community. |
| Verification of the Effectiveness of a Communication Application in Improving Social Connectedness and Physical Health among Unacquainted Older Men: A Mixed-Methods Pilot Study^261^ | Measuring step count added purpose to intervention, which bettered engagement. Study didn’t frame as a direct mediator of less loneliness, but based on other work included in the current review, this interpretation is also viable. |
