## Supplementary file 17 for "Conceptualising the context and mechanisms for tackling loneliness in older adults through interventions: A Critical Interpretive Synthesis"

Supplementary file 17. Articles evidencing that a feeling of empowerment results in reduced loneliness

| **Article title** | **Reason for inclusion** |
| --- | --- |
| ‘It’s not just about the dinner; it’s about everything else that we do’: A qualitative study exploring how Meals on Wheels meet the needs of self-isolating adults during COVID-19^261^ | Encouraging independence happened as well as identifying and addressing loneliness in the intervention. May be interrelated benefits to intervention. |
| Being cut off from social identity resources has shaped loneliness during the coronavirus pandemic: A longitudinal interview study with medically vulnerable older adults from the United Kingdom^145^ | Addressing age-based stereotypes able to counteract loneliness. |
| Can digital technology enhance social connectedness among older adults? A feasibility study^50^ | Some technology highlighted feelings of inadequacy and awareness of frailty, therefore has a negative impact on loneliness. |
| Combating loneliness: A friendship enrichment programme for older women^267^ | Study facilitated empowerment by helping women clarify their needs in friendship, analyse their current social network, set goals in friendship and develop strategies to achieve goals. |
| Conditions for Feasibility of a Multicomponent Intervention to Reduce Social Isolation and Loneliness in Noninstitutionalized Older Adults^147^ | Interventions with a clear purpose focused on empowerment were identified as beneficial to tackling loneliness. |
| How to fulfil social needs of older people: Exploring design opportunities for technological interventions^274^ | Facilitating independence is a factor in design able to facilitate social connectedness. Also may relate to general confidence in interactions. |
| Interventions against social isolation of older adults: A systematic review of existing literature and interventions^202^ | Interventions that empower and motivate people, and which enable participants to control loneliness, are among the most effective interventions. |
| Interventions Aimed at Alleviating Loneliness and Social Isolation among the Older Population: Perspectives of Service Providers^114^ | Empowerment of service users and staff/volunteers had a positive effect on loneliness of service users. |
| Online acceptance and commitment therapy as treatment for loneliness among older adults: Report of a pilot study^301^ | Teach participants skills to empower the patient to alter behaviours in service of his or her values. |
| Promoting the empowerment and emancipation of community-dwelling older adults with chronic multimorbidity through a home visiting programme: a hermeneutical study^103^ | Personalised services, enacted via trusting relationship with home visitor, empowers the older adult to act on and improve their loneliness. |
| Psychosocial group rehabilitation for lonely older people: Favourable processes and mediating factors of the intervention leading to alleviated loneliness^257^ | Sharing experiences inspired lively discussions, created a feeling of togetherness and led to empowerment and self-esteem. |
| Pulling out all the stops: What motivates 65+ year olds with depressive symptoms to participate in an outreaching preference-led intervention programme?^281^ | Many participating individuals did not see a match between the intervention programme and their needs, especially regarding meeting new people. As such, it was a disempowering experience. |
| Solitude in old age: a scoping review of conceptualisations, associated factors and impacts^228^ | Promoting “positive solitude” and destigmatizing time spent alone can empower older adults to choose fulfilling activities and connect with themselves. |
| Student-senior isolation prevention partnership: a Canada-wide programme to mitigate social exclusion during the COVID-19 pandemic^216^ | Community partnership model - empowerment, behaviour change, and good organization of services combine to tackle loneliness. |
| When Technologies are Not Enough: The Challenges of Digital Interventions to Address Loneliness in Later Life^51^ | Stigma, ageism, social trust, and lack of privacy impact experience of technology use in disempowering ways, thus can increase on loneliness. |
